# Anti-inflammatory therapy with nebulised dornase alfa for severe COVID-19 pneumonia

**DOI:** 10.1101/2022.04.14.22272888

**Authors:** Joanna C. Porter, Jamie Inshaw, Vincente Joel Solis, Emma Denneny, Rebecca Evans, Mia I. Temkin, Nathalia De Vasconcelos, Iker Valle Aramburu, Dennis Hoving, Donna Basire, Tracey Crissell, Jesusa Guinto, Alison Webb, Hanif Esmail, Victoria Johnston, Anna Last, Thomas Rampling, Elisa Theresa Helbig, Lena Lippert, Florian Kurth, Bryan Williams, Aiden Flynn, Pauline T Lukey, Veronique Birault, Venizelos Papayannopoulos

**Affiliations:** UCL Respiratory, University College London, UK; University College London Hospitals NHS Trust, London, UK; Exploristics, Belfast, N. Ireland; Antimicrobial Defence lab, The Francis Crick Institute, London, UK; National Institute for Health Research, University College London Hospital Biomedical Research Centre, UK; Clinical Research Department, London School of Hygiene and Tropical Medicine, London, UK; Charité – Universitätsmedizin Berlin, Department of Infectious Diseases and Respiratory Medicine, Berlin, Germany; Target to Treatment Consulting Ltd, Stevenage, UK; Translation, The Francis Crick Institute, London, UK

**Keywords:** COVID-19, SARS-CoV-2, endonuclease, Dornase, DNA, virus, infection, pneumonia, anti-inflammatory, DNAse, NETs, neutrophil, neutrophil extracellular traps

## Abstract

**Background:** Cell-free (cf)-DNA, from cellular sources, including Neutrophil Extracellular Traps (NETs), is found in the circulation of COVID-19 patients and may contribute to immune dysregulation. This study determined whether pulmonary administration of the endonuclease, dornase alfa, reduced systemic inflammation by degrading local and systemic cf-DNA.

**Methods:** Eligible patients were randomized (3:1) to receive twice-daily nebulised dornase alfa in addition to best available care (BAC) or BAC alone for seven days or until discharge. A 2:1 ratio of matched contemporary controls (CC-BAC) provided additional comparators. The primary endpoint was improvement in C-reactive protein (CRP) over time, analysed using a repeated-measures mixed model, adjusted for baseline factors.

**Results:** Between June 2020-October 2021 we recruited 39 evaluable patients: 30 randomised to dornase alfa (R-BAC+DA); 9 randomised to BAC (R-BAC); with the addition of 60 CC-BAC participants. Dornase alfa was well tolerated and reduced CRP by 33% compared to combined BAC groups (T-BAC). Least squares (LS) mean post-dexamethasone CRP fell from 101.9mg/L to 23.23 mg/L in the BAC+ dornase alfa group versus a fall from 99.5mg/L to 34.82 mg/L in the T-BAC group at 7 days; P=0.01. This effect of dornase alfa on CRP was confirmed with subgroup and sensitivity analyses that mitigated potential biases associated with the use of the CC-BAC group. Dornase alfa increased the chance of live discharge by 63% (HR 1.63, 95% CI 1.01 to 2.61, P=0.03), increased lymphocyte counts (LS mean: 1.08 vs 0.87, P=0.02) and reduced circulating cf-DNA and the coagulopathy marker D-dimer (LS mean: 570.78 vs 1656.96 μg/mL, P=0.004).

**Conclusion:** We provide proof-of-concept evidence that dornase alfa reduces pathogenic inflammation in hospitalised patients with COVID-19 pneumonia, suggesting that best available care can be improved by the inclusion of anti-inflammatory treatments that target damage-associated molecules.

## INTRODUCTION

SARS-CoV-2 pneumonia can lead to hyperinflammation, coagulopathy, respiratory failure, and death (Siddiqi and Mehra, 2020; Zhou et al., 2020), even with less pathogenic variants and in immunised patients (2000). In severe COVID-19 neutrophil activation drives neutrophil extracellular trap (NET) formation (Radermecker et al., 2020; Zuo et al., 2020). NETs, are composed of DNA-histone complexes and have been associated with coagulopathy and endothelial dysfunction (Papayannopoulos, 2018). Moreover, extracellular histones promote inflammation, immune dysfunction and lethality in sepsis (Ioannou et al., 2022; Tsourouktsoglou et al., 2020; Xu et al., 2011; Xu et al., 2009). Digestion of chromatin DNA by endonucleases suppresses the proinflammatory activity of histones and enables their clearance from the circulation (Ioannou et al., 2022; Tsourouktsoglou et al., 2020). Consistently, DNAse I treatment reduces pathology in murine pulmonary viral infections (Cortjens et al., 2018; Pillai et al., 2016). Endogenous DNAse activity and NET clearance capacity are defective in severe COVID-19 pneumonia, and the extent of these defects correlates with mortality (Aramburu et al., 2022). Hence, supplementation with exogenous DNases may facilitate extracellular chromatin degradation and increase survival.

Pulmozyme, dornase alfa, is a recombinant human DNAase approved since 1993 for patients with cystic fibrosis (CF) (Konstan and Ratjen, 2012; Lazarus and Wagener†, 2019). Dornase alfa solubilizes NETs, reduces inflammation and improves pulmonary function in chronic and acute exacerbations of CF (Konstan and Ratjen, 2012; Papayannopoulos et al., 2011). Pulmozyme is safe and well-tolerated in children and adults with CF at doses up to 10mg BD. Nebulized pulmozyme does not increase circulating endonucleases. Hence, it remained unclear whether the degradation of extracellular chromatin in the lungs would reduce circulating chromatin and systemic hyper-inflammation.

To probe the effectiveness of DNase I as an anti-inflammatory therapy in severe COVID-19 pneumonia, we measured as our primary endpoint C-reactive protein (CRP). CRP is a reliable marker of systemic inflammation in severe infection (McArdle et al., 2004). In patients infected with COVID-19, a high CRP of ≥40 mg/L carried a poor prognosis with a lower cut-off of ≥35 mg/L in the elderly (Villoteau et al., 2021), particularly when combined with lymphopenia and coagulopathy marked by elevated D-dimer (Fisher et al., 2022; Liu et al., 2020; Luo et al., 2020; Smilowitz et al., 2021; Tornling et al., 2021; Ullah et al., 2020). Poor outcomes are associated with an early rise (Mueller et al., 2020), higher peak and delayed reduction in CRP (Cui et al., 2021). Moreover, elevated IL-6 and CRP can predict the need for mechanical ventilation(Herold et al., 2020). In addition, CRP correlated well with levels of DNAse and NET degradation activity in COVID-19 patient plasma suggesting a link between CRP and the capacity to degrade cf-DNA in the circulation (Aramburu et al., 2022). CRP became a reliable primary endpoint in subsequent trials demonstrating the effectiveness of other systemic therapies such as namilumab or infliximab that target inflammatory cytokines (Fisher et al., 2022).

In this proof-of-concept study we evaluated the effect of dornase alfa on inflammation, as measured by the impact on circulating CRP in patients with COVID-19, compared with best available care (BAC). The trial was initiated in June 2020 and was completed in October 2021. At the start of the trial only dexamethasone had been proven to benefit hospitalized COVID-19 pneumonia patients and was thus included in both arms of the trial. To increase the chance of reaching significance under challenging constraints in patient access, we opted to increase our sample size by using a combination of randomized individuals in the BAC group (R-BAC) and the dornase alfa treated arm (R-BAC+DA) with available CRP data from matched contemporary controls (CC-BAC) hospitalized at the same site at UCL but not recruited to a trial and using the same selection criteria as the randomised group to minimise potential biases. In the analysis framework we compared treated R-BAC-DA participants to both R-BAC and CC-BAC groups individually or combined (T-BAC). These approaches demonstrated that when combined with dexamethasone, nebulized DNase treatment was an effective anti-inflammatory treatment in randomized individuals with or without the implementation of CC-BAC data.

## METHODS

### Sponsor and location

The trial was sponsored by University College London (UCL) and carried out at University College London Hospital UCLH with ethical (REC: 20/SC/0197, Protocol: 132333, RAS ID:283091) and UK MHRA approvals. All randomized participants provided informed consent. Consent for CC-BAC participants was covered by Health Service (Control of Participant Information) Regulations 2002. Safety and data integrity were overseen by the Trial Monitoring Group and Data Monitoring Committee. All data was collected at UCLH. Additional CRP data is reported from the Pa-COVID-19 study, Charité Universitätsmedizin Berlin with ethical approval, Berlin (EA2/066/20). Both Pa-COVID-19 and COVASE studies were carried out according to the Declaration of Helsinki and the principles of Good Clinical Practice (ICH 1996).

### Trial design

The COVASE trial was a single-site, randomised, controlled, parallel, open-label investigation. The primary endpoint was change in CRP. Screening was performed within 24 hours prior to the administration of dornase alfa (**Figure 1A**). Eligible, consented participants were randomly assigned (3:1) to either BAC plus nebulised dornase alfa (R-BAC+DA) or R-BAC group alone. On Day 1 a baseline sample was collected. From Day 1-7 participants randomised to active arm received 2.5mg BD nebulised dornase alfa in addition to BAC. In all cases BAC included dexamethasone (6 mg/day) for 10 days or until discharge, whichever was shorter as per RECOVERY (Group et al., 2021). Participants received additional treatments at the discretion of their physicians. The primary analysis was performed on samples up to Day 7. The final trial visit occurred at day 35.

**Figure 1.**
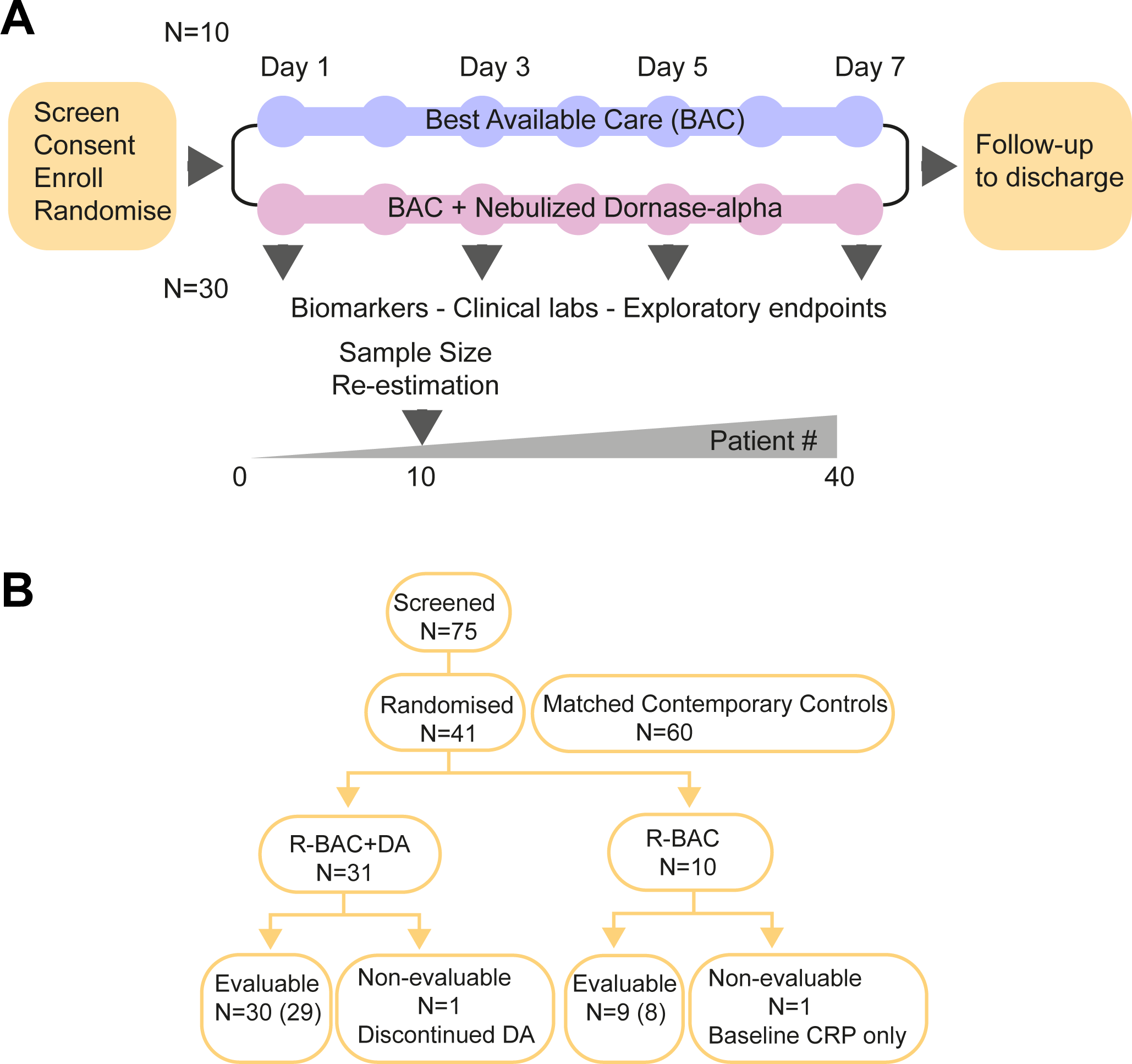
Trial design and Consort diagram. A. COVASE Trial Design. B. Consort diagram summary. Numbers not in parentheses indicate the participants in the ITT population and the numbers in parentheses indicate the number of participants in the per-protocol population. A complete consort diagram is shown in Supplementary figure 1.

In addition to the recruited randomised group, we included CC-BAC participants to increase the sample size due to practical difficulties in recruiting patients in the UK. For every COVASE participant randomised to active treatment, 2 matched CC-BAC participants were included. CC-BAC participants were admitted to UCLH over the same time as randomised patients and treated with the same BAC, including dexamethasone. CC-BAC participants fulfilled the inclusion and exclusion criteria of the COVASE study and were matched for age, gender, BMI, comorbidities and CRP. CRP matching was based either on admission CRP, or matched based on the first CRP reading after starting dexamethasone.

### Eligibility

#### Trial participant inclusion criteria

Male and female participants, aged ≥ 18 years, who were hospitalised for suspected Coronavirus (SARS-CoV)-2 infection confirmed by polymerase chain reaction (PCR) test and optionally with radiological confirmation with chest CT. Participants with stable oxygen saturation (>=94%) on supplementary oxygen and a CRP >= 30 mg/L. Participants that provided written informed consent to participate in the study and were able to comply with instructions and the use of a nebuliser.

#### Exclusion criteria

Females who were pregnant, planning pregnancy or breastfeeding. Concurrent and/or recent involvement in other research or use of another experimental investigational medicinal product that was likely to interfere with the study medication within 3 months of study enrolment. Serious condition meeting either respiratory distress with respiratory rate >=40 breaths/min, or an oxygen saturation<=93% on high-flow oxygen. Participants who required mechanical invasive or non-invasive ventilation at screening or had concurrent severe respiratory disease such as asthma, COPD and/or ILD, or any major disorder that in the opinion of the Investigator would interfere with the evaluation of the results or constitute a health risk for the trial participant. Participants with terminal disease and life expectancy <12 months without COVID-19, known allergies to dornase alfa and excipients, or participants who were unable to inhale or exhale orally throughout the entire nebulisation period.

### Participants

Adults (≥18 years of age) admitted to UCLH with confirmed SARS-Cov2 infection by RT-PCR and radiologically confirmed COVID-19 pneumonia on chest radiograph or CT-scan; oxygen saturation < 94% requiring supplemental oxygen; and evidence of hyperinflammation (CRP ≥ 30 mg/L, after administration of dexamethasone) were eligible. While all participants had CRP > 30 mg/L at screening, on two occasions the baseline CRP concentrations fell below 30 mg/L after the patients had already agreed to participate. For full inclusion and exclusion criteria see Protocol in the Supplementary Material.

### Outcomes

The primary outcome was the least square (LS) mean CRP up to 7 days or at hospital discharge whichever was sooner. Data beyond 7 days was included in the model to estimate the slope. Subsequently, the estimated slope in each group was used in order to calculate the LS mean at 7 days. Pre-specified secondary outcomes included days on oxygen; time to hospital discharge; mortality by day 35; and changes in clinically relevant biomarkers including lymphocyte count and D-dimer levels. Day 35 was chosen as being likely to include most early mortality due to COVID-19 being 4 weeks after completion of a week of treatment. (i.e. d7 of treatment +28 (4 x 7 days)).

Efficacy assessments of primary and secondary outcomes in the intention-to-treat (ITT) population were performed on all randomised participants who had received at least one dose of dornase alfa if randomized to treatment. For full details see Statistical Analysis Plan. The ITT was adjusted to mitigate the following protocol violations where one participant in the R-BAC arm and one in the R-BAC+DA arm withdrew before they received treatment and provided only a baseline CRP measurement available. The participant in the DA arm was replaced with an additional recruited patient. Exploratory endpoints were only available in randomised participants and not in the CC-BAC. In this case, a post hoc within group analysis was conducted to compare baseline and post-baseline measurements.

### Sample size calculation

Size calculations were produced using the proc power function in SAS Version 9.4. These were conducted to achieve 80% power to detect difference in the active arm versus the control group at 5% level of significance. Based on a mean of 99mg/L in the control group and a common standard deviation of 62mg/L derived from the literature (Han et al., 2020; Zhou, 2020), a total sample size of 90 participants would provide sufficient power to detect a greater than a 40% relative difference for CRP in the dornase alfa group compared to the control group.

This study used existing data collected at UCLH from CC-BAC participants admitted with COVID-19. This gave a ratio of active versus comparator of 1:2. The required power, would result in 30 participants in the active treatment group and at least 60 in the control group. An additional 10 radnomised control participants were recruited for exploratory objectives and to compare the characteristics of enrolled participants with CC-BAC participants. Therefore, a total of 40 participants were enrolled in the study and 60 CC-BAC participants were included.

### Randomisation

A closed envelope method was implemented to randomize the participants into the control and active treatment groups.

## RESULTS

### Patient charateristics

From June 2020-October 2021, 41 participants were recruited and randomised, with 1 participant in R-BAC group discharged before a second CRP measurement who was excluded from all analyses except for safety (CONSORT Diagram: **Figure 1B and S1**). One participant withdrew consent prior to receiving dornase alfa and was replaced and excluded from all analyses. 39 participants were included in the ITT analysis set, 30 R-BAC+DA and 9 R-BAC. One participant withdrew due to side-effects. This participant was removed from the per protocol population (PPP). All 39 participants were followed up for 35 days or death whichever was sooner. Two participants were excluded from the PPP and one from R-BAC as this was the only patient in whom randomisation occurred prior to dexamethasone being widely used in COVID-19 and a second participant who withdrew after one dose of dornase alfa (**Figure 1B and S1**). The trial ended when 40 eligible participants had been recruited, although thereafter 1 had participant in the R-BAC arm had to be excluded from the primary and secondary endpoint analysis.

Baseline characteristics were well balanced across groups (Table 1). Selection of 60 matched CC-BAC participants via propensity score matching was successful in ensuring the means of the characteristics included in propensity score matching were similar (**Table 1** and **Figure S2**). The overall mean age was 56.8 years (56.8 years R-BAC+DA, 56.8 years T-BAC). The percentage of males was 75.8% (76.7% R-BAC+DA, 75.4% T-BAC). The most prevalent ethnicity was “White British”, with 30.3% of participants identifying in that category (33.3% BAC+DA, 29.0% T-BAC). The mean BMI was 28.0kg/m2 (27.8kg/m2 R-BAC+DA, 28.2kg/m2 T-BAC). The mean baseline CRP (post dexamethasone) was 100.2mg/L (101.9mg/L R-BAC+DA, 99.5mg/L T-BAC). The proportion of participants with a comorbidity, defined as one or more of hypertension, diabetes, or cardiovascular disease, was 52.5% (46.7% R-BAC+DA, 55.1% T-BAC). All randomised participants, except one, received dexamethasone prior to randomisation. In addition, 48 of the total 99 participants also received remdesivir or tocilizumab in addition to dexamethasone within the first 7 days. The average duration of dexamethasone treatment prior to dornase alfa was 1.13+/- 0.79 days (Table S1). The last pre-dexamethasone CRP was also similar between groups with a mean of 125.0mg/L (128.1mg/L R-BAC+DA, 122.7mg/L T-BAC). The number of days between dexamethasone initiation and baseline was 1.2 days (0.7 days R-BAC+DA, 1.3 days T-BAC). There were imbalances noted at baseline between the groups in white blood cell count, neutrophil count, procalcitonin count and D-dimer (**Table 1**).

**Table 1.**
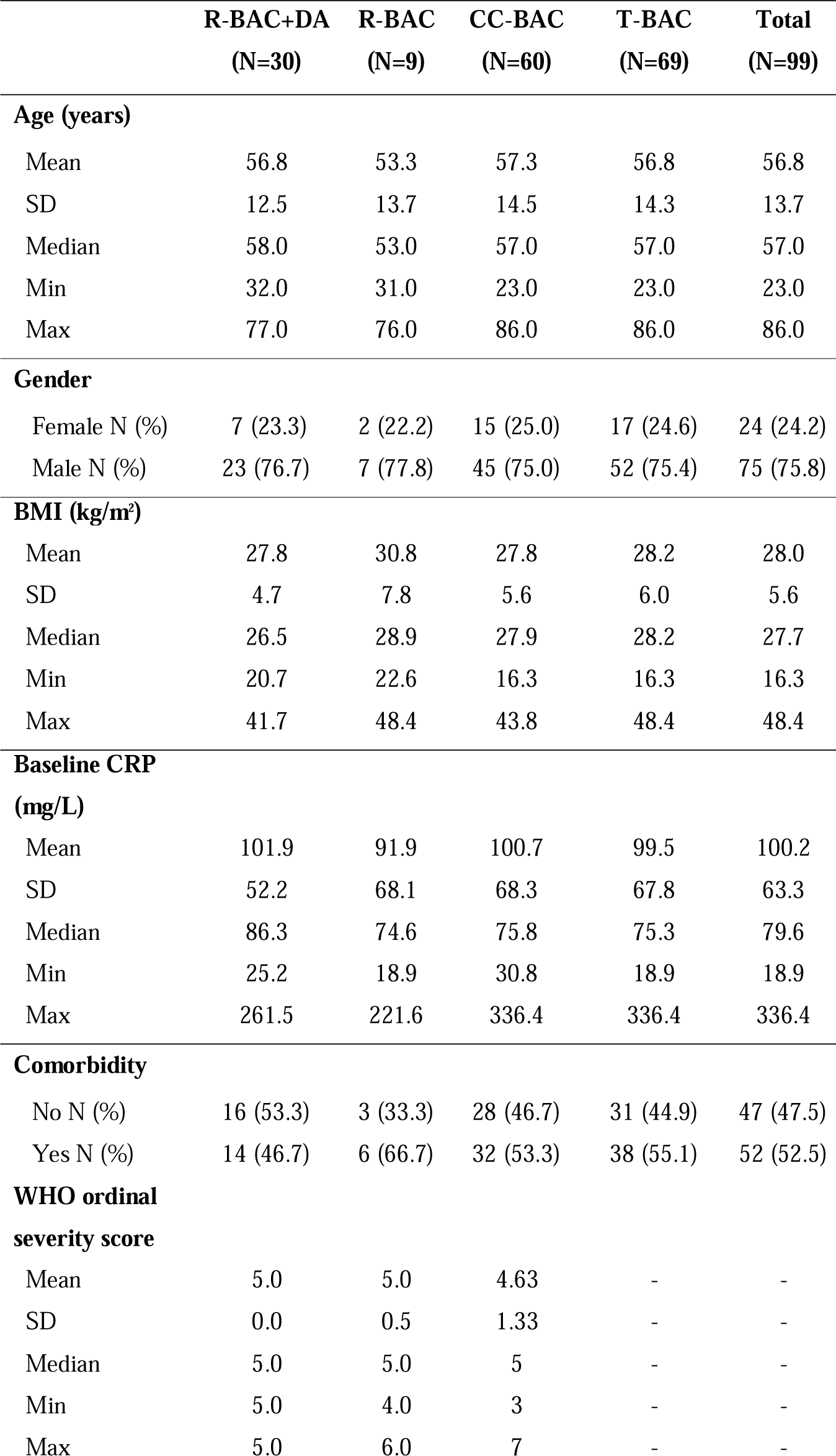
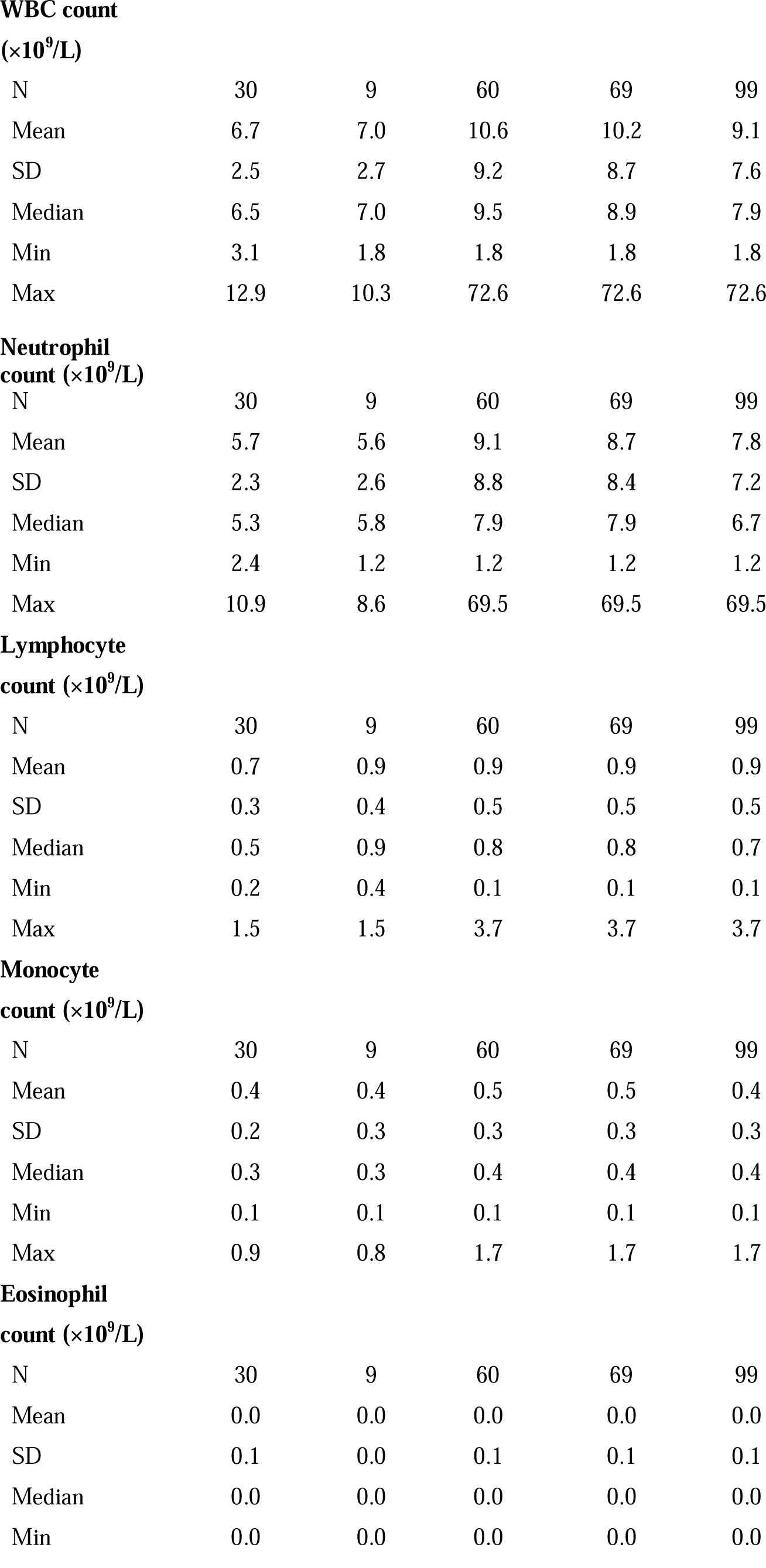
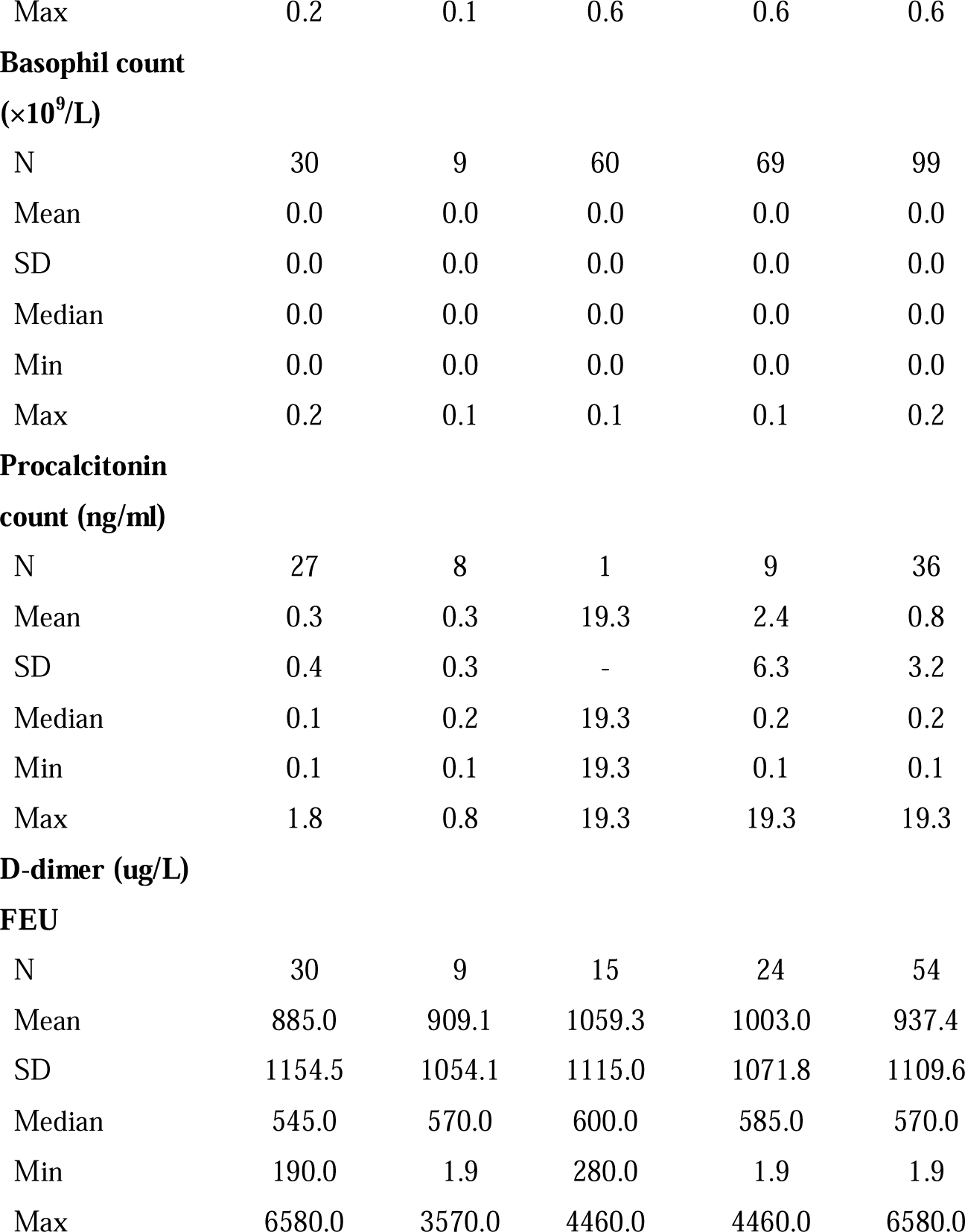
Patient baseline characteristics.

|  | **R-BAC+DA**<br>**(N=30)** | **R-BAC**<br>**(N=9)** | **CC-BAC**<br>**(N=60)** | **T-BAC**<br>**(N=69)** | **Total**<br>**(N=99)** |
| --- | --- | --- | --- | --- | --- |
| **Age (years)** |  |  |  |  |  |
| Mean | 56.8 | 53.3 | 57.3 | 56.8 | 56.8 |
| SD | 12.5 | 13.7 | 14.5 | 14.3 | 13.7 |
| Median | 58.0 | 53.0 | 57.0 | 57.0 | 57.0 |
| Min | 32.0 | 31.0 | 23.0 | 23.0 | 23.0 |
| Max | 77.0 | 76.0 | 86.0 | 86.0 | 86.0 |
| **Gender** |  |  |  |  |  |
| Female N (%) | 7 (23.3) | 2 (22.2) | 15 (25.0) | 17 (24.6) | 24 (24.2) |
| Male N (%) | 23 (76.7) | 7 (77.8) | 45 (75.0) | 52 (75.4) | 75 (75.8) |
| **BMI (kg/m<sup>2</sup>)** |  |  |  |  |  |
| Mean | 27.8 | 30.8 | 27.8 | 28.2 | 28.0 |
| SD | 4.7 | 7.8 | 5.6 | 6.0 | 5.6 |
| Median | 26.5 | 28.9 | 27.9 | 28.2 | 27.7 |
| Min | 20.7 | 22.6 | 16.3 | 16.3 | 16.3 |
| Max | 41.7 | 48.4 | 43.8 | 48.4 | 48.4 |
| **Baseline CRP**<br>**(mg/L)** |  |  |  |  |  |
| Mean | 101.9 | 91.9 | 100.7 | 99.5 | 100.2 |
| SD | 52.2 | 68.1 | 68.3 | 67.8 | 63.3 |
| Median | 86.3 | 74.6 | 75.8 | 75.3 | 79.6 |
| Min | 25.2 | 18.9 | 30.8 | 18.9 | 18.9 |
| Max | 261.5 | 221.6 | 336.4 | 336.4 | 336.4 |
| **Comorbidity** |  |  |  |  |  |
| No N (%) | 16 (53.3) | 3 (33.3) | 28 (46.7) | 31 (44.9) | 47 (47.5) |
| Yes N (%) | 14 (46.7) | 6 (66.7) | 32 (53.3) | 38 (55.1) | 52 (52.5) |
| **WHO ordinal**<br>**severity score** |  |  |  |  |  |
| Mean | 5.0 | 5.0 | 4.63 | - | - |
| SD | 0.0 | 0.5 | 1.33 | - | - |
| Median | 5.0 | 5.0 | 5 | - | - |
| Min | 5.0 | 4.0 | 3 | - | - |
| Max | 5.0 | 6.0 | 7 | - | - |
| **WBC count****( $\times 10^9/L$ )** |  |  |  |  |  |
| N | 30 | 9 | 60 | 69 | 99 |
| Mean | 6.7 | 7.0 | 10.6 | 10.2 | 9.1 |
| SD | 2.5 | 2.7 | 9.2 | 8.7 | 7.6 |
| Median | 6.5 | 7.0 | 9.5 | 8.9 | 7.9 |
| Min | 3.1 | 1.8 | 1.8 | 1.8 | 1.8 |
| Max | 12.9 | 10.3 | 72.6 | 72.6 | 72.6 |
| **Neutrophil count ( $\times 10^9/L$ )** |  |  |  |  |  |
| N | 30 | 9 | 60 | 69 | 99 |
| Mean | 5.7 | 5.6 | 9.1 | 8.7 | 7.8 |
| SD | 2.3 | 2.6 | 8.8 | 8.4 | 7.2 |
| Median | 5.3 | 5.8 | 7.9 | 7.9 | 6.7 |
| Min | 2.4 | 1.2 | 1.2 | 1.2 | 1.2 |
| Max | 10.9 | 8.6 | 69.5 | 69.5 | 69.5 |
| **Lymphocyte count ( $\times 10^9/L$ )** |  |  |  |  |  |
| N | 30 | 9 | 60 | 69 | 99 |
| Mean | 0.7 | 0.9 | 0.9 | 0.9 | 0.9 |
| SD | 0.3 | 0.4 | 0.5 | 0.5 | 0.5 |
| Median | 0.5 | 0.9 | 0.8 | 0.8 | 0.7 |
| Min | 0.2 | 0.4 | 0.1 | 0.1 | 0.1 |
| Max | 1.5 | 1.5 | 3.7 | 3.7 | 3.7 |
| **Monocyte count ( $\times 10^9/L$ )** |  |  |  |  |  |
| N | 30 | 9 | 60 | 69 | 99 |
| Mean | 0.4 | 0.4 | 0.5 | 0.5 | 0.4 |
| SD | 0.2 | 0.3 | 0.3 | 0.3 | 0.3 |
| Median | 0.3 | 0.3 | 0.4 | 0.4 | 0.4 |
| Min | 0.1 | 0.1 | 0.1 | 0.1 | 0.1 |
| Max | 0.9 | 0.8 | 1.7 | 1.7 | 1.7 |
| **Eosinophil count ( $\times 10^9/L$ )** |  |  |  |  |  |
| N | 30 | 9 | 60 | 69 | 99 |
| Mean | 0.0 | 0.0 | 0.0 | 0.0 | 0.0 |
| SD | 0.1 | 0.0 | 0.1 | 0.1 | 0.1 |
| Median | 0.0 | 0.0 | 0.0 | 0.0 | 0.0 |
| Min | 0.0 | 0.0 | 0.0 | 0.0 | 0.0 |
| Max | 0.2 | 0.1 | 0.6 | 0.6 | 0.6 |
| **Basophil count**<br>($\times 10^9/L$) | | | | | |
| N | 30 | 9 | 60 | 69 | 99 |
| Mean | 0.0 | 0.0 | 0.0 | 0.0 | 0.0 |
| SD | 0.0 | 0.0 | 0.0 | 0.0 | 0.0 |
| Median | 0.0 | 0.0 | 0.0 | 0.0 | 0.0 |
| Min | 0.0 | 0.0 | 0.0 | 0.0 | 0.0 |
| Max | 0.2 | 0.1 | 0.1 | 0.1 | 0.2 |
| **Procalcitonin**<br>**count (ng/ml)** |  |  |  |  |  |
| N | 27 | 8 | 1 | 9 | 36 |
| Mean | 0.3 | 0.3 | 19.3 | 2.4 | 0.8 |
| SD | 0.4 | 0.3 | - | 6.3 | 3.2 |
| Median | 0.1 | 0.2 | 19.3 | 0.2 | 0.2 |
| Min | 0.1 | 0.1 | 19.3 | 0.1 | 0.1 |
| Max | 1.8 | 0.8 | 19.3 | 19.3 | 19.3 |
| **D-dimer (ug/L)**<br>**FEU** |  |  |  |  |  |
| N | 30 | 9 | 15 | 24 | 54 |
| Mean | 885.0 | 909.1 | 1059.3 | 1003.0 | 937.4 |
| SD | 1154.5 | 1054.1 | 1115.0 | 1071.8 | 1109.6 |
| Median | 545.0 | 570.0 | 600.0 | 585.0 | 570.0 |
| Min | 190.0 | 1.9 | 280.0 | 1.9 | 1.9 |
| Max | 6580.0 | 3570.0 | 4460.0 | 4460.0 | 6580.0 |

### Clinical outcomes

#### Longitudinal CRP correlates with disease outcomes

To ascertain the clinical value of CRP as the COVASE trial’s primary endpoint, we first examined the correlation of CRP with mortality in a cohort of 63 hospitalized patients with severe COVID-19 (Aramburu et al., 2022; Kurth et al., 2020; Messner et al., 2020). All participants who reached a WHO severity ordinance scale 7 were recruited at the Charité Hospital in Berlin in spring 2020 and did not receive anti-inflammatory therapies such as dexamethasone or anti-IL-6 antibody treatments. Plasma CRP measurements in 465 samples were segregated by disease outcome. This analysis revealed that deceased participants had significantly higher CRP than participants in the survivor group (**Figure 2A**). Clustering the 63 participants according to their longitudinal average CRP indicated that the frequency of mortality increased as the average CRP per patient increased (**Figure 2B**). Consistently, Mantel Cox survival analysis of the participants segregated into three groups with average longitudinal CRP ranges of 0-100 mg/L, 100-200 mg/L and 200-450 mg/L, showed that survival decreased significantly as the average CRP increased (**Figure 2C**). These results highlighted the strong correlation between CRP and survival in hospitalized individuals with severe COVID-19 pneumonia, suggesting that the blood CRP concentration was a strong predictor of clinical outcomes such as the probability of survival.

**Figure 2.**
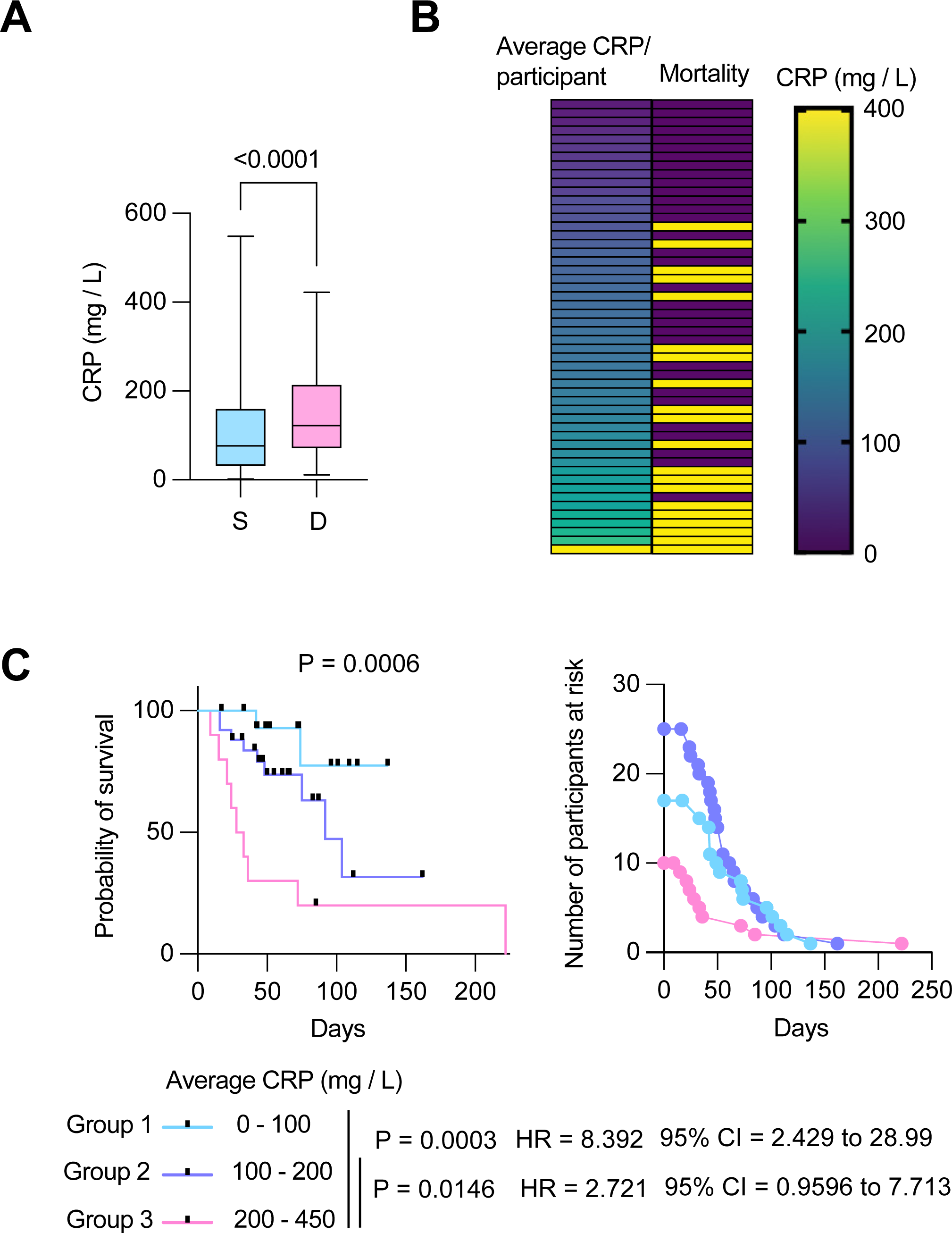
Longitudinal CRP predicts survival probability in severe COVID-19 pneumonia. A. Individual CRP concentrations in 465 plasmas from 63 participants with maximum WHO severity grade 7 COVID-19 pneumonia segregated into survivor and deceased groups, extracted from the Berlin COVID-19 study. B. Participants ordered by their longitudinal average CRP concentrations shown in the left column. Mortality is depicted in yellow in the right column. C. Kaplan Meier survival probabilities (left panel) and numbers at risk (right panel) for patients segregated into three categories of longitudinal average CRP ranges: 0-100, 100-200, and 200-450 mg / L. Statistical significance (P), Hazard ratios (HR) and 95% confidence intervals (95% CI) for group 1 against group 3 and group 2 against group 3 are shown below the survival plot. Statistics by Mann-Whitney and Mantel-Cox log rank tests.

#### Primary outcome

Next, we analysed the results from our COVASE participants. The individual CRP traces over time are shown in **Figure S3A**. Blood collection for both R-BAC and R-BAC+DA groups occurred at similar times and comparable frequencies (**Figure S3B**). In the participants who were randomised into the COVASE trial, excluding the CC-BAC participants the LS mean Ln (CRP) over 7 days follow-up was 3.10 (95% confidence interval [CI] 2.84 to 3.35) R-BAC+DA (n=30), and 3.59 (95% CI, 3.13 to 4.06) in R-BAC (n=9) (Table 2 and **Figure 3A**). Moreover, in the ITT group which included randomised and CC-BAC groups, the LS mean Ln (CRP) over 7 days follow-up was 3.15 (95% confidence interval [CI] 2.87 to 3.42) R-BAC+DA (n=30), and 3.55 (95% CI, 3.35 to 3.75) in T-BAC (n=69) (Table 2 and **Figure 3A**), p=0.01. This indicates a reduction in mean CRP of approximately 33% in the R-BAC+DA (23.23 mg/mL) compared to T-BAC (34.82 mg/mL) at mean follow-up over 7 days. The suppressive effect of dornase alfa on CRP was confirmed in various other subgroup analyses: the per-modified-protocol population only; participants who were randomised to BAC+DA in the COVASE trial, and CC-BAC participants alone excluding those randomised to BAC (**Table 2**). In addition, to ensure that the CC-BAC participants did not have a significantly different CRP trajectory to those randomised to BAC, we compared participants who were randomised to BAC (R-BAC) with the CC-BAC group by excluding those randomised to R-BAC+DA and found no significant differences (**Table 2**). Sensitivity analyses supported the observed effect on CRP. These included the Ln(CRP) as an area under the curve; the CC-BAC matched for the last pre-dexamethasone CRP measurement as opposed to their first CRP after starting dexamethasone; and the effect of remdesivir or tocilizumab (**Table 2**). A daily mean Ln CRP measurement chart is also provided (**Table 3**).

**Table 2.** Primary endpoint and sensitivity analysis.

| CRP (mg/L) | BAC+DA | BAC | Difference | p-value* |
| --- | --- | --- | --- | --- |
| **ITT population (R-BAC+ DA, R-BAC, CC-BAC)** |  |  |  |  |
| N | 30 | 69 |  |  |
| LS means log(CRP)* | 3.15 | 3.55 | -0.4 | 0.010 |
| (95% CI) | (2.87 to 3.42) | (3.35 to 3.75) | (-0.71 to -0.10) |  |
| LS means CRP ** | 23.23 | 34.82 | 0.67 |  |
| (95% CI) | (17.71 to 30.46) | (28.55 to 42.47) | (0.49 to 0.91) |  |
| **Sensitivity Analyses** |  |  |  |  |
| **PP population (R-BAC+DA, R-BAC, CC-BAC)** |  |  |  |  |
| N | 29 | 68 |  |  |
| LS means log(CRP)* | 3.12 | 3.55 | -0.43 | 0.006 |
| (95% CI) | (2.85 to 3.39) | (3.36 to 3.74) | (-0.73 to -0.13) |  |
| LS means CRP** | 22.64 | 34.82 | 0.65 |  |
| (95% CI) | (17.35 to 29.54) | (27.7 to 42.21) | (0.48 to 0.88) |  |
| **ITT population (R-BAC+DA, R-BAC)** |  |  |  |  |
| N | 30 | 9 |  |  |
| LS means log(CRP)* | 3.1 | 3.59 | -0.5 | 0.041 |
| (95% CI) | (2.84 to 3.35) | (3.13 to 4.06) | (-0.97 to -0.02) |  |
| LS means CRP** | 22.12 | 36.34 | 0.61 |  |
| (95% CI) | (17.16 to 28.5) | (22.79 to 57.94) | (0.38 to 0.98) |  |
| **ITT population (R-BAC+DA, CC-BAC)** |  |  |  |  |
| N | 30 | 60 |  |  |
| LS means log(CRP)* | 3.18 | 3.56 | -0.37 | 0.019 |
| (95% CI) | (2.91 to 3.45) | (3.35 to 3.76) | (-0.68 to -0.06) |  |
| LS means CRP** | 24.09 | 35.03 | 0.69 |  |
| (95% CI) | (18.36 to 31.6) | (28.44 to 43.15) | (0.5 to 0.94) |  |
| **ITT population (R-BAC, CC-BAC)** |  |  |  |  |
| N | 9 | 60 |  |  |
| LS means log(CRP)* | 3.78 | 3.53 | 0.24 | 0.386 |
| (95% CI) | (3.23 to 4.33) | (3.3 to 3.77) | (-0.32 to 0.8) |  |
| LS means CRP** | 43.69 | 34.23 | 1.28 |  |
| (95% CI) | (25.18 to 75.8) | (27.11 to 43.24) | (0.73 to 2.23) |  |
| **Area under the log(CRP), standardised by days followed up, over 7 days follow-up** |  |  |  |  |
| **ITT population (R-BAC+DA, R-BAC, CC-BAC)** |  |  |  |  |
| N | 30 | 69 |  |  |
| LS means area<sup>a</sup> | 3.45 | 3.72 | -0.27 | 0.043 |
| (95% CI) | (3.22 to 3.68) | (3.55 to 3.88) | (-0.53 to -0.01) |  |
| **ITT population (R-BAC+DA, R-BAC, CC-BAC) including last pre-Dexamethasone CRP value** |  |  |  |  |
| N | 30 | 69 |  |  |
| LS means log(CRP)* | 3.16 | 3.69 | -0.53 | 0.007 |
| (95% CI) | (2.83 to 3.5) | (3.44 to 3.93) | (-0.91 to -0.14) |  |
| LS means CRP** | 23.57 | 39.92 | 0.59 |  |
| (95% CI) | (16.85 to 32.97) | (31.32 to 50.89) | (0.4 to 0.87) |  |
| **ITT population (R-BAC+DA, R-BAC, CC-BAC) stratified by BAC treatment** |  |  |  |  |
| **No Remdesivir or Tocilizumab** |  |  |  |  |
| N | 12 | 39 |  |  |
| LS means log(CRP)* | 3.29 | 3.75 | -0.45 | 0.079 |
| (95% CI) | (2.83 to 3.76) | (3.45 to 4.04) | (-0.96 to 0.05) |  |
| LS means CRP** | 26.97 | 42.35 | 0.64 |  |
| (95% CI) | (16.87 to 43.11) | (31.44 to 57.04) | (0.38 to 1.06) |  |
| **Remdesivir no Tocilizumab** |  |  |  |  |
| N | 16 | 23 |  |  |
| LS means log(CRP)* | 3.16 | 3.5 | -0.35 | 0.123 |
| (95% CI) | (2.79 to 3.53) | (3.18 to 3.83) | (-0.79 to 0.1) |  |
| LS means CRP** | 23.53 | 33.26 | 0.71 |  |
| (95% CI) | (16.29 to 33.99) | (23.97 to 46.15) | (0.45 to 1.1) |  |
| **Tocilizumab no Remdesivir** |  |  |  |  |
| N | 1 | 5 |  |  |
| **Remdesivir and Tocilizumab** |  |  |  |  |
| N | 1 | 2 |  |  |
| * From linear repeated measures model, adjusted for natural log(baseline CRP, age, sex, BMI, serious condition, time, treatment, a treatment*time interaction, and subject as a random effect. Least squares means compared at mean follow-up time. |  |  |  |  |
| ** Antilog of estimates from *. Ratio of BAC + dorna-alfa: BAC shown in the difference column. |  |  |  |  |
| <sup>a</sup> From linear model, adjusted for natural log(baseline CRP, age, sex, BMI, serious condition, and treatment. |  |  |  |  |

**Figure 3.**
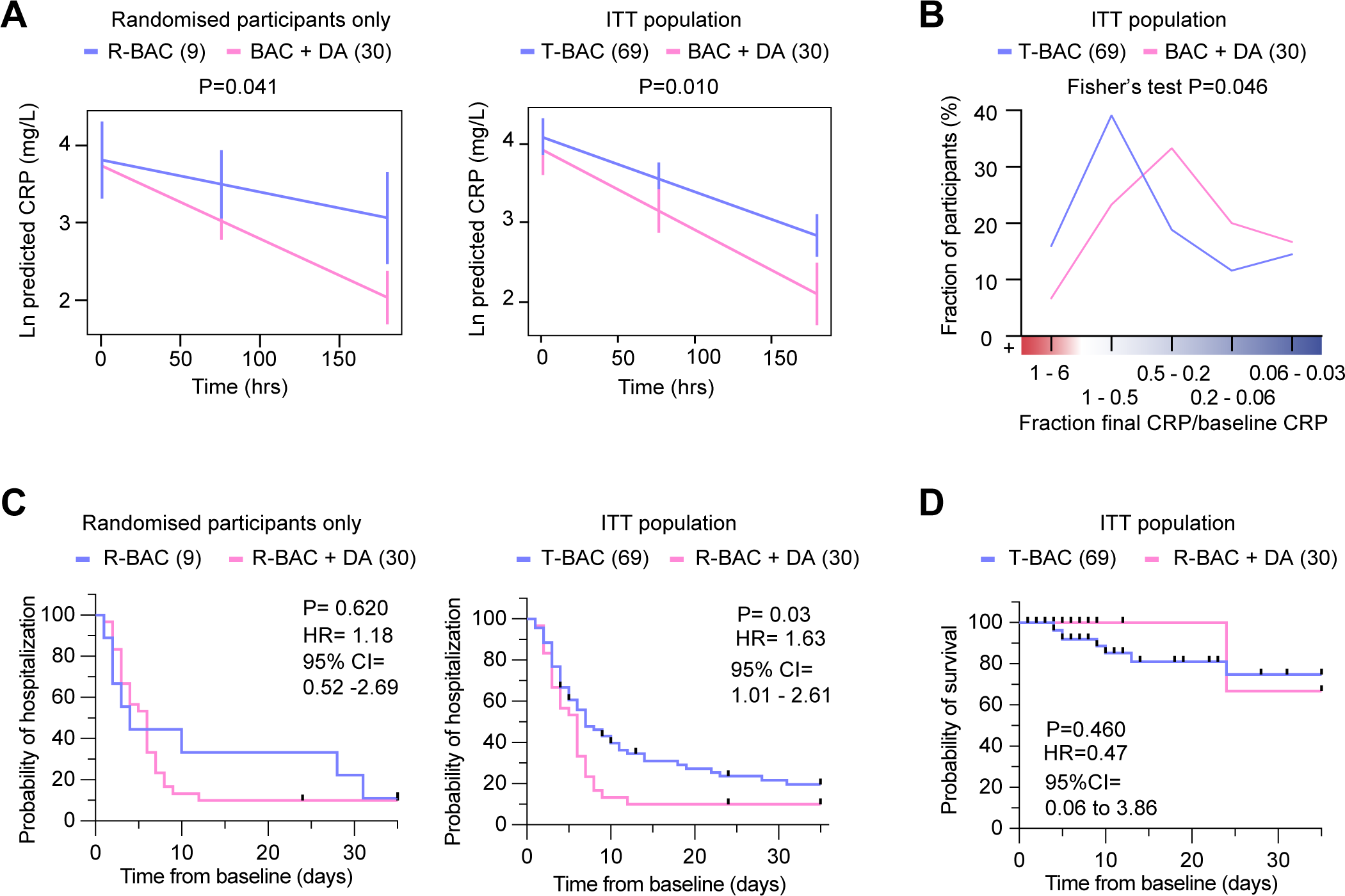
Analysis of primary and clinical endpoints. A. Fitted mean (95% confidence interval) from mixed model of natural log (CRP) over 7 days follow-up as the outcome. (Left panel) randomised participants only: Blue: participants randomised to R-BAC, N=9; Pink: participants randomised to R-BAC+DA, N=30. (Right panel) ITT population: Blue: T-BAC (CC-BAC and R-BAC) N=69; Pink: R-BAC+DA, N=30. Results were adjusted for natural log baseline CRP, age, sex, BMI, serious comorbidity (Diabetes, Cardiovascular disease or hypertension), time and a treatment × time interaction. P-value generated by comparing least-square means between arms. B. Distribution of participants based on the change in CRP measured as a ratio of the final CRP reading within the 7-day treatment period over the baseline CRP reading per patient. Statistical analysis by Fisher’s test. C. Kaplan-Meier plot showing time to discharge from hospital from baseline. Hazard ratio from Cox proportional hazards model adjusted for baseline CRP, age, sex, BMI, serious comorbidity (diabetes, cardiovascular disease of hypertension). P-value from log-rank test. (Blue: CC-BAC and participants randomised to R-BAC, N=69. Pink: participants randomised to R-BAC+DA, N=30). D. Kaplan-Meier plot showing time to death over 35 days follow up. Hazard ratio from Cox proportional hazards model adjusted for baseline CRP, age, sex, BMI, serious comorbidity (Diabetes, Cardiovascular disease of hypertension). P-value from log-rank test.

**Table 3.** Primary endpoint by day.

| **Days from**<br>**baseline** | **Mean**<br>**BAC +**<br>**DA** | **SD BAC**<br>**+ DA** | **N**<br>**BAC**<br>**+ DA** | **Mean**<br>**BAC** | **SD BAC** | **N**<br>**BAC** |
| --- | --- | --- | --- | --- | --- | --- |
| 1 | 4.058 | 0.591 | 21 | 4.029 | 0.734 | 42 |
| 2 | 3.538 | 0.658 | 33 | 3.852 | 0.937 | 52 |
| 3 | 3.332 | 0.816 | 24 | 3.839 | 1.037 | 53 |
| 4 | 2.917 | 1.017 | 17 | 3.573 | 1.132 | 37 |
| 5 | 2.52 | 1.118 | 15 | 3.872 | 1.157 | 24 |
| 6 | 3.059 | 1.508 | 12 | 3.547 | 1.516 | 26 |
| 7 | 3.386 | 1.824 | 9 | 3.504 | 1.451 | 27 |
| 8 | 3.428 | - | 1 | 2.478 | 1.393 | 9 |

To better understand the effects of dornase alfa on the CRP of the treated population we also performed a frequency distribution analysis of the change in CRP over time. Instead of fitting a slope on the CRP values of the individual samples of each participant, we plotted the fraction between the CRP measurement of the baseline and final sample collected for each participant during the 7 day treatment period. We then plotted the distribution of participants within the different ranges of change in CRP (**Figure 3B**). Compared to T-BAC, the R-BAC+DA treatment group exhibited a higher CRP reduction distribution with fewer participants that did not respond to treatment as indicated by an increase in CRP concentrations. Therefore, the addition of dornase alfa to dexamethasone resulted in a significant and sustained reduction in CRP in a greater number of COVID-19-infected participants compared to treatment with dexamethasone alone.

#### Secondary outcomes

The length of hospitalisation was analysed as a time-to-event outcome of alive discharge from the hospital censored at 35 days. The hazard ratio observed in the Cox proportional hazards model was 1.63 (95% CI, 1.01 to 2.61), p=0.03 (**Figure 3C and S3C**). Showing that throughout 35 days follow-up, there was a 63% higher chance of discharge alive at any given time-point in R-BAC+DA compared to T-BAC. Although the rate of discharge was similar in 50% of patients, 80% discharge occurred by 8 days in R-BAC+DA whereas, whereas the same proportion was reached at 30 days in T-BAC. This trend was also seen when only the randomised R-BAC participants were considered, although not powered to reach significance and with a smaller HR of 1.18 (95% CI, 0.52-2.69), p=0.62, (**Table S2 and Figure S3C**).

Over 7 days of follow up there was no significant difference between R-BAC+DA and T-BAC in either the fraction of participants admitted to ICU (23.3% versus 21.74%), p=0.866, or the length of ICU stay, LS mean 21.25 (95% CI, 4.65 to 37.84) hours versus 19.85 (95% CI, 8.00 to 31.70) hours, p=0.883. The same was seen over 35-day follow-up, LS mean 55.21 95% CI, -23.59 to 134.00) hours versus 60.60 (95% CI, 4.34 to 116.86) hours, p=0.905. At any point during the 35 days follow-up, 23% of R-BAC+DA were admitted to ICU compared to 23.19% T-BAC, p= 0.983 (**Table S3**). Similarly, there was no significant difference in the time requiring oxygen between the two groups (R-BAC-DA vs T BAC), at either 7 days, LS mean 94.32 (95% CI, 72.8 to 115.79) hours, versus 88.96 (95% CI, 73.64 to 104.29) hours, p=0.662, or 35 days, LS mean 133.22 (95% CI, 52.01 to 214.43) hours versus 156.35 (95% CI, 98.36, 214.33) hours, p=0.618. At 35 days follow up, there were only 9 randomised participants to evaluate, but mean oxygen use exhibited a reduction of 123 hours in R-BAC+DA participants, versus 241 hours for the T-BAC group, p=0.187 (**Table S3**).

The time to event data was censored at 28 days post last dose (up to d35) for the randomised participants (R-BAC and R-BAC+DA) and at the date of the last electronic record for the CC-BAC group. Over 35 days follow up, 1 person amongst the 30 patients in the R-BAC+DA group died, compared to 8 of the 69 T-BAC participants. The hazard ratio observed in the Cox proportional hazards model was 0.47 (95% CI, 0.06 to 3.86), indicating a trend towards a reduced chance of death at any given time-point in R-BAC+DA compared to T-BAC, but this did not reach significance p= 0.460 (**Figure 3D and S3C**). The hazard ratio observed in the Cox proportional hazards model (95% CI) was 0.47 (0.06, 3.86), which estimates that throughout 35 days follow-up, there was a 53% reduced chance of death at any given timepoint in the R-BAC+DA group compared to the T-BAC group. However, the confidence intervals are wide due to the small number of events and consequently the p-value from a log-rank test was 0.460, which does not reach statistical significance.

There was no significant difference at either 7- or 35-days follow-up, in the number of participants that required mechanical ventilation in R-BAC+DA compared with T-BAC (16.67% vs 13.04%), p=0.628. Amongst participants that were ventilated, the mean length of mechanical ventilation at 7 days follow-up in R-BAC+DA particpants was 76.8 hours, compared to 88.78 in the T-BAC group. At 35 days follow-up, the mean length of mechanical ventilation in BAC+DA was 76.8 hours compared to 411.17 hours in T-BAC participants (**Table S3**). There was no significant difference in superadded bacterial pneumonia at either 7 or 35 days follow-up: 7 days, 1 (3.33%) participant in BAC+DA compared to 3 (4.35%) participants in T-BAC, p= 0.934; 35 days, 2 (6.67%) R-BAC+DA participants had bacterial pneumonia, compared to 3 (4.35%) T-BAC participants, p=0.548 (**Table S3**).

Blood analysis with no adjustment for multiple testing showed a significant treatment effect in the BAC+DA vs. T-BAC group comparison for three parameters: lymphocyte counts, D-dimer, and procalcitonin (PCT). First, R-BAC+DA exhibited higher lymphocyte counts with LS mean of 0.87 (95% CI, 0.76-0.98) in the T-BAC group vs. 1.08 (95% CI, 0.92-1.27) in R-BAC+DA participants, p=0.02 (**(Figure 4A, Table S2**). Amongst individuals with lymphopenia at baseline (<1×10^9^ lymphocytes/L), the R-BAC+DA group exhibited a greater increase in blood lymphocyte numbers compared to the T-BAC group throughout the entire length of treatment. Furthermore, D-dimer levels were lower in R-BAC+DA participants compared to T-BAC participants, with LS mean D-dimer difference of 1657 (95% CI, 3131-877) (**(Figure 4B and S4A**). R-BAC+DA participants also exhibited lower levels of PCT, mean 0.18 ng/mL (95% CI, -0.2-0.56) compared to the T-BAC group, mean 1.31 ng/mL (95% CI, 0.56-2.05), p=0.005. Repeat analysis excluding the CC-BAC population replicated these results, and changes in all 3 parameters were significant (**Table S2**).

**Figure 4.**
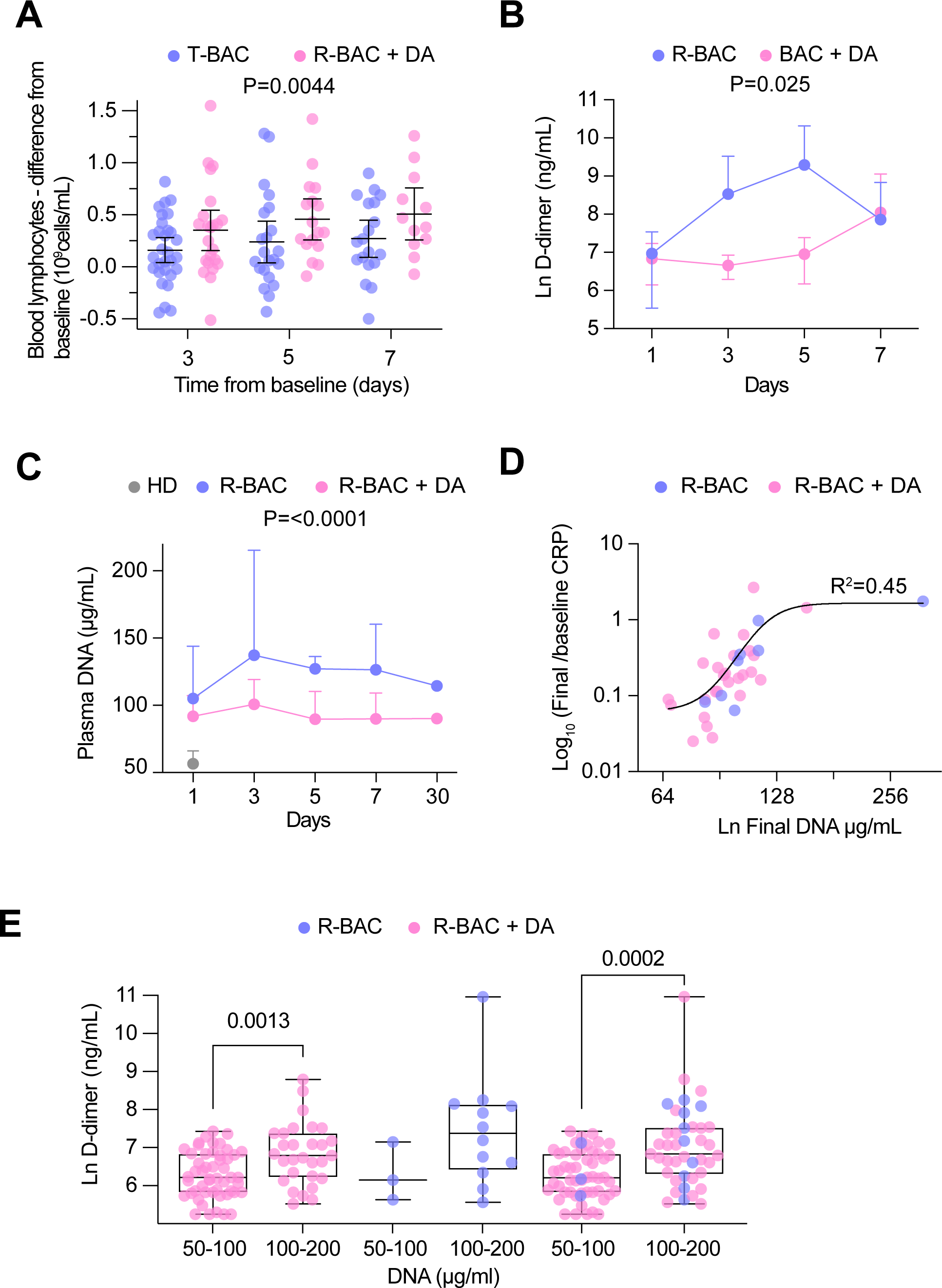
Analysis of secondary and exploratory endpoints in blood. A. Difference between the lymphocyte count for each day of the treatment period and the baseline in each ITT participant who exhibited lymphopenia at baseline (<1×10^9^ lymphocytes/mL). Mean and 95%CI interval is shown with statistical analysis by two-way Anova. B. Mean D-dimer levels per day in randomised R-BAC (blue) and R-BAC+DA (pink) participants with error bars depicting 95% CI. Statistical difference by mixed effects Anova analysis. C. Mean cf-DNA levels per day in randomised R-BAC (blue) and R-BAC+DA (pink) participants, with error bars depicting standard deviation. Statistical analysis by mixed effects Anova. D. Correlation between the final cf-DNA levels and ratio of CRP at day-7 normalized to the baseline CRP (CRP_final_/CRP_baseline_) per randomised participant. Fitting by non-linear regression. E. Correlation between D-dimer and cf-DNA levels in the blood of participants randomised to R-BAC (blue) or to R-BAC+DA (DA) (pink), where samples were segregated depending on whether the corresponding levels of cf-DNA were below or above 100 μg/mL. Statistical analysis by unpaired parametric t-test.

#### Exploratory outcomes

Given the potential role of circulating cf-DNA in pathology, we examined whether the pulmonary administration of dornase alfa influenced cf-DNA levels circulating in plasma. Compared to anonymized healthy donor volunteers at the Francis Crick institute (HD), cf-DNA levels were elevated in the plasmas of all R-BAC and R-BAC-DA randomised COVASE participants. There was no difference in baseline plasma cf-DNA levels between the two randomised participant groups. Notably, during the treatment period plasma cf-DNA was reduced in R-BAC+DA compared to R-BAC participants (**(Figure 4C and Figure S4B**). Moreover, there was a positive correlation between the levels of cf-DNA in the final sample collected during the treatment period and the magnitude of CRP reduction measured as the ratio of final over baseline CRP (CRP_final_/CRP_baseline_) (**(Figure 4D)**. Moreover, samples with cf-DNA >100μg/mL exhibited significantly higher D-dimer levels compared to samples with cf-DNA levels <100μg/mL ((**Figure 4E**). Hence, dornase alfa treatment was associated with a suppression of circulating cf-DNA that correlated with a reduced risk for coagulopathy and a more sustained reduction in CRP.

#### Safety

Dornase alfa was well tolerated with no systemic side-effects which is consistent with its short half-life in vivo (**Table S4**). Overall, there were 10 reported adverse events (AEs) in the study, in 9 R-BAC and 30 R-BAC+DA participants **(Table S5)**. Of these, one was reported by the clinical team as definitely related and one as unlikely to be related to dornase alfa. No treatment-related serious AEs were reported.

## DISCUSSION

Our findings show that nebulised dornase alfa significantly reduced systemic inflammation as measured by plasma CRP concentrations in patients with severe COVID-19 pneumonia even when already receiving dexamethasone. Dornase alfa exhibited a robust anti-inflammatory effect as assessed with several sensitivity analyses including analysis of only randomised participants, excluding CC-BAC participants. These findings confirm the usefulness of CRP as a sensitive readout to monitor the impact of anti-inflammatory agents in small cohorts. Improvements in care were also indicated by other biomarkers and secondary endpoints when both randomised and CC-BAC participants were considered. Moreover, despite not being formally powered for length of stay, dornase alfa reduced time to discharge over 35 days.

During the pandemic, the identification of novel and repurposed treatments effective for COVID-19 was hampered by patient recruitment to competing studies during pandemic. This resulted in small studies with inconclusive or contrary findings. Our study design took advantage of frequent repeated measures of CRP in each patient, to allow a smaller sample size to determine efficacy. CRP was also a primary readout in CATALYST (Fisher et al., 2022) and ATTRACT studies (Tornling et al., 2021). In addition, we used contemporary controls as additional comparators to use limited resources more efficiently.

Following the RECOVERY trial, dexamethasone became the standard care in patients with COVID-19 pneumonitis requiring oxygen. We recruited participants with CRP ≥ 30 mg/L, on the day after receiving dexamethasone to minimise steroid-dependent effects on CRP. The finding that dornase alfa can significantly reduce CRP in participants receiving dexamethasone suggests a complementary mechanism of action with significant improvement in existing standard anti-inflammatory care. Moreover, dornase alfa may be a treatment choice for patients with mild COVID-19 pneumonia not requiring oxygen, in whom dexamethasone may be harmful (Group et al., 2021).

The primary endpoint effect was consistently reflected in secondary outcomes. Dornase alfa increased the chance of live discharge by 63% at any time up to 35 days, reduce overall hospital stay. The reduction in hospital occupancy during the COVID-19 pandemic proved critical in sustaining life-saving treatment availability. In addition, dornase alfa significantly increased lymphocyte counts and reduced D-dimer. In addition to the increased mortality data we provide here, CRP levels are associated with venous thromboembolic disease in COVID-19 with the worst outcomes seen in patients with high CRP and D-dimers (Smilowitz et al., 2021). In a metanalysis of 32 studies involving 101491 COVID-19 patients, elevated CRP (OR 4.37, p<0.00001), lymphopenia (3.33, p<0.00001), elevated D-dimer (3.39 (p<0.00001) and elevated PCT (6.33, p<0.00001) were independent markers of poor outcomes (Malik et al., 2021).

Our study design offered a solution to the early screening of compounds for inclusion in larger platform trials. The study took advantage of frequent repeated measures of quantifiable CRP in each patient, to enable the determination of efficacy using a smaller sample size to than if powered were based on clinical outcomes. We applied a CRP-based approach that was similar to the CATALYST and ATTRACT studies (Fisher et al., 2022; Tornling et al., 2021). CATALYST showed in much smaller groups (usual care, 54, namilumab, 57 and infliximab, 35) that the GM-CSF-blocking antibody namilumab reduced CRP even in participants treated with dexamethasone whereas infliximab that targets TNF-α had no significant effect on CRP. This led to a suggestion that namilumab should be considered as an agent to be prioritised for further investigation in the RECOVERY trial. A direct comparison of our results with CATALYST is difficult due to the different nature of the modelling employed in the two studies. Nevertheless, dornase alfa exhibited comparable significance in the reduction in CRP compared to standard of care as described for namilumab at a fraction of the cost. Furthermore, endonuclease therapies may prove safer and more applicable than cytokine-blocking monotherapies, as they are unlikely to increase the risk for microbial co-infections which are a risk factor associated with antibody therapies that neutralize inflammatory cytokines that are critical for immune defence such as IL-1β, IL-6 or GM-CSF.

In addition, dornase alfa reduced circulating cf-DNA levels, suggesting that lung administration of the enzyme exerts systemic effects on circulating chromatin. The reduction in plasma cf-DNA levels suggests that by stripping the DNA from chromatin, dornase alfa suppresses the proinflammatory properties of histones and potentiates their degradation (Papayannopoulos et al., 2011; Tsourouktsoglou et al., 2020). The inverse correlation between final circulating cf-DNA and the reduction in CRP during treatment is consistent with link between circulating chromatin and systemic inflammation observed in animal models (Ioannou et al., 2022). The correlation between d-dimer and cf-DNA as well as the reduction in D-dimer after dornase alfa treatment is consistent with the reported pro-thrombotic role of NETs in the alveoli (Radermecker et al., 2020). Moreover, the improvement in the recovery rates of lymphopenia support the link between extracellular chromatin and lymphocyte death during sepsis that we also detected in individuals with severe COVID-19 pneumonia (Aramburu et al., 2022; Ioannou et al., 2022). Another reason why recombinant endonuclease treatment may be warranted in severe COVID-19 and microbial sepsis infections is the significant reduction in endogenous NET chromatin degradation capacity that strongly correlates with a high risk for mortality in these conditions (Aramburu et al., 2022).

Nebulized dornase alfa treatment has several advantages. Whilst immunisation has reduced COVID-19 hospital admissions, viral evolution and immune escape may still cause substantial mortality and other symports associated with long-COVID. Hence, there will always be a need for virally agnostic therapies that retain efficacy as viruses mutate. Importantly, nebulised dornase alfa has the potential to control immune pathology in a range of infections. Moreover, it can be administered safely and effectively in its nebulized form, outside the health-care setting. Three other small trials of dornase alfa (totalling 18 patients) in COVID-19 have reported improved oxygenation (Holliday et al., 2021; Okur et al., 2020; Weber et al., 2020). One small study indicated improvements in plasma and sputum proteomic profiles (Fisher et al., 2021). Despite differing study designs, patient populations and endpoints, there is a consensus for improvement in clinical outcomes.

Yet, there are a few limitations in this study. This single centre open-label study was designed to powerfully report futility or efficacy despite including just under 40 randomised patients and 60 CC-BAC. However, the trial was not powered to report mortality nor to overcome confounders, such as the use of antivirals and tocilizumab, an IL-6 inhibitor recognised to reduce CRP (Galvan-Roman et al., 2021), or the impact of an open-label study on influencing other therapeutic/ discharge decisions. Although underpowered, we demonstrate a trend to a reduction in CRP with dornase alfa in participants that had received tocilizumab and/or remdesivir, and those that had not. Moreover, the open-label nature of the study could potentially introduce a placebo effect bias. Nevertheless, we tried to minimize this by applying a standard testing schedule and discharge criteria.

In conclusion, we demonstrate that nebulised dornase alfa significantly reduces inflammation in hospitalised patients with severe COVID-19 pneumonia leading to improved clinical profiles and earlier discharge from hospital. These results confirm the role of cf-DNA in promoting systemic inflammation, coagulopathy and immune dysfunction in acute respiratory infections and suggest that recombinant endonucleases provide an additional mode of action that can broaden the effectiveness and sustainability of anti-inflammatory regimens. These encouraging data warrant further investigation in other serious respiratory infections characterized by hyperinflammation, cell lysis and NET formation.

## Supporting information

Supplemental figurs and tables, Consort checklist, SAP, Protocol

## Data Availability

All data produced in the present work are contained in the manuscript

## Acknowledgements

We thank the patients, caregivers, and families who participated in the trial; and acknowledge the help of the following: Additional BRC Contributors: Margaret Duku, Gulten Geneci, Farah Islam, Ciprian-Ionut Matei, Marta Merida, Eleni Nastouli, Marivic Ricamara, Anisa Tariq. Pharmacy: Matthew Baker, Nina Bason, Chi Yee Chung, Zoila Gilham-Fernandez, Temi Olusi. Sponsors/UCL: Liam Banks, Helen Cadiou, Novin Fard, Farhat Gilani, Vince Greaves, Yusuf Jaami, Pushpsen Joshi, Misha Ladva, David Lomas, Catherine Maidens, Anthea Mo, Anisha Nayar, Nick McNally, Samim Patel. Data Monitoring Committee: Balaji Ganeshan, Maria Leandro, Kay Roy. COVID Clinical Consultants: Diana Ayoola, Robin Bailey, David Brealey, Mike Brown, Anna Checkley, Charlie Coughlan, Philip Gothard, Robert Heyderman, Sarah Logan, Nicky Longley, Jessica Manson, Michael Marks, David Moore, Neil Stone, Emma Wall. T8 Nursing Staff: Adam Cureton-Griffiths, Amy Mann, Laura Nichols, Pantelis Savvides. NOCRI Respiratory Translational Research Collaboration: Chris Brightling, Jane Davies, Ratko Djukanovic, Liam Heeney, Ling-Pei Ho, Alex Horsley, Tracy Hussell, Stefan Marciniak, Lorcan McGarvey, Thomas Wilkinson. Pari/Roche Products Limited/LifeArc: Mal Apter, Ruth Davies, Ciara O’Brien, Pauline Stasiak, Davia Viellec.

## Author contributions

VP conceived the study. JCP, JI, ED, BW, AF, PTL, VB and VP contributed to the concept and design of the study. JCP, VJS, ED, RE, MIT, NDV, IVA, DH, DB, TC, JG, AW, HE, VJ, AL, TR, VP contributed to the acquisition of data. ETH, LL, FK designed and directed the Berlin Pa-COVID-19 study. JCP, JI, AF and VP contributed to analysis, and/or interpretation of data or the creation of new software used in the work. JCP, JI, PTL and VP drafted the manuscript. VP graphed and compiled figures and handled revisions. All authors have approved the submitted version (and any substantially modified version that involves the author’s contribution to the study); and to have agreed both to be personally accountable for the author’s own contributions and to ensure that questions related to the accuracy or integrity of any part of the work, even ones in which the author was not personally involved, are appropriately investigated, resolved, and the resolution documented in the literature. Roche Products Ltd conducted a factual accuracy check on the final version but any decision to incorporate comments was made solely at the discretion of the authors.

The authors declare no conflicts of interest.

