## Supplemental figurs and tables, Consort checklist, SAP, Protocol for "Anti-inflammatory therapy with nebulised dornase alfa for severe COVID-19 pneumonia"

Joanna C. Porter *et al.*

**Contents:**

I. Supplementary figures 1-4
II. Supplementary tables 1-4
III. Consort checklist
IV. SAP
V. Protocol

#### Supplementary Figure 1

CONSORT
TRANSPARENT REPORTING of TRIALS

##### CONSORT 2010 Flow Diagram

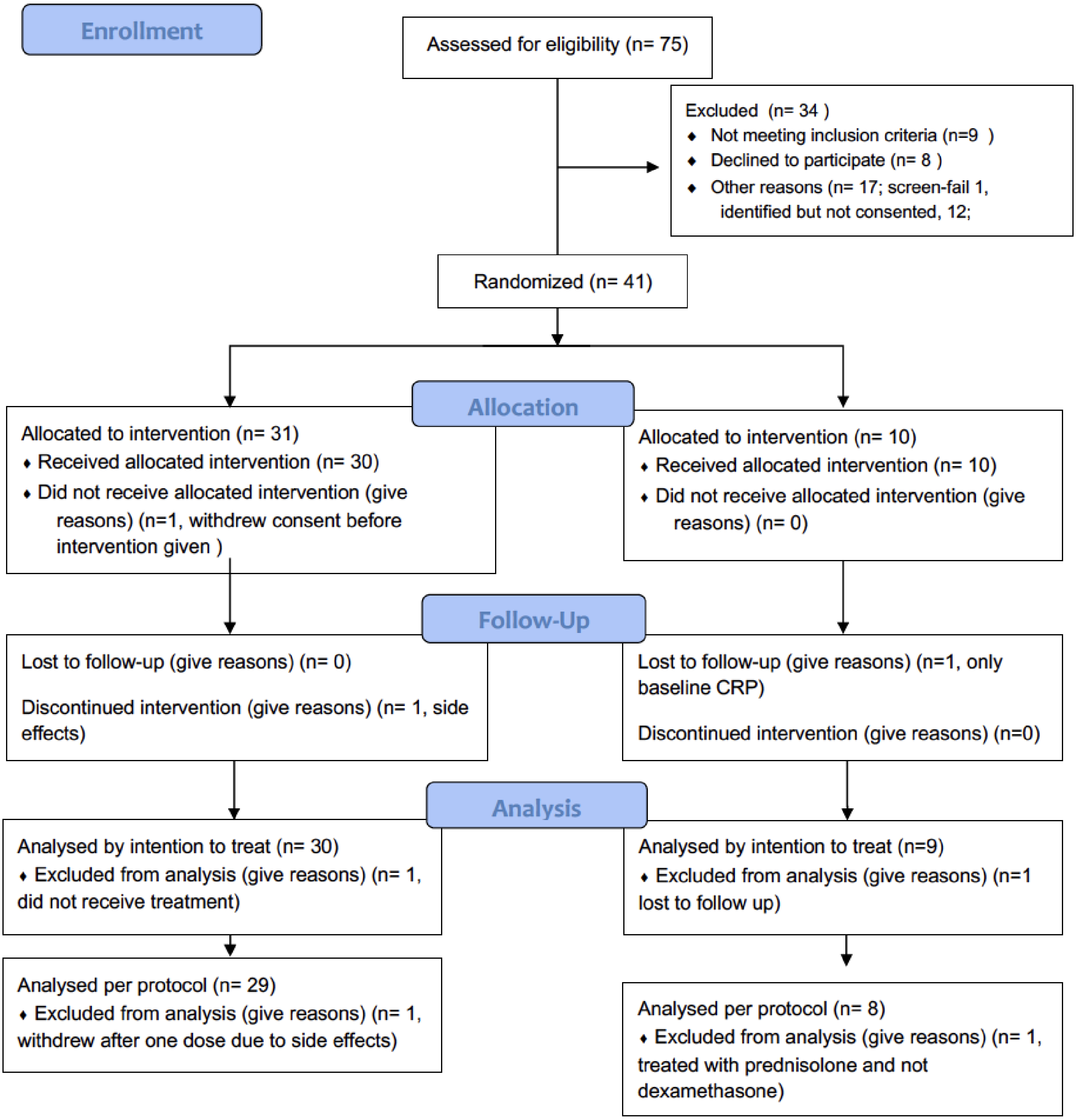

### Supplementary Figure 2

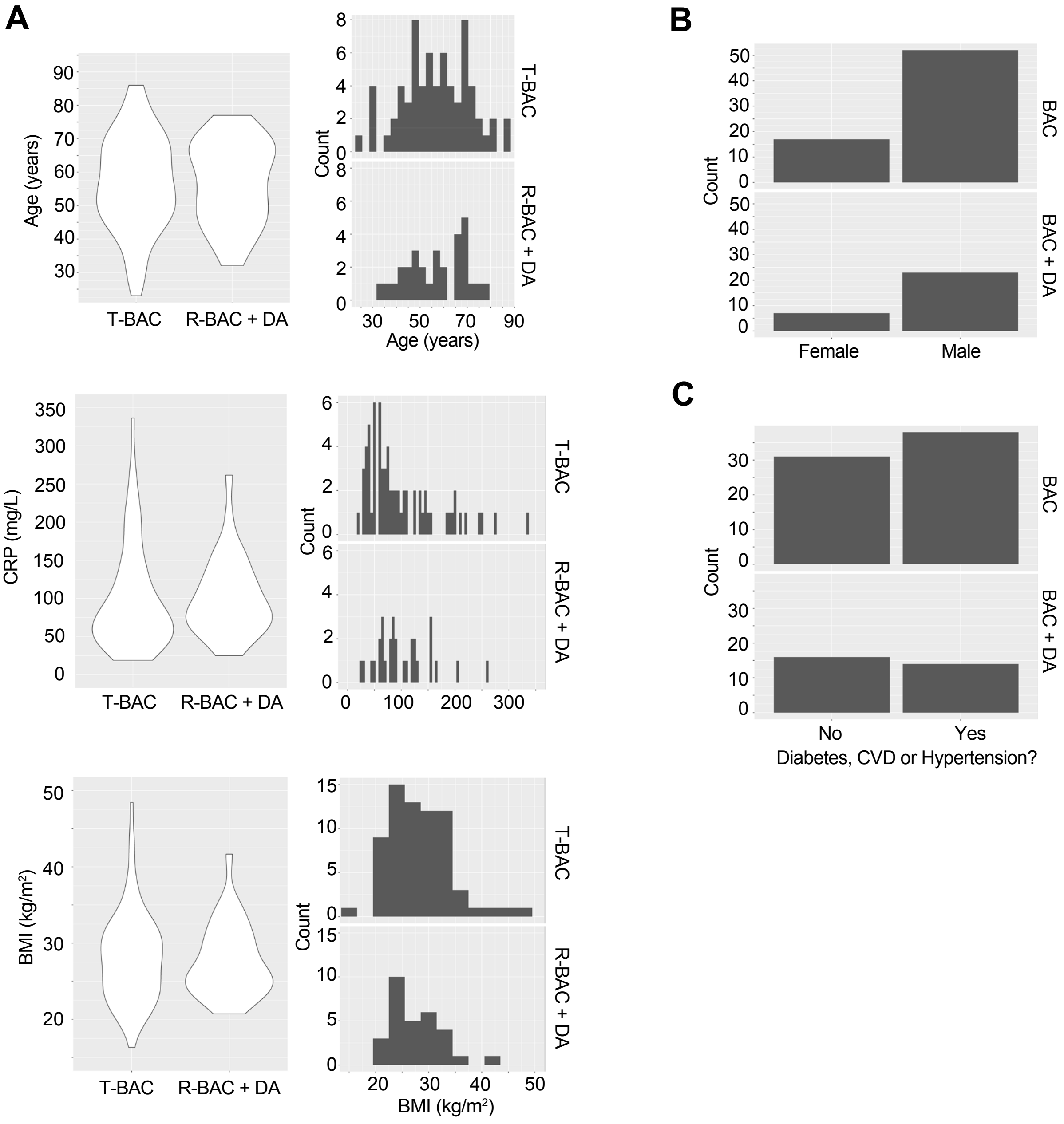

Supplementary Figure 2. Baseline characteristics of patients analysed in the trial.

(A). Violin plots (left) and frequency distribution (right) of baseline clinical parameters between participants in the contemporary control and randomised BAC group (T-BAC) and the randomised BAC+Dornase alfa (R-BAC+DA) group. (Top) Age, (middle) Baseline CRP and (bottom) Body mass index (BMI).
B. Number of male and female participants in the two groups.
C. Incidence of cardiovascular comorbidities in the two groups.

### Supplementary Figure 3

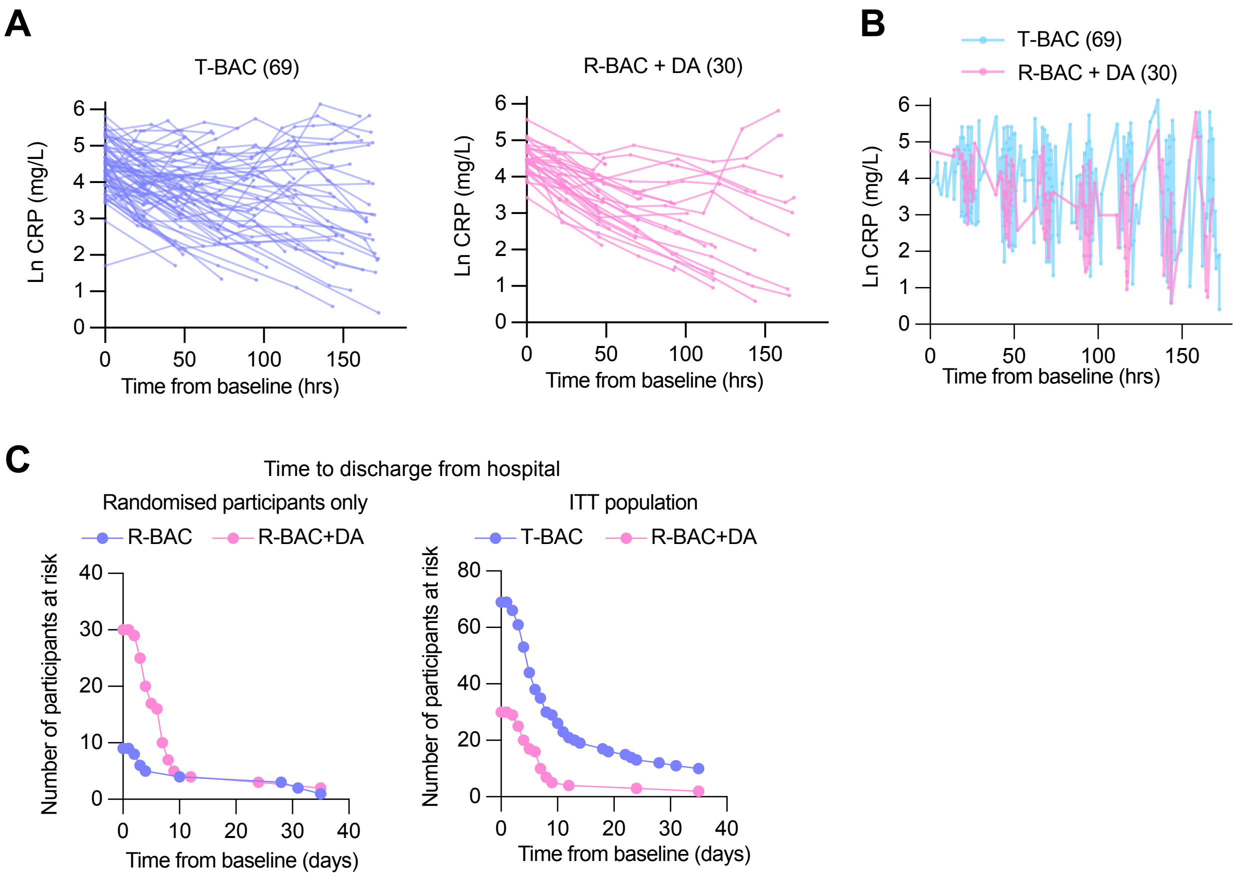

Supplementary Figure 3. Primary and clinical endpoints

A. (left panel) Natural log CRP in BAC (CC and randomised participants; blue). (Right panel) Natural log CRP in participants randomised to BAC+DA (pink).
B. Graph depicting the periodicity and frequency of blood sample collection for all post-baseline CRP values from contemporary control and randomised BAC (blue) or BAC+Dornase alfa (BAC+DA, pink) patients pooled into a single timeline.
C. Numbers at risk for Kaplan-Meier plots of the time to discharge from hospital (Figure 3C). ITT population (Blue: CC and participants randomised to BAC, N=69. Pink: participants randomised to BAC+DA, N=30).
D. Numbers at risk (right panel) depicting the time to discharge from hospital from baseline. Blue: participants randomised to R-BAC, N=9; Pink: participants randomised to R-BAC+DA, N=30. (Right panel) ITT population: Blue: T-BAC (CC-BAC and R-BAC) N=69; Pink: R-BAC+DA, N=30. Hazard ratio from Cox proportional hazards model adjusted for baseline CRP, age, sex, BMI, serious co-morbidity (Diabetes, Cardiovascular disease of hypertension).

### Supplementary Figure 4

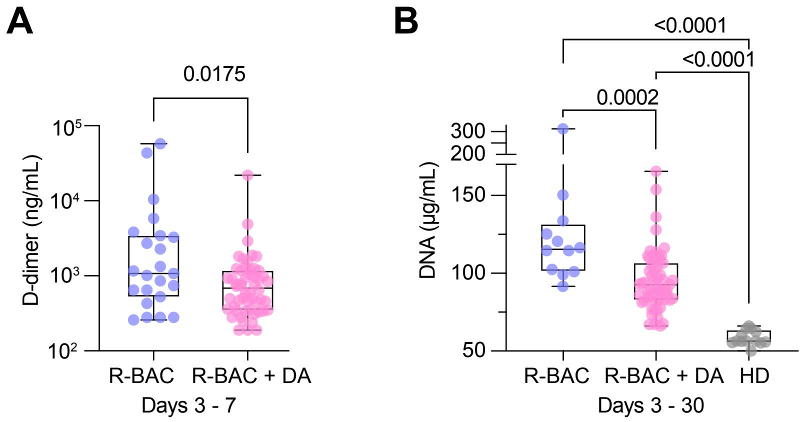

Supplementary Figure 4. Secondary and exploratory endpoints

A. D-dimer concentration in randomised participants post-baseline blood samples pooled into R-BAC and R-BAC + DA groups. Statistical analysis by two-tailed unpaired parametric t-test.
B. DNA concentration in randomised participants post-baseline blood samples pooled into R-BAC and R-BAC + DA groups. Statistical analysis by one-way Anova.

#### Supplementary Tables:

Supplementary Table 1: Duration of dexamethasone treatment prior to recruitment and initiation of Dornase alfa treatment

| | R-BAC+DA (N=30) | R-BAC (N=9) | CC-BAC (N=60) | All BAC (N=69) | Total (N=99) |
|---|---|---|---|---|---|
| **Length of Dexamethasone at baseline (days)** | | | | | |
| N | 30 | 8 | 60 | 68 | 98 |
| Mean | 0.67 | 1 | 1.38 | 1.34 | 1.13 |
| SD | 0.76 | 1.20 | 0.64 | 0.73 | 0.79 |
| Median | 1 | 0.5 | 1 | 1 | 1 |
| Min | 0 | 0 | 0 | 0 | 0 |
| Max | 3 | 3 | 3 | 3 | 3 |

Supplementary Table 2. Secondary endpoints in randomised participants only

| | R-BAC+DA | R-BAC | Difference | p-value* |
|---|---|---|---|---|
| **Time to discharge (days)** | | | | |
| Number discharged | 27 | 8 | 19 | |
| Median time to discharge** (95% CI) | 6 (4 to 7) | 4 (2 to-n.a.) | 2 | |
| Hazard ratio*** (95% CI) | | | 1.18 (0.53 to 2.69) | 0.62 |
| **D-dimer (ug/L) FEU** | | | | |
| N | 28 | 6 | | |
| Least-squares mean (log)* (95% CI) | 6.37 (6.01 to 6.74) | 7.55 ( 6.71 to 8.39) | -1.18 (-2.02 to -0.33) | 0.008 |
| Least-square mean** (95% CI) | 586.87 (407.44 to 845.31) | 1903.82 (821.57 to 4411.69) | 0.31 (0.13 to 0.72) | |
| **Lymphocyte count (×$10^9$/L)** | | | | |
| N | 30 | 9 | | |
| Least-squares mean (log)* (95% CI) | -0.06 (-0.25 to 0.12) | -0.46 (-0.82 to -0.1) | 0.4 (0.03 to 0.76) | 0.033 |
| Least-square mean** (95% CI) | 0.94 (0.78 to 1.13) | 0.63 (0.44 to 0.9) | 1.49 (1.03 to 2.13) | |
| **Procalcitonin count (ng/ml)** | | | | |
| N | 26 | 7 | | |
| Least-square mean* (95% CI) | 0.18 (-0.2 to 0.56) | 1.31 (0.56 to 2.05) | -1.13 (-1.88 to -0.37) | 0.005 |

*From log-rank test with treatment as a stratification variable.
**Estimated from Kaplan-Meier curve.
***From Cox proportional hazard model, adjusting for age, baseline CRP and treatment.

Supplementary Table 3. Secondary clinical endpoints

| | | | |
|---|---|---|---|
| **Admission to ICU over 7 days of follow up** | | | |
| N | BAC+DA n=30 | BAC n=69 | |
| | 23.3% | 21.74% | p=0.866 |
| **Length of ICU stay** | | | |
| LSM | 21.25 h | 19.85h | p=0.883 |
| 95% CI | 4.65-37.84 | 8-31.7 | |
| **Admission to ICU over 35 d follow up** | | | |
| N | 23% | 23.19 | p=0.983 |
| LSM | 55.21 h | 60.6 h | P=0.905 |
| 95% CI | -23.59-134.00 h | 4.34-116.86 h | |

| | | | | |
|---|---|---|---|---|
| **Time on Oxygen over 7 days follow-up (hours)** | | | | |
| N | 30 | 69 | | |
| Least-square mean* (95% CI) | 94.32 (72.86 to 115.79) | 88.96 (73.64 to 104.29) | 5.36 (-18.92 to 29.65) | 0.662 |
| **Time on Oxygen over 35 days follow-up (hours)** | | | | |
| N | 30 | 69 | | |
| Least-square mean* (95% CI) | 133.22 (52.01 to 214.43) | 156.35 (98.36 to 214.33) | -23.12 (-115.02 to 67.77) | 0.618 |
| **Proportion of individuals on mechanical ventilation over 7 days follow-up** | | | | |
| N (%) | 5 (16.67) | 9 (13.04) | -4 (3.62) | |
| Odds ratio* (95%CI) | | | 1.36 (0.39 to 4.66) | 0.628 |
| **Proportion of individuals on mechanical ventilation over 35 days follow-up** | | | | |
| N (%) | 5 (16.67) | 9 (13.04) | -4 (3.62) | |
| Odds ratio* (95% CI) | | | 1.36 (0.39 to 4.66) | 0.628 |

*From Logistic regression model, adjusted for age, sex, BMI, baseline CRP, serious condition and treatment.

| | | | |
|---|---|---|---|
| **Proportion of individuals with Superadded Bacterial Pneumonia over 7 days follow-up** | | | |
| N (%) | 1 (3.33) | 3 (4.35) | |
| Odds ratio* (95% CI) | 0.9 (0.08 to 10.21) | | 0.934 |
| **Proportion of individuals with Superadded Bacterial Pneumonia over 35 days follow-up** | | | |
| N (%) | 2 (6.67) | 3 (4.35) | |
| Odds ratio* (95% CI) | 1.81 (0.26 to 12.61) | | 0.548 |

*From Logistic regression model, adjusted for age, sex, BMI, baseline CRP, serious condition, and treatment.

Supplementary Table 4. Safety

| Subject | R-BAC+DA or R-BAC only | Adverse event | Serious? | Relationship to study drug |
|---|---|---|---|---|
| COV002 | Dornase-alfa + BAC | Cough & SOB | No | Not related |
| COV003 | Dornase-alfa + BAC | Mild depression | No | Not related |
| COV003 | Dornase-alfa + BAC | Mild cognitive impairment | No | Not related |
| COV005 | Dornase-alfa + BAC | Struggle to sleep | No | Not related |
| COV005 | Dornase-alfa + BAC | Transaminitis (ALT 91 - NR 10-35 iu/L) | No | Not related |
| COV005 | Dornase-alfa + BAC | Constipation | No | Not related |
| COV007 | Dornase-alfa + BAC | Blood stain in sputum | No | Not related |
| COV012 | Dornase-alfa + BAC | Small Pericardial Effusion | No | Not related |
| COV012 | Dornase-alfa + BAC | Dysphonia | No | Not related |
| COV012 | Dornase-alfa + BAC | Hypercapnia | No | Not related |
| COV013 | Dornase-alfa + BAC | Ulcerative Colitis flare | No | Not related |
| COV013 | Dornase-alfa + BAC | Bradycardia | No | Not related |
| COV015 | Dornase-alfa + BAC | Mechanical Fall | No | Not related |
| COV015 | Dornase-alfa + BAC | Dizziness | No | Not related |
| COV018 | Dornase-alfa + BAC | Dehydration | No | Not related |
| COV018 | Dornase-alfa + BAC | Lower Respiratory Tract Infection | No | Not related |
| COV018 | Dornase-alfa + BAC | Haemoptysis | No | Not related |
| COV020 | Dornase-alfa + BAC | Chest pain | No | Not related |
| COV022 | Dornase-alfa + BAC | Microcytic anaemia | No | Not related |
| COV022 | Dornase-alfa + BAC | Elevated Blood glucose | No | Not related |
| COV023 | Dornase-alfa + BAC | Tachypnoea (PR 32BPM) | No | Not related |
| COV023 | Dornase-alfa + BAC | Hyperglycaemia (BM 14.9) | No | Not related |
| COV031 | Dornase-alfa + BAC | Chest Pain | No | Not related |
| COV035 | Dornase-alfa + BAC | Left leg spasm | No | Not related |
| COV035 | Dornase-alfa + BAC | Rectal bleed due to haemorrhoids | No | Not related |
| COV037 | Dornase-alfa + BAC | Chest Pain | No | Not related |
| COV002 | Dornase-alfa + BAC | Tingling of the mouth | No | Definitely |
| COV035 | Dornase-alfa + BAC | Headache | No | Unlikely |

Supplementary Table 5: Cumulative Summary Tabulations of Serious Adverse Events (SAEs)

| **System Organ Class (SOC)**<br>Preferred Term | **Total** | |
|---|---|---|
| | **Dornase alfa (IMP) arm** | **Best Available Care (control) arm** |
| **Infections and infestations** | 1 | |
| Pyelonephritis | 1 | |
| **Respiratory, thoracic and mediastinal disorders** | 3 | 1 |
| Aspiration pneumonia | | 1 |
| Hospital acquired pneumonia | 1 | |
| Organising pneumonia | 1 | |
| Pulmonary embolism | 1 | |
| **Vascular disorders** | | 1 |
| Acute subdural haematoma | | 1 |
| **Total** | 4 | 2 |

**<u>Appendices:</u>**

Supplementary appendix 1:Protocol

Supplementary appendix 2: Statistical Analysis Plan (SAP)

### Reporting checklist for randomised trial.

Based on the CONSORT guidelines.

#### Instructions to authors

Complete this checklist by entering the page numbers from your manuscript where readers will find each of the items listed below.

Your article may not currently address all the items on the checklist. Please modify your text to include the missing information. If you are certain that an item does not apply, please write "n/a" and provide a short explanation.

Upload your completed checklist as an extra file when you submit to a journal.

In your methods section, say that you used the CONSORTreporting guidelines, and cite them as:

Schulz KF, Altman DG, Moher D, for the CONSORT Group. CONSORT 2010 Statement: updated guidelines for reporting parallel group randomised trials

| | | Reporting Item | Page Number |
|---|---|---|---|
| **Title and Abstract** | | | |
| Title | #1a | Identification as a randomized trial in the title. | 1 |
| Abstract | #1b | Structured summary of trial design, methods, results, and conclusions | 2 |
| **Introduction** | | | |
| Background and objectives | #2a | Scientific background and explanation of rationale | 3-4 |
| Background and objectives | #2b | Specific objectives or hypothesis | 3-4 |
| **Methods** | | | |
| Trial design | #3a | Description of trial design (such as parallel, factorial) including allocation ratio. | 5 |

| | | | |
|---|---|---|---|
| Trial design | #3b | Important changes to methods after trial commencement (such as eligibility criteria), with reasons | N/A |
| Participants | #4a | Eligibility criteria for participants | 6-7 |
| Participants | #4b | Settings and locations where the data were collected | 5 |
| Interventions | #5 | The experimental and control interventions for each group with sufficient details to allow replication, including how and when they were actually administered | 5 |
| Outcomes | #6a | Completely defined prespecified primary and secondary outcome measures, including how and when they were assessed | 8 |
| Outcomes | #6b | Any changes to trial outcomes after the trial commenced, with reasons | N/A |
| Sample size | #7a | How sample size was determined. | 8 |
| Sample size | #7b | When applicable, explanation of any interim analyses and stopping guidelines | SAP page 10 |
| Randomization – Sequence generation | #8a | Method used to generate the random allocation sequence. | |
| Randomization - Sequence generation | #8b | Type of randomization; details of any restriction (such as blocking and block size) | 9 |
| Randomization - Allocation concealment mechanism | #9 | Mechanism used to implement the random allocation sequence (such as sequentially numbered containers), describing any steps taken to conceal the sequence until interventions were assigned | |

| | | | |
|---|---|---|---|
| Randomization - Implementation | #10 | Who generated the allocation sequence, who enrolled participants, and who assigned participants to interventions | Dr. Joanna Porter |
| Blinding | #11a | If done, who was blinded after assignment to interventions (for example, participants, care providers, those assessing outcomes) and how. | NA |
| Blinding | #11b | If relevant, description of the similarity of interventions | NA |
| Statistical methods | #12a | Statistical methods used to compare groups for primary and secondary outcomes | 8 and statistical analysis plan |
| Statistical methods | #12b | Methods for additional analyses, such as subgroup analyses and adjusted analyses | 8 and statistical analysis plan |
| **Results** | | | |
| Participant flow diagram (strongly recommended) | #13a | For each group, the numbers of participants who were randomly assigned, received intended treatment, and were analysed for the primary outcome | Figure 1B and S2 |
| Participant flow | #13b | For each group, losses and exclusions after randomization, together with reason | Figure 1B and S2 |
| Recruitment | #14a | Dates defining the periods of recruitment and follow-up | 10 |
| Recruitment | #14b | Why the trial ended or was stopped | 10 |
| Baseline data | #15 | A table showing baseline demographic and clinical characteristics for each group | TABLE 1 |
| Numbers analysed | #16 | For each group, number of participants (denominator) included in each analysis and | Figure 1B and S2 |

| | | | |
|---|---|---|---|
| | | whether the analysis was by original assigned groups | |
| Outcomes and estimation | #17a | For each primary and secondary outcome, results for each group, and the estimated effect size and its precision (such as 95% confidence interval) | 11 |
| Outcomes and estimation | #17b | For binary outcomes, presentation of both absolute and relative effect sizes is recommended | N/A |
| Ancillary analyses | #18 | Results of any other analyses performed, including subgroup analyses and adjusted analyses, distinguishing pre-specified from exploratory | 11-14 |
| Harms | #19 | All important harms or unintended effects in each group (For specific guidance see CONSORT for harms) | 14 and table S5 and S6 |
| **Discussion** | | | |
| Limitations | #20 | Trial limitations, addressing sources of potential bias, imprecision, and, if relevant, multiplicity of analyses | 16, 18 |
| Generalisability | #21 | Generalisability (external validity, applicability) of the trial findings | 16-18 |
| Interpretation | #22 | Interpretation consistent with results, balancing benefits and harms, and considering other relevant evidence | 17 |
| Registration | #23 | Registration number and name of trial registry | 1 |
| **Other information** | | | |
| Protocol | #24 | Where the full trial protocol can be accessed, if available | Protocol attached |
| Funding | #25 | Sources of funding and other support (such as supply of drugs), role of funders | 18 |

None The CONSORT checklist is distributed under the terms of the Creative Commons Attribution License CC-BY. This checklist can be completed online using https://www.goodreports.org/, a tool made by the EQUATOR Network in collaboration with Penelope.ai

**CONFIDENTIAL**

**Exploristics Limited** Identifier: EXP20012

Clinical Study Identifier: 132333

**Customer:** University College London (UCL)

**Information Type:** Statistical Analysis Plan v1.6

| | |
|---|---|
| Title: A single-site, randomised, controlled, parallel design, open-label investigation of an approved nebulised recombinant human DNase enzyme (dornase alfa) to reduce hyperinflammation in hospitalised participants with COVID-19 (The COVASE trial) | Statistical Analysis Plan v1.6 for 132333 |
| Effective Date: | 12-AUG-2021 |

**Description:** Statistical analysis plan for UCL's COVID-19 trial, COVASE.

**Author's Name, Title and Functional Area:**

Jamie Inshaw, Senior Statistician, Strategic consulting

**Approved by:**

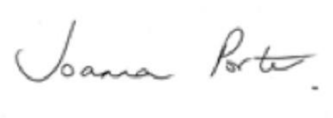

Prof. Joanna Porter Date: 16th August 2021

Chief Investigator

UCL

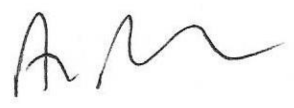

Aiden Flynn Date: 12th August 2021

Director

Exploristics Limited

Copyright 2021 Exploristics Limited. All rights reserved. Unauthorised copying or use of this information is prohibited.

| **Audit Trail For This Document** | | | |
|---|---|---|---|
| **Compiled by/Affiliation** | **Type of Change** | **Date** | **Version** |
| Jamie Inshaw, Exploristics | Creation | 29OCT2020 | 1.0 |
| Jamie Inshaw, Exploristics | Edits following discussions with the client. | 06NOV2020 | 1.1 |
| Jamie Inshaw, Exploristics | Accepted client re-wording and changed the maximum permitted number of individuals to 48 (not 52). | 16DEC2020 | 1.2 |
| Jamie Inshaw, Exploristics | Changed from propensity score matching at the interm to inclusion of all eligible historical controls.<br>Included mock interim tables.<br>Changed "number of comorbidities" in the adjustment and matching analysis to "any key comorbities". | 04JAN2021 | 1.3 |
| Jamie Inshaw, Exploristics | Updated to expand the details of the primary and secondary analyses in preparation for the final analysis | 29JAN2021 | 1.4 |
| Jamie Inshaw, Exploristics | Updated the mechanical ventilation and ventilator-associated pneumonia analysis, as incidence cannot be reliably inferred from the data. Clarified how "time on oxygen" will be calculated. Clarified which covariates will be removed from models if convergence fails or over-fitting present. | 14JUL2021 | 1.5 |
| Jamie Inshaw, Exploristics | Removed oxygen use summary by day as data available in COVASE and HC participants not sufficient for the analysis to be performed.<br>Removed HCs from the | 12AUG2021 | 1.6 |

| | | | |
|---|---|---|---|
| | WHO ordinal scale analysis, as not available in HCs. | | |
| Effective Date | | 12AUG2021 | 1.6 |

**ABBREVIATIONS**

| | |
|---|---|
| AE | Adverse Event |
| APACHE | Acute physiology score + age points + chronic health points |
| BAC | Best Available Care |
| BID | Twice per day |
| CF | Cystic Fibrosis |
| cfDNA | Cell-free DNA |
| CI | Chief Investigator |
| CoV | Coronavirus |
| CRF | Case Report Form |
| CRP | C-reactive protein |
| DNA | Deoxyribonucleic acid |
| DNase | Deoxyribonuclease |
| EudraCT | European Clinical Trials Database |
| FDA | Federal drug administration |
| FVC | Forced vital capacity |
| H3 | Histone 3 |
| ICU | Intensive care unit |
| IL-1 β | Interleukin -1 beta |
| IL-6 | Interleukin-6 |
| IL-8 | Interleukin-8 |
| ISARIC | International Severe Acute Respiratory and Emerging Infection Consortium |
| MPO | Myeloperoxidase |
| MV | Mechanical Ventilation |
| NIHR | National Institute for health research |
| NETs | Neutrophil extracellular traps |
| PCR | Polymerase chain reaction |
| PCT | ProCalcitonin |
| PD | Pharmacodynamics |
| QD | Once per day |
| RECOVERY | Randomised evaluation of COVID-19 therapy |
| RCT | Randomised Controlled Trial |
| SAP | Statistical Analysis Plan |
| SAE | Serious Adverse Event |
| SOFA | Sepsis-related Organ Failure Assessment |
| SOP | Standard Operating Procedure |
| TNFα | Tumour necrosis factor alpha |
| TEAE | Treatment Emergent Adverse Event |
| UCL | University College London |
| UCLH | University College London hospital |
| VAP | Ventilator-Associated Pneumonia |
| WHO | World health organisation |

### 1. INTRODUCTION

The clinical spectrum of SARS-CoV-2 infection (COVID-19) appears to be wide, encompassing asymptomatic infection, mild upper respiratory tract illness (majority of cases) and severe viral pneumonia with respiratory failure and even death in the minority of subjects. In severe viral pneumonia excessive and inappropriate activation of neutrophils can result in the formation of neutrophil extracellular traps (NETs) which exacerbate the clinical course of the pneumonia. These NETs consist of DNA, histones and other components of neutrophils (e.g. myeloperoxidase (MPO)). These NETs are found in the lungs and in the circulation and contribute to organ damage. A treatment that reduces NET formation is likely to reduce the exuberant inflammatory response and thereby improve the clinical course of viral pneumonia and save lives.

NETs have been shown to drive disease in influenza pneumonia as well as in subjects with cystic fibrosis (CF). In addition, their role has been explored in various pre-clinical models of viral infection (mouse and bovine) and shown that reduction of the NETs improves symptoms and increases survival. High neutrophil infiltration is prominent in the lungs of COVID-19 patients and evidence of NET components in the circulation and lung biopsies has been reported in clinical study of COVID-19 patients.

Dornase alfa is a recombinant human DNase I that has been approved since 1994 for the treatment of CF. It is delivered directly to the lungs by nebulisation and has been shown to:

- reduce NETs and inflammation
- reduce the relative risk of developing a respiratory tract infection
- improve pulmonary function in both chronic and acute exacerbation of inflammatory CF

Dornase alfa is safe and well-tolerated in children and adults with CF at doses ranging from 2.5mg QD up to a maximum of 10mg BID.

This study proposes to treat hospitalised subjects with COVID-19 by administration of 2.5mg dornase alfa BID for 7 days. The effect on NETs, inflammation and clinical course will be closely monitored. We expect to see a reduction in circulating NETS and inflammatory biomarkers that will result in clinical benefit. Historic controls will be obtained from an existing database of 120 subjects with COVID-19 that have been admitted to UCL since the beginning of the outbreak.

It is worth noting that dornase alfa can be self-administered at home. Therefore, dornase alfa has the potential to provide benefit in subjects with COVID-19 who have mild disease and are self-isolating and in those discharged from hospital to recuperate at home.

This statistical analysis plan (SAP) describes the key elements of the data handling and analysis strategy that generates evidence of the efficacy and safety of dornase alfa in participants with COVID-19 from the data collected in the study.

### 2. STUDY OBJECTIVE(S) AND ENDPOINT(S)

#### 2.1 Study Objective(s)

**Primary objective**: to assess the effect of dornase alfa on C-reactive Protein (CRP) in hospitalised participants with COVID-19.

**Secondary objective**: to assess the effect of dornase alfa on clinical responses in hospitalised participants with COVID-19.

**Exploratory objective**: to assess the effect of dornase alfa on inflammation, biomarkers of NETs, coagulation, complement activation and haemolysis in hospitalised participants with COVID-19.

#### 2.2 Study Endpoint(s)

##### 2.2.1 Primary Endpoint

The primary endpoint is the levels of acute phase reactant CRP over 7 days follow-up.

##### 2.2.2 Secondary Endpoints

Secondary endpoints include, but are not limited to:

- Levels of acute phase reactant CRP over 35 days follow-up
- Length of hospitalisation from baseline (days)
- Survival at Day35, mortality data collected from EPIC database for both HCs and randomised individuals
- White blood cell count over 7 days follow-up
- Neutrophil count over 7 days follow-up
- Lymphocyte count over 7 days follow-up
- Monocyte count over 7 days follow-up
- Eosinophil count over 7 days follow-up
- Basophil count over 7 days follow-up
- Procalcitonin over 7 days follow-up
- D-dimer count over 7 days follow-up
- Blood pressure over 7 days follow-up
- Pulse rate over 7 days follow-up
- Temperature over 7 days follow-up
- Respiratory rate over 7 days follow-up
- Time on Oxygen over 7 days follow-up
- Time on Oxygen over 35 days follow-up
- Proportion of individuals with Pneumonia over 7 days follow-up
- Ordinal score (WHO scoring tool) over 7 days follow-up (including randomized individuals only)
- Proportion of individuals on Mechanical Ventilation (MV) over 7 days follow-up
- Time on MV over 7 days follow-up
- Length of ICU stay (hours) over 7 days follow-up
- Proportion of individuals with Pneumonia over 35 days follow-up
- Proportion of individuals on Mechanical Ventilation (MV) over 35 days follow-up
- Time on MV over 35 days follow-up
- Length of ICU stay (hours) over 35 days follow-up

##### 2.2.3 Exploratory Endpoints

Exploratory endpoints may be measured in the circulation (blood) and, when these are available, in bronchial secretions (spontaneous expectorant or routine bronchoscopy during MV). They may include, but are not limited to:

- Circulating pro-inflammatory cytokines (e.g. IL-6, TNFα, IL-1β, IL-8)
- Cell-free DNA (cfDNA)
- Circulating histone
- Citrullinated H3
- NET Elisa assay
- NET formation assay
- Coagulation (e.g fibrin, tissue factor, Von Willebrand factor, thrombin, thromboxane A2)
- Complement cascade (e.g C1q)
- Haemolysis (e.g RBC lysis)
- Expression profiling of white blood cells by RNA seq

#### 2.3 Statistical Hypotheses

The statistical hypotheses are defined in terms of a 'zero effect' i.e. no difference between the treatments; the null hypothesis. The subsequent analyses test these statements to determine how much evidence there is to support the null hypothesis. A small p-value can be interpreted as a small probability of observing the result we obtained if there really was no difference between treatments, hence leading to rejection of the null hypothesis.

##### 2.3.1 Primary null hypothesis

- There is no difference in CRP levels between hospitalised COVID-19 positive individuals receiving BAC + dornase alfa and hospitalised COVID-19 positive individuals receiving BAC only.

##### 2.3.2 Secondary null hypotheses

- There is no difference in secondary endpoints between hospitalised COVID-19 positive individuals receiving BAC + dornase alfa and hospitalised COVID-19 positive individuals receiving BAC only.

##### 2.3.3 Exploratory null hypotheses

- There is no difference in exploratory endpoints between hospitalised COVID-19 positive individuals receiving BAC + dornase alfa and hospitalised COVID-19 positive individuals receiving BAC only.

#### 3. STUDY DESIGN

A single-site, randomised, controlled, parallel design, open-label investigation of an approved nebulised recombinant human DNase enzyme (dornase alfa) to reduce hyperinflammation in hospitalised participants with COVID-19 (the COVASE Trial: Figure 1).

Figure 1: COVASE Trial Schematic

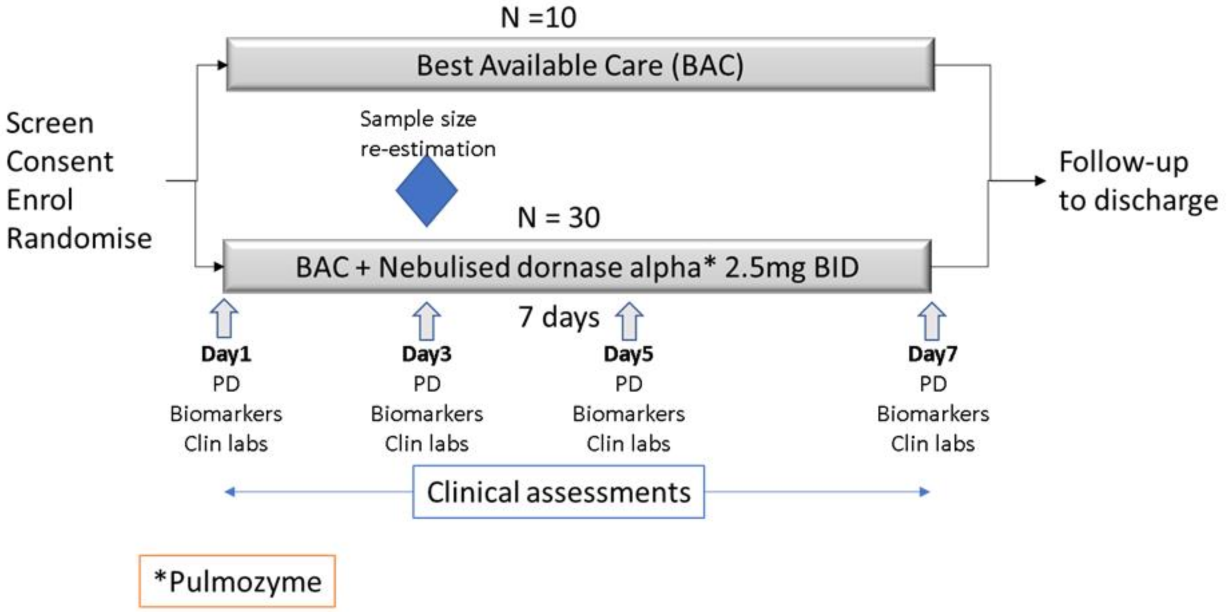

Participants will be screened, consented, enrolled and randomised up to 3 days after they are admitted to the hospital. They will be randomised in a 3:1 ratio to receive BAC + dornase alfa or BAC alone. A total of 40 participants will be enrolled (30 to receive BAC plus dornase alfa and 10 to receive BAC). On Day1 to Day7 of the trial, participants randomised to the active arm, will receive 2.5mg BID nebulised dornase alfa in addition to BAC. On Day1, Day3, Day5 and Day7, blood samples will be drawn in both trial arms in order to test pharmacodynamic endpoints (PD), biomarkers and clin labs. Clinical assessments will be undertaken daily (as per UCLH clinical guidelines). Participants will be followed until discharge or death or a maximum of 28 days follow-up.

A sample size re-estimation is planned when 12 randomised participants have completed Day7 of follow-up. This analysis will ensure that the assumptions made in the sample size calculation remain valid. However, if the variability is higher than expected then up to an additional 10 participants will be enrolled and treated with dornase alfa (up to 48 participants in total).

CRP has been chosen as the Primary Endpoint because it is a clinically important marker of inflammation and is used to make clinical treatment decisions. In addition, it is induced by the over-exuberant inflammation mediated by the NETs and inflammatory histones. CRP is a prognostic marker and correlates with clinical symptoms, inflammation and response to therapy.

Based on clinical judgement, it may be decided to keep some participants on treatment for up to 14 days. In particular, if participants have significant benefits from therapy, but show relapses of the COVID-19 inflammatory state (rising CRP and increasing oxygen requirements in the absence of bacterial infection), on completing 7 days of treatment, then the medical team have the choice of reinstating dornase alfa treatment for up to 7 further days (14 days in total). Blood sampling, as specified, will continue until the last day of dosing with dornase alfa.

**Study controls**

Due to the evolving situation with hospitalised COVID-19 participants, the burden on the NHS and the availability of other COVID-19 trials, it is considered inappropriate to conduct a placebo-controlled study. Therefore, a randomised, controlled, open-label approach where dornase alfa is administered on top of BAC and compared to BAC alone has been adopted.

The data derived from the 10 participants who are randomised to the BAC arm of the study who do not receive dornase alfa will provide control data for all of the study endpoints.

Figure 2 Comparator Data.

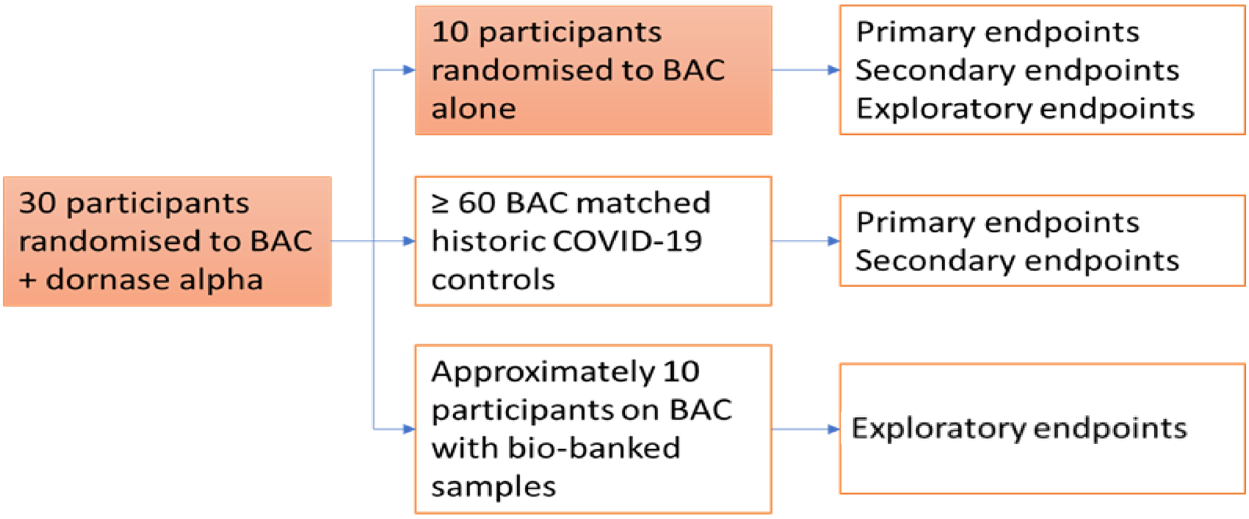

The 30 participants randomized to BAC + dornase alfa will be compared to 10 randomised controls from the COVASE study. In addition, 10 participants in other clinical trials on BAC (bio-banked samples) and 60 participants from a historic database of the first 120 people with COVID-19 treated at UCLH will also be used as comparator data.

However, to enhance the control group, a hybrid approach will be applied such that additional data will be combined with the randomised controls, illustrated in Figure 2. The sample size calculation indicates that 90 evaluable participants are required (60 control:30 active). These control data may include: data from the randomised BAC arm, historic COVID-19 UCLH database, biobanked samples from other ongoing trials and observational trials in COVID-19.
This hybrid strategy allows two things:

- provides comparators for all of the study objectives (including limited data for the exploratory objectives)
- demonstrates that the historic controls are representative/similar to the study population

Comparator data from UCLH is available as historic controls for the Primary and Secondary Endpoints. CRP is routinely measured daily (or on alternate days) in all participants admitted to UCLH and will be used to control for the Primary Endpoint. All of the Secondary Endpoints are also routinely measured and will be available in the database. Participants in the database will be selected to act as controls as follows:

1. Apply the inclusion and exclusion criteria of the COVASE study
2. Additional selection to identify closest matches using a propensity score based on age, gender, BMI, baseline CRP (defined as the last CRP value prior to randomisation, or the first CRP following Dexamethasone for HCs) and whether they have a key comorbidity, defined as one or more of hypertension, diabetes or cardiovascular disease (see Section 11 for details).

At least 60 control participants are expected to be available to act as controls.

Other ongoing and planned trials may be a source of cytokine data as well as biobanked samples that could be used to provide control information on the exploratory endpoints (e.g. ISARIC and RECOVERY Trials) assuming suitable consent is available.

#### 4. PLANNED ANALYSES

There are two planned analyses, an interim analysis after 12 randomised individuals have completed Day7 follow-up and the final analysis, once all randomised individuals have completed follow-up (which is the minimum of discharge, death or 35 days after randomisation). At the interim analysis, only the primary endpoint and safety data will be examined, and no secondary or exploratory endpoints. A promising zones approach [1] (see Section 12 for details) will be followed at the interim analysis stage to identify the conditional power and make a decision as to whether to continue enrolment up to the planned 40 individuals randomised, or whether to increase the planned sample size up to a maximum of 48 randomised individuals. See Section 12 for details.

After the final randomised participant has completed follow-up, a more complete analysis will be undertaken, including examination of endpoints relating to the primary, secondary and exploratory objectives. A full safety analysis will also be carried out at this stage.

#### 5. SAMPLE SIZE CONSIDERATIONS

Sample size calculations were produced using the proc power function in SAS Version 9.4. These were conducted to achieve 80% power to detect a difference in the active arm versus the control group at the 5% level of significance. Based on a mean of 99mg/L in the control group and a common standard deviation of 62mg/L derived from the literature [2, 3], a total sample size of 90 participants would provide sufficient power to detect a greater than a 40% relative difference for CRP in the dornase alfa group compared to the control group. Given the reported average values in severe and non-severe participants and on clinical observations from COVID-19 patients, this difference would be achievable and clinically relevant.

This study will use existing data collected at UCLH from participants admitted with COVID-19 as a comparator group. Participants in the database will be selected to act as controls as follows:

- Apply the inclusion and exclusion criteria of the COVASE study where the appropriate data are available
- Additional selection to identify closest matches

This will give the correct ratio of active versus comparator (ratio of 1:2). To achieve the required power, this would result in 30 participants in the active treatment group and at least 60 in the control. An additional 10 participants will be recruited as a control for the exploratory objectives and to compare the characteristics of enrolled participants with the historical controls. This gives a total of 40 participants enrolled in the study and 60 historical controls.

Participants who drop out of the study will be replaced so the sample size relates to the number of evaluable participants required.

A re-estimation of the sample size will be carried following an interim analysis when 12 randomised participants have completed Day7 follow up. This sample size calculation will be carried out using a promising zones approach [1] (see Section 12 for details).

#### 6. ANALYSIS POPULATIONS

The primary analysis will be conducted using the primary analysis population and is based on the ITT principle. Details on subjects enrolled but not included in the analysis populations will be presented as part of the CONSORT diagram.

##### 6.1 Primary analysis population (Intention to treat)

The primary analysis population will be all evaluable patients randomised to BAC + dornase alfa or BAC only who have at least one post-baseline CRP measurement, as well as matched historical comparators.

##### 6.2 Per protocol population

The per protocol (PP) population will be all evaluable patients randomised to BAC + dornase alfa or BAC only, as well as matched historical comparators, excluding important protocol violations. Important protocol violations will be defined as:

- Initiated dornase-alfa prior to Dexmathasone.
- Withdrew from the study prior to 7 days follow-up.
- Discontinued the study drug prior to discharge from hospital or 7 days follow-up.

##### 6.3 Comparator population

The comparator population will be the matched historical controls and the patients randomised to BAC only.

##### 6.4 Safety population

The safety population will be all patients randomised to either BAC + dornase alfa or BAC only.

##### 6.5 Exploratory analysis population

The exploratory analysis population will be all evaluable patients randomised to BAC + dornase alfa or to BAC only, plus historical patient data from biobanked samples.

### 7. TREATMENT COMPARISONS

The primary treatment comparison will be between individuals randomised to BAC + dornase alfa and the comparator population.

However, comparisons will also be made between the groups forming the comparator population, namely the patients randomised to BAC only and the matched historical controls. These analyses will quantify how similar the matched historical control population is to the participants randomised to BAC only, see section 8.3.1 for details.

### 8. GENERAL CONSIDERATIONS FOR DATA ANALYSES

#### 8.1 Dependent Variables (Endpoint Variables)

For the primary endpoint analysis, the dependent variable will be CRP levels.

#### 8.2 Independent Variables

For the primary endpoint analysis, the independent variables included in the model will be treatment (dornase alfa vs. no dornase alfa), age, gender, BMI, baseline CRP level (defined as the last CRP value prior to randomisation, or the first CRP following Dexamethasone for HCs), time from baseline and whether they have a key comorbidity, defined as one or more of hypertension, diabetes or cardiovascular disease. An interaction term between treatment and time from baseline will also be included in the model.

For secondary endpoint analyses, the same covariates will be included in the models as for the primary endpoint analysis.

#### 8.3 Examination of Subgroups

##### 8.3.1 Primary endpoint analyses

At the interim and final analyses, the primary analysis will be to combine the two sets of individuals on BAC only in the comparator population to the individuals randomised to BAC + dornase alfa.

However, three additional analyses will be carried out at the final analysis for the primary endpoint analysis.

1. A comparison of the BAC + dornase alfa arm to those randomised to BAC only (no historical controls)
2. A comparison of the BAC + dornase alfa arm to matched historical controls (no individuals randomised to BAC only)
3. A comparison of those randomised to BAC only to matched historical controls (within control comparison).

The treatment effect estimate will be compared between the primary analysis and analyses 1. and 2. above. No formal test will be carried out to test the difference in treatment effect between the three analyses, but substantial differences will be noted.

In analysis 3., no treatment effect will be included in the model, since all individuals are on BAC only. Instead, a covariate will be included to indicate whether the individual is a historical control or a randomised individual.

In addition, analyses stratified by BAC treatment will be carried out for the primary endpoint.

###### 8.3.2 Secondary endpoint analyses

A comparison of the secondary endpoint, time on MV over 7 and 35 days follow-up, between treatment groups will be conducted only in participants that received MV.

##### 8.4 Multiple Comparisons and Multiplicity

This study was not powered to detect any effects relating to secondary endpoints. Therefore, the secondary analysis will involve descriptive statistics and thus there is no requirement to consider multiple comparisons in formal hypothesis testing for secondary endpoints.

#### 9. DATA HANDLING CONVENTIONS

##### 9.1 Premature Withdrawal and Missing Data

Mixed models will be used to analyse most endpoints; these models handle missing data naturally. Therefore, all available data will be included in all models.

##### 9.2 Derived and Transformed Data

Continuous endpoints will be assessed for conformance to normality and homogeneity of variance assumptions and the appropriate transformation will be conducted if necessary.

#### 10. DESCRIPTIVE STATISTICS

##### 10.1 Disposition of Subjects

Patient disposition will be listed. Summaries of the following patients will be presented:

- Number of individuals screened
- Number of individuals randomised
- Number of individuals who completed 7 days follow up, or follow-up to discharge from hospital, whichever occurred sooner.
- Number of individuals who withdrew consent or were lost to follow up
- Number of individuals who discontinued dornase alfa for any reason
- Number of individuals with protocol violations, to define PP population
- Number of matched historical controls or biobank samples included in the analysis

##### 10.2 Demographic and Baseline Characteristics

Baseline characteristics will be summarised by treatment (BAC + dornase alfa vs. BAC only) and by population: BAC + dornase alfa, randomised to BAC only, matched historical controls.

The following baseline characteristics will be presented:

- Age
- Gender
- Ethnicity
- BMI
- Baseline CRP (defined as the last CRP value prior to randomisation, or the first CRP following Dexamethasone for HCs)
- Whether they have a key comorbidity, defined as one or more of hypertension, diabetes or cardiovascular disease
- White blood cell count
- Neutrophil count
- Lymphocyte count
- Monocyte count
- Eosinophil count
- Basophil count
- Procalcitonin
- D-dimer count
- Ordinal score (WHO scoring tool), using randomized individuals only.
- Last Pre-Dexamethasone CRP (mg/L)
- Days between diagnosis and hospitalization
- Days between hospitalization and baseline
- Days between Dexamethasone initiation and baseline

#### 11. ANALYSIS CORRESPONDING TO THE STUDY OBJECTIVES

To identify individuals to include in the analysis from the historical controls cohort, the matching procedure will include an initial application of the study inclusion and exclusion criteria to identify the subjects that meet the criteria within the database. Further matching will involve the use of propensity scores to select the controls that most closely match with participants in the active treatment group. The propensity score model will be a logistic regression including all individuals randomised to dornase-alfa and all HCs, with an indicator variable as the outcome to indicate whether the individual is randomised or in the historical control cohort, and covariates included in the model for age, gender, BMI, baseline CRP (defined as the last CRP value prior to randomisation, or the first CRP following Dexamethasone for HCs), and whether they have a key comorbidity, defined as one or more of hypertension, diabetes or cardiovascular disease. The propensity matching will be done using a nearest neighbour approach.
Two controls will be matched for each participant in the active group.
As a supplementary analysis, the propensity score matching will be repeated, but instead of matching on baseline CRP as defined as the last CRP value prior to randomisation, or the first CRP following Dexamethasone for HCs, the matching will be done using the last pre-Dexamethasone CRP measurement, as well as the other factors defined above.

##### 11.1 Primary endpoint analysis

The primary endpoint will be compared between groups using a repeated measures mixed model, adjusted for age, gender, BMI, baseline CRP value (defined as the last CRP measurement prior to randomisation, or the first CRP following Dexamethasone for HCs), time from baseline, and whether they have a key comorbidity, defined as one or more of hypertension, diabetes or cardiovascular disease. An interaction between treatment and time from baseline will also be included in the model. CRP measurements more than 7 days after randomisation (or date of first CRP measurement following Dexamethasone initiation for HCs) will be removed from the analysis, so the comparison is over 7 days.

Due to the treatment by time interaction effect being included in the model, examining the treatment effect alone would be an inappropriate comparison, therefore the least squares means and 95% confidence intervals at the mean follow-up time will be compared between arms, which will take into account both the treatment effect and the treatment by time interaction. These means will be presented on the real scale, e.g. taking the antilog of the least squares mean if the dependent variable has been log-transformed. The difference between the least squares means (on the scale the model was fitted) and the standard error of the difference will be used to generate a treatment z-score, which will be compared against the normal distribution with mean 0 and variance 1, to obtain the two-sided p-value.

In addition, a supplementary analysis will be conducted where the area under the log(CRP) curve will be calculated for each individual up to 7 days of follow-up, divided by the number of days that the individual has been followed up for, to get a standardised area under the curve. The area will be calculated by assuming a linear line between the log(CRP) measurements, regardless of the time between measurements. If measurements are available after day 7, they will be excluded, and the last available log(CRP) measurement prior to day 7 will be used to calculate the area; for example, if the last measurement within 7 days of randomisation for a given individual was on day 4, but they have a day 10 measurement available, the area up to day 4 will be calculated, and divided by 4 to get the standardised area. A linear model will then be fitted, with standardised area under the log(CRP) curve as the outcome, adjusted for age, gender, BMI, log(baseline CRP value) (defined as the last log(CRP) measurement prior to randomisation, or the first log(CRP) following Dexamethasone for HCs) and whether they have a key comorbidity, defined as one or more of hypertension, diabetes or cardiovascular disease. A two-sided p-value will be calculated by comparing the z-score from the Wald test of the treatment effect against the normal distribution with mean 0 and variance 1.

Finally, as a supplementary analysis, the primary analysis will be performed, but including the HCs from the second propensity score matching analysis, where the matching was performed using the last pre-Dexamethasone CRP as opposed to the baseline CRP as defined above. The same model will be fitted for this analysis as the primary analysis, but the model will be adjusted for their last pre-Dexamethasone CRP as opposed to baseline CRP.

Prior to analysis, the primary endpoint will be assessed for conformance to normality assumptions and the appropriate transformation will be conducted if necessary. This model-based approach is likely to be more robust to missing or spurious data. Treatment effect will be declared significant at the 5% level of significance.

#### 11.2 Secondary endpoint analyses

This study was not powered to detect any effects relating to secondary endpoints. Therefore, the secondary analysis will involve descriptive statistics. In general, continuous data will be summarised using the number of individuals, mean (standard deviation), median (1st and 3rd quartiles), minimum and maximum, and categorical data will be represented as frequency counts (percentages).

A further comparison of the secondary endpoints will involve the appropriate general linear models.

For the following secondary endpoints, the same model will be fitted as for the primary endpoint, except with the dependent variable as the secondary endpoint, and adjusting for the baseline level of the secondary endpoint, as opposed to baseline CRP (baseline defined as the last measurement prior to randomisation, or the first measurement following Dexamethasone for HCs):

- Levels of acute phase reactant CRP over 35 days follow-up. The difference from the primary analysis model is that observations will be included up to 35 days after baseline. The treatment comparison will be at the mean follow-up time.
- White blood cell count over 7 days follow-up
- Neutrophil count over 7 days follow-up
- Lymphocyte count over 7 days follow-up
- Monocyte count over 7 days follow-up
- Eosinophil count over 7 days follow-up
- Basophil count over 7 days follow-up
- Procalcitonin over 7 days follow-up
- D-dimer count over 7 days follow-up
- Ordinal score (WHO scoring tool) over 7 days follow-up (using randomized individuals only)

For the following secondary endpoints, a survival analysis will be conducted. The number of events by arm will be compared. The time to event data will be censored at 28 days post last dose (Day35) for the randomised participants and at 35 days after baseline, or the date of the last electronic record, whichever is earlier, for the historical control group. If median time-to-event times exist, the Kaplan-Meier method will be used to estimate the median time-to-event times and the associated 95% confidence intervals. The survival model will include treatment as a stratification variable and significance assessed using a log-rank test. A Cox proportional hazards model will be used to generate a hazard ratio and associated confidence intervals, adjusting for age, baseline CRP value (defined as the last CRP measurement prior to randomisation, or the first CRP following Dexamethasone for HCs) and treatment as a main effect. For the time to discharge from hospital analysis, data will be censored if the participant dies prior to discharge at the date of death.

- Survival at Day35
- Time to discharge from hospital from baseline (days) to Day 35

For the following secondary endpoint, a logistic regression model will be fitted, with age, gender, BMI baseline CRP value (defined as the last CRP measurement prior to randomisation, or the first CRP following Dexamethasone for HCs), whether they have a key comorbidity, defined as one or more of hypertension, diabetes or cardiovascular disease as covariates and treatment included as covariates.

- Proportion of individuals on Mechanical Ventilation (MV) over 7 days follow-up
- Proportion of individuals on Mechanical Ventilation (MV) over 35 days follow-up
- Proportion of individuals with Pneumonia over 7 days follow-up
- Proportion of individuals with Pneumonia over 35 days follow-up

For the following secondary endpoints, a linear regression model will be fitted, with age, gender, BMI, baseline CRP (defined as the last CRP measurement prior to randomisation, or the first CRP following Dexamethasone for HCs), whether they have a key comorbidity, defined as one or more of hypertension, diabetes or cardiovascular disease as covariates and treatment included as covariates.

In addition, a supplementary analysis will be fitted for the length of ICU stay, a logistic regression model will be fitted, with a 1 for the outcome if the individual had any hours in the ICU and a 0 if the individuals had no hours in the ICU, with age, gender, BMI, baseline CRP (defined as the last CRP measurement prior to randomisation, or the first CRP following Dexamethasone for HCs), whether they have a key comorbidity, defined as one or more of hypertension, diabetes or cardiovascular disease as covariates and treatment included as covariates.

- Length of ICU stay (hours) over 7 days follow-up
- Length of ICU stay (hours) over 35 days follow-up
- Time on Oxygen over 7 days follow-up
- Time on Oxygen over 35 days follow-up
- Length of hospitalisation from baseline (days)

For the “time on Oxygen” analyses, regardless of the amount of oxygen received, if there is evidence that any oxygen was provided to the participant on that day, 24 hours will be added to the time on Oxygen.

For the following secondary endpoints, descriptive summary statistics only will be presented:

- Blood pressure over 7 days follow-up
- Pulse rate over 7 days follow-up
- Temperature over 7 days follow-up
- Respiratory rate over 7 days follow-up
- Time on MV over 7 days follow-up. This will only include individuals who were on MV at any point over 7 days follow-up.
- Time on MV over 35 days follow-up. This will only include individuals who were on MV at any point over 35 days follow-up.

Continuous endpoints will be assessed for conformance to normality assumptions and the appropriate transformation will be conducted if necessary.

The secondary outcome analysis will use the primary analysis population.

Note: if models fail to converge or have evidence of over-fitting, covariates will be removed from the models in the following order: serious condition, Sex, Age, BMI, baseline value of measurement.

##### 11.3 Exploratory endpoint analyses

Exploratory analyses will not be performed by Exploristics and will be performed separately.

##### 11.4 Safety analyses

Safety of dornase alfa will be assessed by comparisons of adverse events (AEs), serious adverse events (SAEs), treatment-emergent adverse events (TEAEs) and deaths. The safety population is defined in Section 6.4.

All AEs will be summarized by BAC + dornase alfa/BAC only. A TEAE is an AE that starts or worsens at any time after initiation of study drug on Day1 through to end of follow-up.

For SAEs occurring in ≥5% of individuals, follow-up-adjusted event rates based on events by randomised arm will be provided. The rates will be calculated as the total number of events in the randomised arm divided by the total sum of days follow-up in that randomisation arm, multiplied by 365.25, to give the expected number of events per patient year.

Listings of all TEAEs and SAEs will be provided.

#### 12. INTERIM ANALYSIS

At the interim analysis, baseline demographics, subject disposition and primary endpoint analyses will be carried out. This will occur after the first 12 randomised individuals have completed Day7 follow-up.

A sample size re-estimation will be performed based on a promising zones approach [1].

At the interim stage, the following individuals will be included:

- approximately 9 individuals randomised to BAC + dornase alfa
- approximately 3 individuals randomised to BAC only
- approximately 18 individuals (2 × the number of individuals randomised to BAC + dornase alfa) included from the matched historical control cohorts

This gives a total of approximately 30 individuals included in the analysis. However, if there are insufficient HC data available at the interim analysis to perform propensity score matching (less than 3 × the number of individuals randomised to dornase-alfa), all HCs will be included in the interim analysis.

Prior to an interim analysis sample size re-estimation, the planned sample size at the final analysis is to include:

- 30 individuals randomised to BAC + dornase alfa

- 10 individuals randomised to BAC only
- 60 individuals (2 × the number of individuals randomised to BAC + dornase alfa) included from the matched historical control cohorts

Therefore the planned sample size is 100 individuals.

However, at the interim analysis, a sample size re-estimation will be carried out, and the overall sample size could be increased to a maximum of:

- 36 individuals randomised to BAC + dornase alfa
- 12 individuals randomised to BAC only
- 72 individuals (2 × the number of individuals randomised to BAC + dornase alfa) included from the matched historical control cohorts

Giving a maximum permitted sample size of 120 individuals.

A decision rule for whether to increase the number of individuals randomised into the trial will be derived using a promising zones approach [1]. This approach calculates the power at the interim analysis, conditional on the interim standardised treatment effect size, termed the conditional power. The conditional power at the interim stage will fall into one of three possible zones, a futility zone, a promising zone or a favourable zone.

- If the conditional power is below 0.46 it will be considered to fall into the futility zone. If the conditional power is in the futility zone at the interim analysis, it is unlikely that the final analysis will produce a p-value of <0.05 for the treatment effect, even if the sample size were increased to the maximum permitted sample size of 48 individuals randomised. Therefore, if the conditional power falls in the futility zone at the interim, recruitment will continue as planned to 40 randomised individuals and a final analysis will be performed as originally planned.
- If the conditional power is between 0.46 and 0.8, it will be considered to fall into the promising zone. In this zone, there is less than the 80% power that the study was powered for originally, but if the sample size were to be increased to somewhere between 40 and 48 randomised individuals, the power could be increased to close to 80%, and there would be an increased probability of observing a p-value of <0.05 for the treatment effect at the final analysis.
- If the conditional power is above 0.8, the conditional power will fall into the favourable zone. In this zone, there is more than the 80% power that the study was powered for originally. Therefore, recruitment will be continued to the planed 40 randomised individuals without a change to the planned sample size. There will be no decrease in planned sample size since the treatment effect at the interim analysis could be an over-estimate by chance.

### 13. ATTACHMENTS

#### 13.1 Table of Contents for Data Display Specifications

##### Final analysis

See TFLs_v1.6_clean.docx for final set of Tables, Figures and Listings.

| | |
|---|---|
| **Full title of trial** | A single-site, randomised, controlled, parallel design, open-label investigation of an approved nebulised recombinant human DNase enzyme (dornase alfa) to reduce hyperinflammation in hospitalised participants with COVID-19 (The COVASE trial) |
| **Short title** | Dornase alfa in COVID-19 |
| **Version and date of protocol** | V2.0, 25 April 2020 |
| **Sponsor:** | University College London (UCL) |
| **Sponsor protocol number** | 132333 |
| **Funder (s):** | |
| **EudraCT no** | 2020-001937-11 |
| **ISRCTN / Clinicaltrials.gov no:** | [Insert ISRCTN or Clinicaltrials.gov reference no] |
| **Active treatment** | Dornase alfa (Pulmozyme: recombinant human DNAse I) |
| **PLACEBO IMP(s):** | None |
| **Phase of trial** | Phase IIa |
| **Sites(s)** | UCLH |

**Chief investigator:**
Prof Joanna Porter
UCL Respiratory
The Rayne Building
5 University Street
London WC1E 6JF

**Sponsor Representative**:
Joint Research Office, UCL, 1st Floor Maple House,
149 Tottenham Court Road,
London W1T 7NF
Postal address:
Joint Research Office, UCL
Gower Street,
London WC1E 6BT

#### Protocol Version History

| Version Number | Date | Protocol Update Finalised By (insert name of person): | Reasons for Update |
|---|---|---|---|
| 2.0 | 25 April 2020 | | Response to REC/MHRA |

### 1 Signatures

The Chief Investigator and the JRO have discussed this protocol. The investigator agrees to perform the investigations and to abide by this protocol

The investigator agrees to conduct the trial in compliance with the approved protocol, EU GCP and UK Regulations for CTIMPs (SI 2004/1031; as amended), the UK Data Protection Act (1998), the Trust Information Governance Policy (or other local equivalent), the current UK Policy Framework for Health and Social Care Research , the Sponsor's SOPs and other regulatory requirements as amended.

**Chief investigator**

| | | |
|---|---|---|
| **Professor Joanna Porter**<br>UCL Respiratory | 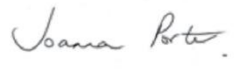 | 25th May 2020 |
| | Signature | Date |
| **Sponsor**<br><br>Dr Nick McNally<br>UCL | 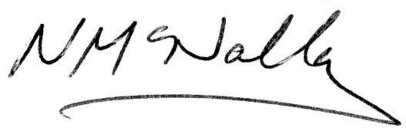 | 25 May 2020 |
| | Signature | Date |

11.2 Sample size and recruitment ... 42
11.2.1 Sample size calculation ... 42
11.2.2 Planned recruitment rate ... 42
11.3 Statistical analysis plan ... 42
11.3.1 Summary of baseline data and flow of participants ... 42
11.3.2 Primary outcome analysis ... 43
11.3.3 Secondary outcome analysis ... 44
11.3.4 Sensitivity and other planned analyses ... 44
11.4 Interim analysis ... 45
11.5 Other statistical considerations ... 45
12 Record keeping and archiving ... 45
13 Oversight committees ... 45
13.1 Data Monitoring Committee (DMC) ... 45
14 Direct Access to Source Data/Documents ... 45
15 Ethics and regulatory requirements ... 46
16 Monitoring requirement for the trial ... 46
17 Finance ... 46
18 Insurance ... 47
19 Publication policy ... 47
20 References ... 47
21 Appendix 1 - Schedule of Assessments ... 49

#### 1.2 List of abbreviations

| | |
|---|---|
| AE | Adverse Event |
| AR | Adverse Reaction |
| ARDS | Acute respiratory distress syndrome |
| BAC | Best Available Care |
| BID | Twice per day |
| CA | Competent Authority |
| CF | Cystic Fibrosis |
| cfDNA | Cell-free DNA |
| CI | Chief Investigator |
| Conmeds | Concomitant medications |
| CoV | Coronavirus |
| CRF | Case Report Form |
| CRO | Contract Research Organisation |
| CRP | C-reactive protein |
| CT | Computer tomography |
| CTA | Clinical Trial Authorisation |
| CTIMP | Clinical Trial of Investigational Medicinal Product |
| DI | Designated Individual |
| DMC | Data Monitoring Committee |
| DNA | Deoxyribonucleic acid |
| DNase | Deoxyribonuclease |
| DSUR | Development Safety Update Report |
| EC | European Commission |
| EMA | European Medicines Agency |
| EU | European Union |
| EUCTD | European Clinical Trials Directive |
| EudraCT | European Clinical Trials Database |
| EudraVigilance | European database for Pharmacovigilance |
| FDA | Federal drug administration |
| FVC | Forced vital capacity |
| GAfREC | Governance Arrangements for NHS Research Ethics |

| | |
|---|---|
| GCP | Good Clinical Practice |
| GMP | Good Manufacturing Practice |
| H3 | Histone 3 |
| HTA | Human Tissue Authority |
| ICF | Informed Consent Form |
| ICU | Intensive care unit |
| IL-1 β | Interleukin -1 beta |
| IL-6 | Interleukin-6 |
| IL-8 | Interleukin-8 |
| ISARIC | International Severe Acute Respiratory and Emerging Infection Consortium |
| ISF | Investigator Site File |
| ISRCTN | International Standard Randomised |
| MA | Marketing Authorisation |
| MHRA | Medicines and Healthcare products Regulatory Agency |
| MPO | Myeloperoxidase |
| MS | Member State |
| MV | Mechanical Ventilation |
| NIHR | National Institute for health research |
| NHS R&D | National Health Service Research & Development |
| NETs | Neutrophil extracellular traps |
| NOCRI | NIHR Office for Clinical Research Infrastructure |
| PCR | Polymerase chain reaction |
| PCT | ProCalcitonin |
| PD | Pharmacodynamics |
| PI | Principal Investigator |
| PO | Purchase order |
| PIS | Participant Information Sheet |
| PL | Product License |
| QD | Once per day |
| QA | Quality Assurance |
| QC | Quality Control |

| | |
|---|---|
| RECOVERY | Randomised evaluation of COVID-19 therapy |
| RCT | Randomised Controlled Trial |
| REC | Research Ethics Committee |
| RSI | Reference Safety Information |
| SAP | Statistical Analysis Plan |
| SAR | Serious Adverse Reaction |
| SAE | Serious Adverse Event |
| SDV | Source Document Verification |
| SOFA | Sepsis-related Organ Failure Assessment |
| SOP | Standard Operating Procedure |
| SMPC | Summary of Product Characteristics |
| SUSAR | Suspected Unexpected Serious Adverse Reaction |
| TNFα | Tumour necrosis factor alpha |
| TRC | Translational Research Collaboration |
| UCL | University College London |
| UCLH | University College London hospital |
| WHO | World health organisation |

#### 1.3 Trial personnel

See protocol cover page for Chief Investigator and Sponsor contact details.

**Statistician**

Aiden Flynn

Exploristics Ltd

20 Rosemount Road

Northern Ireland

BT24 8SY

**External laboratory**

The Francis Crik Institute

Dr Veni Papayannopoulos

1 Midland Rd

London NW1 1AT

### 2 Summary

The clinical spectrum of SARS-CoV-2 infection (COVID-19) appears to be wide, encompassing asymptomatic infection, mild upper respiratory tract illness (majority of cases) and severe viral pneumonia with respiratory failure and even death in the minority of subjects. In severe viral pneumonia excessive and inappropriate activation of neutrophils can result in the formation of neutrophil extracellular traps (NETs) which exacerbate the clinical course of the pneumonia. These NETs consist of DNA, histones and other components of neutrophils (e.g. myeloperoxidase (MPO)). These NETs are found in the lungs and in the circulation and contribute to organ damage. A treatment that reduces NET formation is likely to reduce the exuberant inflammatory response and thereby improve the clinical course of viral pneumonia and save lives.

NETs have been shown to drive disease in influenza pneumonia as well as in subjects with cystic fibrosis. In addition, their role has been explored in various pre-clinical models of viral infection (mouse and bovine) and shown that reduction of the NETs improves symptoms and increases survival. High neutrophil infiltration is prominent in the lungs of COVID-19 patients and evidence of NET components in the circulation and lung biopsies has been reported in clinical study of COVID-19 patients.

Dornase alfa is a recombinant human DNase I that has been approved since 1994 for the treatment of cystic fibrosis (CF). It is delivered directly to the lungs by nebulisation and has been shown to:

- reduce NETs and inflammation
- reduce the relative risk of developing a respiratory tract infection
- improve pulmonary function in both chronic and acute exacerbation of inflammatory CF

Dornase alfa is safe and well-tolerated in children and adults with CF at doses ranging from 2.5mg QD up to a maximum of 10mg BID.

This study proposes to treat hospitalised subjects with COVID-19 by administration of 2.5mg Dornase alfa BID for 7 days. The effect on NETs, inflammation and clinical course will be closely monitored. We expect to see a reduction in circulating NETS and inflammatory biomarkers that will result in clinical benefit. Historic controls will be derived from an existing database of 120 subjects with COVID-19 that have been admitted to UCL since the beginning of the outbreak.

It is worth noting that dornase alfa can be self-administered at home. Therefore, dornase alfa has the potential to provide benefit in subjects with COVID-19 who have mild disease and are self-isolating and in those discharged from hospital to recuperate at home.

| | |
|---|---|
| **Objectives:** | **Primary objective**: to assess the effect of nebulised dornase alfa on C-reactive Protein (CRP) in hospitalised participants with COVID-19.<br><br>**Secondary objective**: to assess the effect of nebulised dornase alfa on clinical responses in hospitalised participants with COVID-19.<br><br>**Exploratory objective**: to assess the effect of nebulised dornase alfa on inflammation, biomarkers of NETs, coagulation, complement activation and haemolysis in hospitalised participants with COVID-19. |
| **Type of trial:** | A single-site, randomised, controlled, parallel design, open-label trial of an approved nebulised recombinant human DNase enzyme (dornase alfa) to reduce hyperinflammation in hospitalised participants with COVID-19 (The COVASE Trial). |
| **Trial design and methods:** | An open-label, randomised, Best-Available-Care (BAC) and historic-controlled trial of nebulised dornase alfa [2.5 mg BID] for 7 days in participants with COVID-19 who are admitted to hospital and are at risk of ventilatory failure (the COVASE study). Controls will include a randomised arm to receive BAC, historic data from UCLH patients with COVID-19 and biobanked samples will be used to demonstrate an effect of dornase alfa. CRP will be measured to assess the effect of dornase alfa on inflammation. Clinical endpoints and biomarkers (e.g. d-dimer) will be used to assess the clinical response. Exploratory endpoints will explore the effects of dornase alfa on features of neutrophil extracellular traps (NETs). |
| **Trial duration per participant:** | Six weeks from consent to last trial assessment. |
| **Estimated total trial duration:** | Four - five months from when first participant enrolled to last participant follow-up. |
| **Planned trial sites:** | Single site |
| **Total number of participants planned:** | 40 participants will be enrolled. A sample size re-estimation is planned to occur approximately one third of the way through the study (e.g. when 12 participants have been randomised). At this point, the decision may be made to randomise additional participants if the variability of the primary endpoint is higher than anticipated. |
| **Main inclusion/exclusion criteria:** | Participants who are hospitalised for COVID-19 will be recruited into the study. |

**Inclusion criteria:**

1. Male and female participants, aged ≥ 18 years.
2. Participants who are hospitalised for suspected Coronavirus (SARS-CoV)-2 infection confirmed by polymerase chain reaction (PCR) test or radiological confirmation with chest CT scan
3. Participants with stable oxygen saturation (>=94%) on supplementary oxygen
4. CRP >= 30 mg/L.
5. Participants will have given their written informed consent to participate in the study and are able to comply with instructions and nebuliser.

**Exclusion criteria**

1. Females who are pregnant, planning pregnancy or breastfeeding.
2. Concurrent and/or recent involvement in other research or use of another experimental investigational medicinal product that is likely to interfere with the study medication within the last 3 months before study enrolment.
3. Serious condition meeting one of the following:
   I. respiratory distress with respiratory rate >=40 breaths/min
   II. oxygen saturation <=93% on high-flow oxygen
4. Require mechanical invasive or non-invasive ventilation at screening
5. Concurrent severe respiratory disease such as asthma, COPD and/or ILD.
6. Any major disorder that in the opinion of the Investigator would interfere with the evaluation of the results or constitute a health risk for the study participant.
7. Terminal disease and life expectancy <12 months without COVID-19.
8. Known allergies to the dornase alfa and excipients.
9. Participants who are unable to inhale or exhale orally throughout the entire nebulisation period.

**Statistical methodology and analysis:**

Full details of the planned statistical analysis will be presented in the Statistical Analysis Plan (SAP). All baseline data, demographics, endpoints, safety and tolerability will be summarised overall and by treatment group and by day. In general, continuous data will be summarised using the mean (standard deviation), median (1st and 3rd quartiles), minimum and maximum, and categorical data will be represented as frequency counts (percentages).

For analyses relating to the primary objective, group comparisons will be performed using a repeated measures mixed model, adjusted for baseline factors and with treatment as the main effect. Prior to analysis, all endpoints will be assessed for conformance to normality assumptions and the appropriate transformation will be conducted if necessary. Some exploratory endpoints may only be available in the active treatment group. In this case, a within group analysis will be conducted to compare baseline and post-baseline measurements.

An interim analysis will be conducted when 12 participants have been randomised. The results of the interim analysis will be used to re-estimate the sample size if necessary.

### 3 Background and rationale

**Clinical background**

COVID-19 is a heterogeneous disease caused by infection with SARS-CoV-2 and although the majority of patients (80%) have mild disease, 15% will require oxygen and of these 25% will require ICU, of which 47 – 71% require ventilatory support. Risk factors for severe disease include older age, male sex, obesity and comorbid disease. There are no specific cures for COVID-19 and current care is supportive. A key challenge is to intercept patients early in the course of their disease to prevent deterioration and reduce the numbers that need ventilatory support, a life-saving treatment that is currently only available for a minority of patients.

SARS-CoV-2 is able to directly infect nasal, bronchiolar and alveolar epithelial cells resulting in lung inflammation, characterised, in severe cases, by an over-exuberant inflammatory response, shortness of breath and hypoxaemia. Once SARS-CoV-2 infection progresses to the stage of pneumonia, key pathogenic drivers result in symptoms characteristic of the acute respiratory distress syndrome (ARDS) with a high mortality.

In patients with confirmed COVID-19 pneumonia, 50% developed dyspnoea at 8 days after illness onset (range: 5–13 days). Mortality of these patients is around 4-15%, as a result of ARDS, coagulation dysfunction, and secondary infection that may result in septic shock and multi-organ failure (Zhou et al., Lancet 2020). Cytokines are thought to be key drivers of these pathological events and in particular levels of IL-6 are found to be raised in COVID-19 patients and to correlate with mortality (Gong et al., medRχiv 2020). Anti-IL-6 is seen as a potential therapy in COVID-19 following favourable reports from a clinical study in China.

There is an urgent need to intervene and reduce the inflammatory response of patients with COVID-19 early in the course of their disease and so prevent progression to ICU.

**Scientific background**

Our hypothesis is that in the COVID-19 lung, the release of neutrophil extracellular traps (NETs) by neutrophils promotes lung damage and the induction of pathogenic pro-inflammatory cytokines, such as IL-6 and IL-1. This inflammatory cascade recruits additional neutrophils leading to a pathogenic feedback amplification loop. High neutrophil infiltration is prominent in the lungs of COVID-19 patients and evidence of NET components in the circulation and lung biopsies has been reported in clinical study of COVID-19 patients (Betsy J. Barnes et al., JEM 2020l; Zuo et al. medRxiv preprint doi:https://doi.org/10.1101/2020.04.09.20059626). Based on this evidence, blocking or clearing NETs to treat severe COVID-19 symptoms has now been proposed by an international consortium (Betsy J. Barnes et al., JEM 2020).

NETs are composed of a backbone of decondensed chromatin fibres coated with antimicrobial granular and cytoplasmic proteins, such as myeloperoxidase, neutrophil elastase (NE) and α- defensins. Double-stranded DNA is a major component of NETs, which prevents their degradation. This extracellular DNA is normally broken down by endogenous deoxyribonucleases (DNases) but in severe inflammation these DNases may become overwhelmed by a massive release of NETs and are unable to completely degrade NETs. Our recent work suggest incomplete NET degradation would lead to amplification rather than reduction of inflammation.

While sensing of microbes and viruses promotes inflammation, the release of endogenous host molecules during infection, known as damage-associated molecular patterns (DAMPs), can amplify cytokine induction. Work in Dr Venizelos Papayannopoulos' lab at the Francis Crick institute has identified specific DAMPs that are critical for the induction of pathogenic hyperinflammation during infection. Free circulating histones are key pathogenic factors in microbial sepsis. We recently found that extracellular chromatin acts as a potent pro-inflammatory signal, allowing extracellular chromatin structures called neutrophil extracellular traps (NETs) to induce IL-1β and IL-6, via Toll-like receptor 4 (TLR4) (Tsourouktsoglou et al. *in press*).

Consistently, NET-mediated pathology causes death in murine models of severe pulmonary *Influenza* infection (Pillai et al., Science 2016) and the presence of NETs correlates with flu severity in humans (Zhu et al., J. Infect. Dis. 2018). DNase treatment significantly delayed mortality in severe flu in immune-compromised mice (Pillai et al., Science 2016) and cleared NETs and lowered airway obstruction in severe bovine respiratory syncytial virus (RSV) infection (Cortjens et al., Thorax 2018). Finally, mice that are genetically pre-disposed to NET overproduction following pulmonary infection with a virulent fungal mutant strains can be rescued from lethality with NET formation inhibitors (Papayannopoulos, Nat. Rev. Immunol. 2018). NETs are major drivers of coagulation during sepsis, particularly in the absence of endogenous DNAses, and are regulated by the complement cascade (Jimenez-Alcazar et al., Science 2017).

**Rationale for the trial**

Dornase alfa is a recombinant human DNase enzyme indicated in conjunction with standard therapies for the management of cystic fibrosis (CF) to improve pulmonary function. Dornase alfa degrades extracellular DNA, and so promotes the clearance of NETs and lead to a significant improvement in lung function for treated CF patients by facilitating mucus clearance in the lung. Dornase alfa is approved worldwide as a nebulised formulation, with an excellent safety profile and is well tolerated. The most common side effect is a hoarse voice. Moreover, dornase alfa could be administered in addition to effective antiviral therapy and should not interfere with antiviral drugs that could be used for COVID-19.

By facilitating the clearance of NETs, dornase alfa not only facilitates sputum clearance in CF patients, but has additional anti-inflammatory activity. Dornase alfa has been shown to reduce NETs in the bronchoalveolar lavage (BAL) and sputum of participants with CF (Konstan et al 2012). In the Bronchoalveolar Lavage for the Evaluation of Anti-inflammatory Treatment (BEAT) study, the percentage of neutrophils in bronchoalveolar lavage fluid significantly increased in untreated CF patients ($P<0.02$) while remaining constant in the dornase alfa-treated group. Levels of elastase and IL-8 also significantly increased from baseline in the untreated group ($P<0.007$ and $P<0.02$ for elastase and IL-8, respectively), but remained stable in patients receiving dornase alfa (Konstan and Ratjen, J. Cyst. Fibros. 2012).

There is scientific evidence to support the potential benefits of dornase alfa in COVID-19 infection. Viral sepsis driven by a hyperinflammation is thought to be a major cause of mortality in COVID-19 infection. Interleukin-1β (IL-1β), IL-6 and TNFα are key cytokines in microbial sepsis. Positive outcomes with Roche's Actemra (tocilizumab), an antibody that blocks the pro-inflammatory cytokine interleukin-6 (IL-6), in COVID-19 treatment has led to several anti-inflammatory trials.

Our hypothesis is that nebulised dornase alfa will break down the DNA backbone of NETs in the COVID-19 lung which will promote the degradation of pro-inflammatory extracellular histones and prevent the amplification of the inflammatory response and the resultant lung damage.

Positive data will enable rapid testing into a large clinical trial in the UK and prevent ICU capacity issues faced today. Dornase alfa is a cost-effective drug and is currently available for prescription.

We propose to test this hypothesis with this COVASE Phase IIa trial. We propose that all people with COVID-19 who are admitted to hospital for supplementary oxygen, who showed evidence of systemic inflammation but did not immediately require intubation and ventilation, would be eligible for nebulised dornase alfa, a safe and cost-effective treatment, twice daily for 7 days.

The hypothesis to be tested is illustrated in Figure 1.

Figure 1: Schematic Representation of the Hypothesis to be tested in the COVASE trial

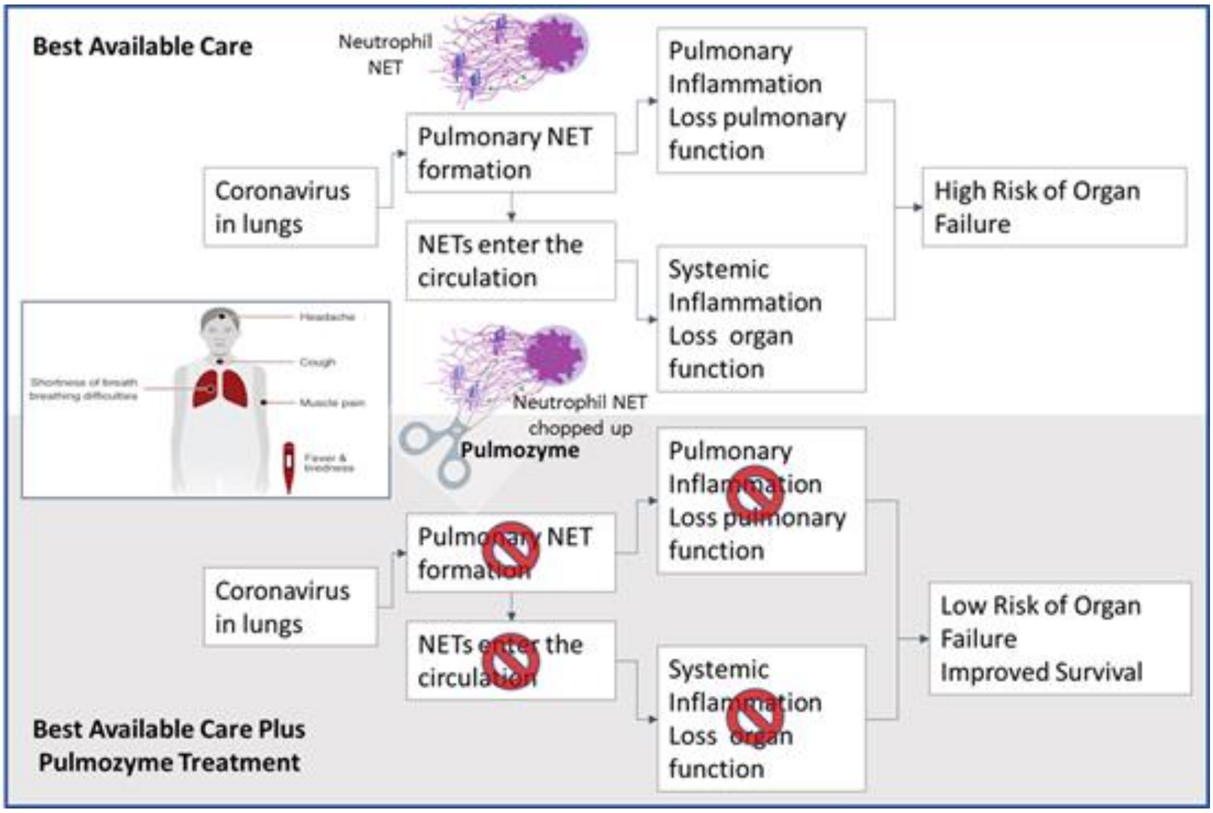

**Potential Real-World evidence**

It is theoretically possible that the CF population could provide Real-World evidence on the effect of dornase alfa on COVID-19. Some participants with CF may develop COVID-19 and may or may not be on dornase alfa. An epidemiological study could determine outcomes in the two groups (CF + COVID-19 not taking dornase alfa and CF + COVID-19 taking dornase alfa). However, currently, the numbers are too low. Considering participants with CF across London, there are a few adults with COVID-19, none of whom have severe disease or have been admitted to ITU. However, they are likely to have been 'shielding' since before the term was coined. So far, no paediatric cases have been reported. However, given that much of the infected paediatric age group elsewhere is asymptomatic or mild, they may not be effectively documented. Therefore, it is not possible at this stage to consider the effect of dornase alfa on the progression of COVID-19 in the setting of CF. All cases of COVID-19 infection in participants with CF will be captured through the national patient registry. This also identifies patients on dornase alfa, so retrospectively these cohorts can be assessed. Data will not be available in high enough numbers for this to be useful during the current pandemic.

**Dornase alfa dosing rational**

The dose to be administered in this trial is 2.5 mg dornase alfa BID (approved) administered with the eRapid nebuliser (or equivalent jet nebuliser connected to an air compressor with an adequate air flow and equipped with a mouthpiece as recommended for dornase alfa-see section 8.3 Table 1). This is twice the standard daily dose (2.5mg QD) and is recommended for older/refractory people with CF. It has been shown to be safe and well tolerated in children and adults with CF. Doses of 2.5mg QD have been administered for years (since 1994) to thousands of people with CF. Higher doses up to 10mg BID have been used in short-term studies with a similar safety and tolerability profile. Dornase alfa has been shown to be safe and well-tolerated during acute exacerbations of CF. Dornase alfa is administered in addition to other treatments for CF (FDA label and EMA SmPC).

Thus, dornase alfa at 2.5 mg BID for seven days is expected to be safe and well-tolerated in hospitalised participants with COVID-19. It is well tolerated and has the potential to block the production of not one but several pro-inflammatory cytokines and acts enzymatically. Moreover, it has been shown not to interfere with anti-viral immune defence (Cortjens et al., Thorax 2018).

#### 3.1 Assessment and management of risk

The table below summarises the risks, frequencies and mitigations of COVASE trial

| **Name of treatment** | **Potential risk** | **Risk Frequency** | **Risk Management** |
|---|---|---|---|
| Recombinant human DNase 1 (dornase alfa) administered by nebulisation | voice alteration<br>pharyngitis<br>rash<br>laryngitis<br>chest pain<br>conjunctivitis | ≥3% | DMC configured to mitigate participant safety and data integrity risks |
| Recombinant human DNase 1 (dornase alfa) administered by nebulisation | rhinitis<br>decrease in FVC of ≥10%*<br>fever<br>dyspnoea | less than 3% | DMC configured to mitigate participant safety and data integrity risks |
| Recombinant human DNase 1 (dornase alfa) administered by nebulisation | There is a low potential immunogenicity risk and antibodies to dornase alfa will not be measured in this study | Low (2-4%) | There have been no reports of anaphylaxis attributed to the administration of dornase alfa. Urticaria, mild to moderate, and mild skin rash have been observed and have been transient. Within all of the studies, a small percentage (average of 2-4%) of people treated with dornase alfa developed serum antibodies to dornase alfa. None of these people developed anaphylaxis, and the clinical significance of serum antibodies to dornase alfa is unknown. |

| | | | |
|---|---|---|---|
| | Failure to recruit patients | unlikely | Feasibility data suggests that sufficient participants can be recruited over the time specified. However, additional sites are available through the NOCRI respiratory TRP that represents 10 of the UK BRCs |
| | Delay in dornase alfa supply | unlikely | UCLH pharmacy to check and put a PO as soon as the grant is awarded and REC/MHRA approval received. |
| | Delay in eRapid supply in the UK | unlikely | UCLH pharmacy to check and put a PO as soon as the grant is awarded and MHRA/REC approval received. |
| | Competing clinical trials | unlikely | There are trials currently ongoing at UCLH that will compete. Trials at UCLH are prioritised and overseen by the COVID-19 committee to ensure equitable recruitment of hospitalised participants to all ongoing trials. Deliverability of trials is also under their management. |
| | Staff sickness due to COVID-19 | unlikely | Additional nursing support is available to cover levels of 25-50% due to COVID-19 infection and time off work. |

*Single measurement only, does not reflect overall FVC changes.

The table below summarise the risks and mitigations of all test above standard care that are being performed in the COVASE Trial:

| **Intervention** | **Potential risk** | **Risk Management** |
|---|---|---|
| Nebulisation of dornase alfa to participants with COVID-19 | As dornase alfa is approved for the treatment of children and adults with CF, we will be using the drug 'off-label' for the first time in participants with COVID-19. There is a low risk that the safety and tolerability in this population may be different to that expected from CF | We will therefore use 'sentinel' dosing for the first three participants enrolled. This means that the three sentinel participants will each commence dosing and safely complete 2 days of treatment individually, before another participant is dosed. Thereafter (participant 4 and onwards) enrolment and dosing may occur in parallel. |

| | | |
|---|---|---|
| Blood draws | Complications that can arise from venepuncture include haematoma formation, nerve damage, pain, haema-concentration, extravasation, iatrogenic anaemia, arterial puncture, petechiae, allergies, fear and phobia, infection, syncope and fainting, excessive bleeding, oedema and thrombus | Experienced hospital staff will be drawing the blood and will minimise these potential risks |

In accordance with the MRC/DH/MHRA Joint Project Risk-adapted Approaches to the Management of Clinical Trials of Investigational Medicinal Products, this trial is categorised as:

Type B = Somewhat higher than the risk of standard medical care

### 4 Objectives and endpoints

**Primary objective**: to assess the effect of nebulised dornase alfa on the inflammatory/immune responses in hospitalised participants with COVID-19

- Primary endpoint:
  - Changes in acute phase reactant (C-Reactive Protein (CRP))

**Secondary objective**: to assess the effect of nebulised dornase alfa on clinical responses in hospitalised participants with COVID-19 compared to control group.

- Secondary endpoints ~~may include, but are not limited to~~:
  - Physical exam and vital signs
  - Whole blood count and differential count
  - Incidence of Mechanical Ventilation (MV)
  - Time on MV
  - ProCalcitonin (PCT)
  - D-dimer
  - Oxygen requirement (oxygen flow or oxygenation index)
  - Length of ICU stay [hours]
  - Length of stay in the hospital [days]
  - Incidence of multi-organ failure according to SOFA (Sepsis-related Organ Failure Assessment)
  - Incidence of Ventilator-Associated Pneumonia (VAP) or hospital acquired pneumonia
  - Acute physiology score + age points + chronic health points (APACHE score)
  - Ordinal score (WHO scoring tool)
  - Survival at Day35

**Exploratory objective**: to assess the effect of nebulised dornase alfa on inflammation, biomarkers of NETs, coagulation, complement activation and haemolysis in hospitalised participants with COVID-19

- Exploratory endpoints may be measured in the circulation (blood) and, when these are available, in bronchial secretions (spontaneous expectorant or routine bronchoscopy during MV). They may include, but are not limited to:
  - Circulating pro-inflammatory cytokines (e.g. IL-6, TNFα, IL-1β, IL-8)
  - Cell-free DNA (cfDNA)
  - Circulating histone
  - Citrullinated H3
  - NET Elisa assay
  - NET formation assay
  - Coagulation (e.g fibrin, tissue factor, Von Willebrand factor, thrombin, thromboxane A2)
  - Complement cascade (e.g C1q)
  - Haemolysis (e.g RBC lysis)
  - Expression profiling of white blood cells by RNA seq

### 5 Trial design

#### 5.1 Overall design

A single-site, randomised, controlled, parallel design, open-label investigation of an approved nebulised recombinant human DNase enzyme (dornase alfa) to reduce hyperinflammation in hospitalised participants with COVID-19 (the COVASE Trial: Figure 2).

Figure 2: COVASE Trial Schematic

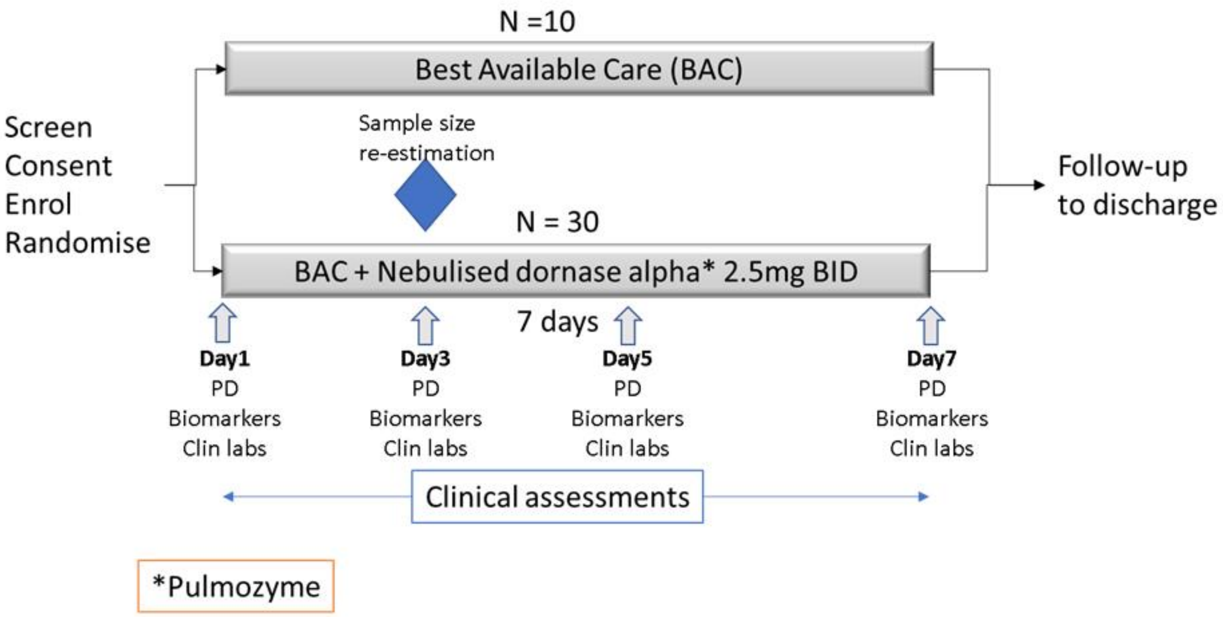

Participants will be screened, consented, enrolled and randomised up to 3 days after they are admitted to the hospital. They will be randomised in a 3:1 ratio to receive BAC + dornase alfa or

BAC alone. A total of 40 participants will be enrolled (30 to receive BAC plus dornase alfa and 10 to receive BAC). On Day1 to Day7 of the trial, participants randomised to the active arm, will receive 2.5mg BID nebulised dornase alfa in addition to BAC. On Day1, Day3, Day5 and Day7, blood samples will be drawn in both trial arms in order to test pharmacodynamic endpoints (PD), biomarkers and clin labs. Clinical assessments will be undertaken daily (as per UCLH clinical guidelines). Participants will be followed until discharge or death or a maximum of 28 days follow-up.

A sample size re-estimation is planned when 12 participants have been randomised. This analysis will ensure that the assumptions made in the sample size calculation remain valid. However, if the variability is higher than expected then up to an additional 10 participants will be enrolled and treated with dornase alfa (up to 50 participants in total).

CRP has been chosen as the Primary Endpoint because it is a clinically important marker of inflammation and is used to make clinical treatment decisions. In addition, it is induced by the over-exuberant inflammation mediated by the NETs and inflammatory histones. CRP is a prognostic marker and correlates with clinical symptoms and response to therapy. Thus, CRP is at the centre of the COVID-19 disease pathway: from NETS to CRP to clinical disease progression.

Based on clinical judgement, it may be decided to keep some participants on treatment for up to 14 days. In particular, if participants have significant benefits from therapy, but show relapses of the COVID-19 inflammatory state (rising CRP and increasing oxygen requirements in the absence of bacterial infection), on completing 7 days of treatment, then the medical team have the choice of reinstating dornase alfa treatment for up to 7 further days (14 days in total). Blood sampling, as specified, will continue until the last day of dosing with dornase alfa.

**Study controls**

Due to the evolving situation with hospitalised COVID-19 participants, the burden on the NHS and the availability of other COVID-19 trials, it is considered inappropriate to conduct a placebo-controlled study. Therefore, a randomised, controlled, open-label approach where dornase alfa is administered on top of BAC and compared to BAC alone has been adopted.

The data derived from the 10 participants who are randomised to the BAC arm of the study who do not receive dornase alfa will provide control data for all of the study endpoints.

Figure 3 Comparator Data.

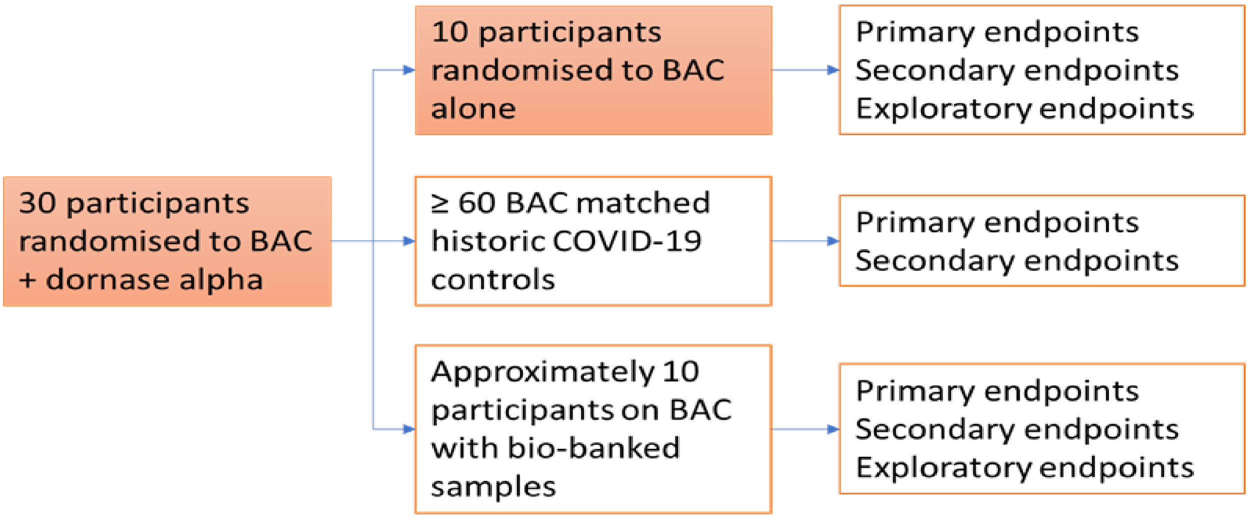

The 30 participants randomized to BAC + dornase alfa will be compared to 10 randomised controls from the COVASE study. In addition, 10 participants in other clinical trials on BAC (bio-banked samples) and 60 participants from a historic database of the first 120 people with COVID-19 treated at UCLH will also be used as comparator data.

However, to enhance the control group, a hybrid approach will be applied such that additional data will be combined with the randomised controls, illustrated in Figure 3. The sample size calculation indicates that 90 evaluable participants are required (60 control:30 active). These control data may include: data from the randomised BAC arm, historic COVID-19 UCLH database, biobanked samples from other ongoing trials and observational trials in COVID-19.

This hybrid strategy allows two things:

- provides comparators for all of the study objectives (including limited data for the exploratory objectives)
- demonstrates that the historic controls are representative/similar to the study population

Comparator data from UCLH is available as historic controls for the Primary and Secondary Endpoints. A database of the first 120 participants with COVID-19 admitted to UCLH is available and being analysed currently. CRP is routinely measured daily (or on alternate days) in all participants admitted to UCLH and will be used to control for the Primary Endpoint. All of the Secondary Endpoints are also routinely measured and will be available in the database. Participants in the database will be selected to act as controls as follows:

1. Apply the inclusion and exclusion criteria of the COVASE study
2. Additional selection to identify closest matches using a propensity score based on age, gender, BMI, baseline CRP, oxygen requirements and the number of comorbidities.

At least 60 control participants are expected to be available to act as controls.

Other ongoing and planned trials may be a source of cytokine data as well as biobanked samples that could be used to provide control information on the exploratory endpoints (e.g. ISARIC and RECOVER Trials) assuming suitable consent is available.

### 6 Off-label use of an Approved Medicinal Product (Pulmozyme)

#### 6.1 Recombinant Human DNase (dornase alfa)

Genentech (a wholly owned subsidiary of Roche) and Chugai Pharmaceutical developed and launched an inhalation solution (Pulmozyme) of dornase alfa, a highly purified recombinant human deoxyribonuclease (dornase alfa), for daily administration in conjunction with standard therapies. The product is indicated for the management of people with cystic fibrosis (CF) to improve pulmonary function. Dornase alfa is safe and well tolerated in adults and children.

Dornase alfa was launched in the US for the management of mild-to-moderate CF in conjunction with standard therapies in January 1994; in December 1996, the FDA expanded approval for use in CF participants with advanced CF. In April 1994, the drug was launched in the UK and Ireland, and by October 1994, it had been launched in France and Germany. In June 2012, dornase alfa was launched in Japan for the improvement of pulmonary function in participants with CF.

In December 2014, the FDA approved the eRapid Nebuliser System from PARI to deliver dornase alfa and to reduce treatment times (three minutes to deliver 2.5mg).

The recommended dosage is one 2.5 mg single-use ampule inhaled once daily using a recommended nebuliser jet nebuliser/compressor system or eRapid™ Nebuliser System. Some participants (older/refractory) benefit from twice daily administration. (FDA label and EMA SmPC).

The most common adverse reactions (occurring in ≥ 3% of participants treated with dornase alfa over placebo) seen in clinical trials in CF participants were: voice alteration, pharyngitis, rash, laryngitis, chest pain, conjunctivitis, rhinitis, decrease in FVC of ≥10%, fever, and dyspnoea.

#### 6.2 Source of dornase alfa, manufacture and distribution

Dornase alfa inhalation solution is a sterile, clear, colourless solution supplied in 30 unit cartons containing 5 foil pouches of 6 single-use ampules. Each 2.5 mL ampule contains 2.5mg of dornase alfa (1 mg/mL): NDC 50242-100-40.

Dornase alfa will be prescribed by the CI (or designee) and dispensed to patients via hospital stock. Alternatively, dornase alfa will be supplied directly from the manufacturer (Roche) as required. Handling and management of dornase alfa will be subject to standard procedures of the pharmacy. The dornase alfa will not be modified in any way, but administered as approved.

#### 6.3 Storage and handling of dornase alfa

Dornase alfa is stored under refrigeration (2°C to 8°C/36°F to 46°F) in their protective foil to protect from light. Dornase alfa should not be used beyond the expiration date stamped on the ampule. Unused ampules must be stored in their protective foil pouch under refrigeration. Dornase alfa must be refrigerated during transport and not exposed to room temperatures for a total time of 24 hours.

#### 6.4 Accountability of dornase alfa

The Drug Accountability Log must be completed to record each dose of dornase alfa dispensed for each trial participant. This log must be retained in the relevant section of the Pharmacy Site File, and a copy must be submitted to the sponsor upon request. It is the responsibility of the Pharmacy Lead to maintain drug accountability records.

All used/unused ampules maybe returned to site pharmacy, to be then updated in the drug accountability log in the pharmacy site file. Following authorisation by the sponsor, drug destruction will be conducted in accordance to local practice, and this will be documented in the drug destruction log in the hospital pharmacy file.

Detailed instructions are contained in the summary of drug arrangements.

#### 6.5 Concomitant medication

Participants in the COVASE trial will continue to receive best available care (BAC) per UCLH guidelines. Dornase alfa will be administered in addition to BAC.

BAC currently consists of symptomatic relief: antipyretics, analgesics and intravenous fluids if needed. In addition, patients may need supplemental oxygen and/or mechanical ventilation.

There is no basis to support a drug-drug interaction risk, as this is a recombinant human protein that is administered directly to the lungs. Systemic exposure is very low and dornase alfa is cleared by proteinases present in the lungs.

There is a potential risk that other medications administered as BAC may affect the endpoints in the study e.g. decrease CRP. However, this cannot be avoided and will be considered in the analysis plan for the data. Furthermore, a sample-size re-estimation is planned in order to take this potential source of unexpected variability into account.

Concomitant medications will be recorded in the participant’s medical records/CRF.

#### 6.6 Post-trial IMP arrangements

No specific arrangements required.

### 7 Selection of participants

People who are at high probability of COVID-19 and admitted to hospital will be identified from inpatient lists and approached, pending COVID PCR results (12 hours). They will be given a patient information sheet (PIS) with details of the clinical trial to read before deciding on entry into the COVASE trial.

#### 7.1 Eligibility of trial participants

##### 7.1.1 Trial participant inclusion criteria

1. Male and female participants, aged ≥ 18 years

2. Participants who are hospitalised for suspected Coronavirus (SARS-CoV)-2 infection confirmed by polymerase chain reaction (PCR) test or radiological confirmation with chest CT
3. Participants with stable oxygen saturation (>=94%) on supplementary oxygen
4. CRP >= 30 mg/L
5. Participants will have given their written informed consent to participate in the study and are able to comply with instructions and nebuliser

##### 7.1.3 Trial participant exclusion criteria

1. Females who are pregnant, planning pregnancy or breastfeeding
2. Concurrent and/or recent involvement in other research or use of another experimental investigational medicinal product that is likely to interfere with the study medication within (specify time period e.g. last 3 months) of study enrolment
3. Serious condition meeting one of the following:
   i. Respiratory distress with respiratory rate >=40 breaths/min
   ii. oxygen saturation<=93% on high-flow oxygen
4. Require mechanical invasive or non-invasive ventilation at screening
5. Concurrent severe respiratory disease such as asthma, COPD and/or ILD
6. Any major disorder that in the opinion of the Investigator would interfere with the evaluation of the results or constitute a health risk for the trial participant
7. Terminal disease and life expectancy <12 months without COVID-19
8. Known allergies to dornase alfa and excipients
9. Participants who are unable to inhale or exhale orally throughout the entire nebulisation period

#### 7.2 Recruitment

Participants will be recruited from inpatients at UCLH.

Participant recruitment will only commence when the trial has been issued with the ‘Open to Recruitment’ letter by the Sponsor.

Recruitment rate is estimated to range from 3 to 6 participants per week. Recruitment is estimated to take 12-15 weeks.

#### 7.3 Informed consent procedure

It is the responsibility of the Investigator, or a person delegated by the Investigator to obtain written informed consent from each participant prior to participation in the trial, following adequate explanation of the aims, methods, anticipated benefits and potential hazards of the trial.

The person taking consent will be GCP trained, suitably qualified and experienced, and will have been delegated this duty by the CI/ PI on the Staff Signature and Delegation of Tasks.

**“Adequate time”** must be given for consideration by the participant before taking part. Due to the rapidly escalating situation, consent will be sought after giving the participant adequate time to consider their decision after being given the study documentation. Patients will be given additional time up to 18 hours if needed. It must be recorded in the medical notes when the participant information sheet (PIS) has been given to the participant.

The Investigator or designee will explain that participants are under no obligation to enter the trial and that they can withdraw at any time during the trial, without having to give a reason.

No clinical trial procedures will be conducted prior to the participant giving consent by signing the Consent form. Consent will not denote enrolment into trial.

A copy of the signed informed consent form will be given to the participant. The original signed form will be retained in the trial file at site and a copy placed in the medical notes.

The PIS and consent form will be reviewed and updated if necessary, throughout the trial (e.g. where new safety information becomes available) and participants will be re-consented as appropriate.

### 8 Trial procedures

#### 8.1 Pre-treatment Assessments

The following trial specific procedures will be carried out after consent and within 3 days of treatment to assess the participant's eligibility:

- Informed consent
- Medical history
- Physical examination
- Vital signs
- Pregnancy test (urine)
- Whole blood count and differential
- Oxygen saturation and record oxygen delivery device if applicable (can be repeated if necessary)
- Oxygen requirement
- Blood draw for PD
- Blood draw for biomarkers
- Clinical Laboratory assessments including CRP, d-dimer and PCT (can be repeated if necessary)
- Concomitant medications

All pre-treatment procedures will be carried out as specified in the schedule of assessments (Appendix 1).

#### 8.2 Randomisation Procedures

Participant randomisation will be undertaken centrally by an independent statistician (ie not the trial statistician) using SAS PROC PLAN according to SOPs. The randomisation schedule will be maintained in a secure, password protected environment, inaccessible to others supporting the trial.

Following participant consent, and confirmation of eligibility (see section 8.1 for pre-treatment assessments) the randomisation procedure described below will be carried out.

Participants are considered to be enrolled into the trial following: consent, pre-treatment assessments (see section 8.1), confirmation of eligibility, completion of the randomisation process, allocation of the participant trial number and treatment by the central coordinating team.

#### 8.3 Treatment Schedule

Dornase alfa will be nebulised at 2.5mg twice per day (12 ± 3 hours apart) for seven days using either:

- the recommended eRapid Nebuliser System, consisting of the eRapid™ Nebuliser Handset with eBase™ Controller OR
- a jet nebuliser connected to an air compressor with an adequate air flow and equipped with a mouthpiece (Table 1).

Table1 Recommended Jet Nebulisers/Compressors

| **Jet nebuliser** | **Compressor** |
|---|---|
| Hudson T Up-draft II with | Pulmo-Aide |
| Marquest Acorn II with | Pulmo-Aide |
| PARI LC Plus with | PARI PRONEB |
| Durable Sidestream with | MOBILAIRE™ |
| Durable Sidestream with | Porta-Neb |

##### 8.3.1 Dose modifications

Based on clinical judgement, it may be decided to extend the dosing period for some participants. For example, a change from 2.5mg BID for 7 days to 2.5mg BID for up to 14 days. In particular, if participants have significant benefits from therapy, but show relapses of the COVID-19 inflammatory state (rising CRP and increasing oxygen requirements in the absence of bacterial infection), on completing 7 days of treatment, then the medical team have the choice of reinstating dornase alfa treatment for up to 7 further days (14 days in total) at 2.5mg BID. Blood sampling will continue until the last day of dosing with dornase alfa.

#### 8.4 Subsequent assessments and procedures

##### 8.4.1 Schedule of assessments

The following assessments and procedures will take place on Day1, Day3, Day5 and Day7 of dosing. In participants who remain on treatment beyond Day7, assessments will occur on Day9, Day11 and Day14 of dosing.

- Eligibility confirmation (at Day1 only)
- Physical examination

- Vital signs (Blood pressure, heart rate, temperature, respiration rate)
- Whole blood count and differential
- Oxygen saturation and record oxygen delivery device if applicable
- Oxygen requirement (oxygen flow or oxygenation index)
- Blood draw and bronchial secretions (when available) for PD
- Blood draw and bronchial secretions (when available) for biomarkers
- Clinical Laboratory assessments including CRP, d-dimer and PCT (can be repeated if necessary)
- Multi-organ failure according to SOFA (Sepsis-related Organ Failure Assessment)
- Acute physiology score + age points + chronic health points (APACHE score) data that has been collected to calculate this score.
- Ordinal score (WHO scoring tool)
- Adverse Events review
- Concomitant Medication review

Other assessments to be recorded if/when they occur:

- Length of ICU stay (hours)
- Length of stay in the hospital (days)
- Length of time on mechanical ventilation (days)
- Ventilator-associated pneumonia (VAP) or hospital-acquired pneumonia
- Survival (days)

**Assessments/procedures at follow-up**. These assessments and procedures will occur before the participant is discharged from the hospital or Day35, whichever comes first.

- Physical examination
- Vital signs (Blood pressure, heart rate, temperature, respiration rate)
- Pregnancy test (urine)
- Whole blood count and differential
- Oxygen saturation and record oxygen delivery device if applicable
- Oxygen requirement (oxygen flow or oxygenation index)
- Blood draw for PD
- Blood draw for biomarkers
- Clinical Laboratory assessments including CRP, d-dimer and PCT (can be repeated if necessary)
- Multi-organ failure according to SOFA (Sepsis-related Organ Failure Assessment)
- Acute physiology score + age points + chronic health points (APACHE score) data that has been collected to calculate this score.
- Ordinal score (WHO scoring tool)
- Adverse Events review (daily)
- Concomitant Medication review

Other assessments to be recorded if/when they occur:

- Length of ICU stay (hours)
- Length of stay in the hospital (days)
- Length of time on mechanical ventilation (days)
- Ventilator-associated pneumonia (VAP) or hospital-acquired pneumonia

- Survival (days)

In participants who are discharged before Day35, a telephone call will occur at Day35 to ask them about their breathing e.g. Are you short of breath? Has your breathing returned to the same level as previously?

A schedule of all trial assessments and procedures is set out in Appendix 1.

#### 8.5 Laboratory Assessments and Procedures

Local laboratories will be used for the primary and secondary assessments and procedures. These include CRP, whole blood count and differential count, proCalcitonin (PCT), and D-Dimer. The samples will be taken as per hospital standard procedures as part of routine clinical care.

The following tests will be carried out at Local Laboratories:

| Laboratory test | Parameters |
|---|---|
| **BLOOD** | |
| Haematology | leukocytes, erythrocytes, haemoglobin, haematocrit, mean corpuscular volume (MCV), mean corpuscular haemoglobin (MCH), platelets, neutrophils, eosinophils, basophils, lymphocytes, monocytes |
| Serum chemistry | glutamate pyruvate transaminase (GPT / ALAT), glutamic-oxaloacetic transaminase (GOT / ASAT), gamma-glutamyl transferase (gamma-GT), alkaline phosphatase, total bilirubin, creatinine, chloride, potassium, sodium, total protein, albumin, Lactate, Renal function: (Creatinine, Urea and Na, K, Cl), Clotting screen including Prothrombin time, Bone profile (ca2+), High-sensitivity troponin, Ferritin, LDH, Creatinine |
| Biomarkers | CRP, PCT, D-dimer |

**PD and biomarkers (exploratory) samples**

The volume of blood required for PD and biomarkers exploratory endpoints is 10ml.

When it is available, bronchial secretions may be collected to measure PD and biomarkers. The bronchial secretion will be obtained due to spontaneous expectoration or route clinical bronchoscopy during MV.

All samples for exploratory endpoints (biomarkers and PD) will be labelled with the unique identifying number (UIN) prior to transfer to The Francis Crick Institute. Samples will be stored in secure UCL research facilities with restricted access under the custodianship of the Chief Investigator or designee until the samples are shipped by courier.

Transfer of samples for exploratory endpoints (biomarkers and PD) to The Francis Crick Institute will be subject to a laboratory agreement in which the Francis Crick Institute will be responsible for restricting use of the samples and data to agreed purposes, maintenance of confidentiality and data security, reporting publications and other outputs to the chief investigator of this trial and restricting onward transfer of samples to a third party.

PD and biomarker samples will be collected under the supervision of the Chief Investigator (or designee) and send to The Francis Crick Institute with courier to a dedicated person in the laboratory of Dr Veni Papayannopoulos. Blood (up to 10 mL – vacutainers- heparin or EDTA) and bronchial secretions will be transferred to The Francis Crick institute. Samples will be processed and stored for further analysis in the Francis Crick Institute Freezer Farm. Tracking of samples will be done with Freezerpro data base. Research samples will be processed in the Crick laboratories by designated research staff within an SOP (COVASE SOP) under the supervision of Dr Veni Papayannopoulos.

**Exploratory samples** (blood draw and bronchial secretion for PD and biomarkers – Appendix I)

Sample processing:

1) Leukocyte pelleting for RNA extraction and sequencing (biomarker).

2) Plasma collected for: Haemolysis (biomarker); cytokine analysis by ELISA and multiplex panel for 67 markers of inflammation ( biomarker); coagulation markers such fibrin, tissue factor, Von Willebrand factor, thrombin, thromboxane A2 (biomarker), complement cascade markers such as C1q, (biomarker), quantification of NETs by NET ELISA ( PD) , quantification of cell-free DNA (PD) and in vitro NET formation assays (biomarker).

3) Plasma samples denatured and boiled in SDS for Western immunoblotting to assess neutrophil markers such as neutrophil elastase; myeloperoxidase; histones; citrullinated H3. (PD).

4) Bronchial secretions will be processed according to local procedures for the measurement of biomarkers.

These samples will be handled at containment level 2 according to local guidelines.

**Total blood draw/participant**

| treatment arm | blood at each timepoint | number of blood draw timepoints | Number of days in the trial | total blood over the trial |
|---|---|---|---|---|
| **BAC for 7 days** | 30mL | 6 | 35 days | 180mL |
| **Dornase alfa for 7 days** | 30mL | 6 | 35 days | 180mL |
| **Dornase alfa for 14 days** | 30mL | 9 | 35 days | up to 270mL |

#### 8.6 Clinical Procedures and Data Collection

All study procedures will be carried out by UCLH medical staff in the hospital and as specified in UCLH guidelines.

Medical examination: The following sites will be examined: head, neck, ears, nose, throat, eyes, chest, lungs, heart, abdomen, skin, and lymph nodes; and the following systems will be assessed: musculoskeletal and neurological.

#### 8.7 Assessment of dornase alfa compliance

Monitoring (e.g. watching participant inhale dornase alfa) and recording this appropriately.

#### 8.8 Discontinuation/withdrawal of participants

In consenting to participate in the trial, participants are consenting to trial treatment, assessments, follow-up and data collection.

**Discontinuation of trial treatment for clinical reasons**

A participant may be withdrawn from trial treatment whenever continued participation is no longer in the participant's best interests, but the reasons for doing so must be recorded. Reasons for discontinuing treatment may include:

- Alternative clinical diagnosis emerges (no longer considered to have primary COVID-19 lung disease).
- Disease progression whilst on therapy.
- Unacceptable toxicity.
- Intercurrent illness which prevents further treatment.
- Patients withdrawing consent to further trial treatment.
- Any alterations in the participant's condition which justifies the discontinuation of treatment in the CI's or designee's opinion.
- Persistent non-compliance to protocol requirements.
- Participant is put on end of life pathway and dies prior to dosing.

The decision to withdraw a participant from treatment must be recorded in the CRF and medical notes, and the sponsor when required should be notified in writing.

**Participant withdrawal from trial treatment or follow-up**

If a participant expresses their wish to withdraw from trial treatment or follow-up, sites should explain the importance of remaining on trial follow-up and seek permission to allow use of routine follow-up data to be used for trial purposes. The importance of safety follow-up should be emphasised to the participant in the Participant Information Sheet.

The decision of the participant to withdraw from treatment or follow-up must be recorded in the CRF and medical notes.

The participant may withhold their reason for withdrawal however, if the participant gives a reason for their withdrawal, this should be recorded.

**Withdrawal of consent to data collection**

If a participant explicitly states that they do not wish to contribute further data to the trial their decision will be respected and recorded in the CRF and medical notes.

#### 8.9 Replacements

40 evaluable participants are required to meet the primary endpoint, Therefore, if a participant withdraws or is withdrawn, a replacement participant may be enrolled to the same treatment arm.

#### 8.10 Stopping rules

The trial may be stopped before completion on the recommendation of the sponsor and CI and following guidance from the DMC following an interim analysis or at any stage during the study.

#### 8.11 Definition of end of trial

Recruitment rate is estimated to range from 3 to 6 participants per week. Recruitment is estimated to take 12-15 weeks.

The expected duration of the trial is 3 – 4 months from recruitment of the first participant to last follow-up visit.

The end of trial is the date of the last visit of the last participant.

### 9 Recording and reporting of adverse events and reactions

Collection, recording and reporting of adverse events (including serious and non-serious events and reactions) to the sponsor will be completed according to the sponsor's SOP (INV/S05).

#### 9.1 Definitions

| Term | Definition |
|---|---|
| Adverse Event (AE) | Any untoward medical occurrence in a participant administered dornase alfa and which does not necessarily have a causal relationship with this treatment. *Therefore, an AE can be any unfavourable or unintended change in the structure (signs), function (symptoms) or chemistry (laboratory data) in a participant to whom dornase alfa has been administered, including occurrences which are not necessarily caused by or related to that product.* |
| Adverse Reaction (AR) | Any untoward and unintended response in a participant to dornase alfa which **is related** to any dose administered to that participant.<br>*This includes medication errors, uses outside of protocol (including misuse and abuse of product)* |
| Serious Adverse Event (SAE), Serious Adverse Reaction (SAR) or Unexpected Serious Adverse Reaction | Any adverse event, adverse reaction or unexpected adverse reaction, respectively, that:<br>• results in death,<br>• is life-threatening*,<br>• requires prolongation of existing hospitalisation**,<br>• results in persistent or significant disability or incapacity, or<br>• consists of a congenital anomaly or birth defect |

| | |
|---|---|
| | * A life-threatening event, this refers to an event in which the participant was at risk of death at the time of the event; it does not refer to an event which hypothetically might have caused death if it were more severe.<br><br>** Hospitalisation is defined as an in-patient admission, regardless of length of stay. Hospitalisation for pre-existing conditions, including elective procedures do not constitute an SAE.<br><br>Some medical events may jeopardise the subject or may require an intervention to prevent one of the above characteristics/consequences. Such **important medical events** should also be considered as serious.<br><br>The term “**severe**” is often used to describe the intensity of an event or reaction (e.g. mild, moderate or severe) and should not be confused or interchanged with the term “**serious**”. |
| Suspected Unexpected Serious Adverse Reaction (SUSAR) | A serious adverse reaction, the nature, severity or outcome of which is not consistent with the Reference Safety Information. |
| Reference Safety Information (RSI) | A list of medical events that defines which reactions are expected for the IMP being administered to clinical trial subjects, and so do not require expedited reporting to the Competent Authority. It is contained in a specific section in the Summary of product characteristics (SmPC) or the Investigator Brochure (IB). |

#### 9.2 Recording adverse events

All adverse events will be assessed every day and recorded in the medical records in the first instance.

All adverse events will be recorded with clinical symptoms and accompanied with a simple, brief description of the event, including dates as appropriate.

Non-serious Adverse Events (AEs) and Serious Adverse Events (SAEs) related to COVID-19 will not be collected in the CRFs or reported to Sponsor for this trial.

The following events listed below describe anticipated Covid-19 related AEs:

Worsening respiratory failure, fever, venous thrombo-embolic disease, worsening respiratory failure, hospital-acquired secondary infection, organ-failure, lymphopenia, rising CRP, death.

#### 9.3 Assessments of Adverse Events

Each adverse event will be assessed for severity, causality, seriousness and expectedness as described below.

##### 9.3.1 Severity

| **Category** | **Definition** |
|---|---|
| Mild | The adverse event does not interfere with the participant's daily routine, and does not require intervention; it causes slight discomfort |
| Moderate | The adverse event interferes with some aspects of the participant's routine, or requires intervention, but is not damaging to health; it causes moderate discomfort |
| Severe | The adverse event results in alteration, discomfort or disability which is clearly damaging to health |

##### 9.3.2 Causality

The assessment of relationship of adverse events to the administration of dornase alfa is a clinical decision based on all available information at the time of the completion of the case report form.

The differentiated causality assessments will be captured in the trial specific CRF/AE Log and/or SAE form.

The following categories will be used to define the causality of the adverse event:

| **Category** | **Definition** |
|---|---|
| Related | A causal relationship between an IMP/investigational treatment and an adverse event is at least a reasonable possibility, i.e., the relationship cannot be ruled out. |
| Not related | There is no reasonable possibility of a causal relationship between an IMP/investigational treatment and an adverse event. |

##### 9.3.3 Expectedness

| Category | Definition |
|---|---|
| *Expected* | An adverse event which is consistent with the information about dornase alfa listed in the current approved Reference Safety Information (RSI) for the trial. |
| *Unexpected* | An adverse event which is not consistent with the information about dornase alfa listed in the current approved Reference Safety Information (RSI) for the trial. |

* This includes listed events that are more frequently reported or more severe than previously reported

The RSI to be used to assess expectedness is section 4.8 of the SmPC for Pulmozyme (dornase alfa).

##### 9.3.4 Seriousness

All events are assessed for seriousness as defined for an SAE in section 9.1.

#### 9.4 Procedures for recording and reporting Serious Adverse Events

All serious adverse events (SAEs/SARs/SUSARs) will be recorded in the medical records in the first instance. Serious Adverse Events (SAEs) related to COVID-19 will not be collected in the CRFs or reported to Sponsor for this trial.

All other SAEs will be recorded in the CRF, and the sponsor's SAE Recording Log and SAE Reporting Form.

All SAEs will be recorded from randomisation until 24h after last study dose.

All SAEs (except Covid-19 related SAEs specified in section as not requiring reporting to the Sponsor), must be recorded on a serious adverse event (SAE) Reporting Form. The CI/PI or designated individual will complete the sponsor's SAE form and email to the Sponsor at, within 24 h of his/her becoming aware of the event. The Chief or Principal Investigator or designee will respond to any SAE queries raised by the sponsor as soon as possible.

Completed SAE forms must be sent within 24 hours of becoming aware of the event to the Sponsor

**Flow Chart for SAE reporting**

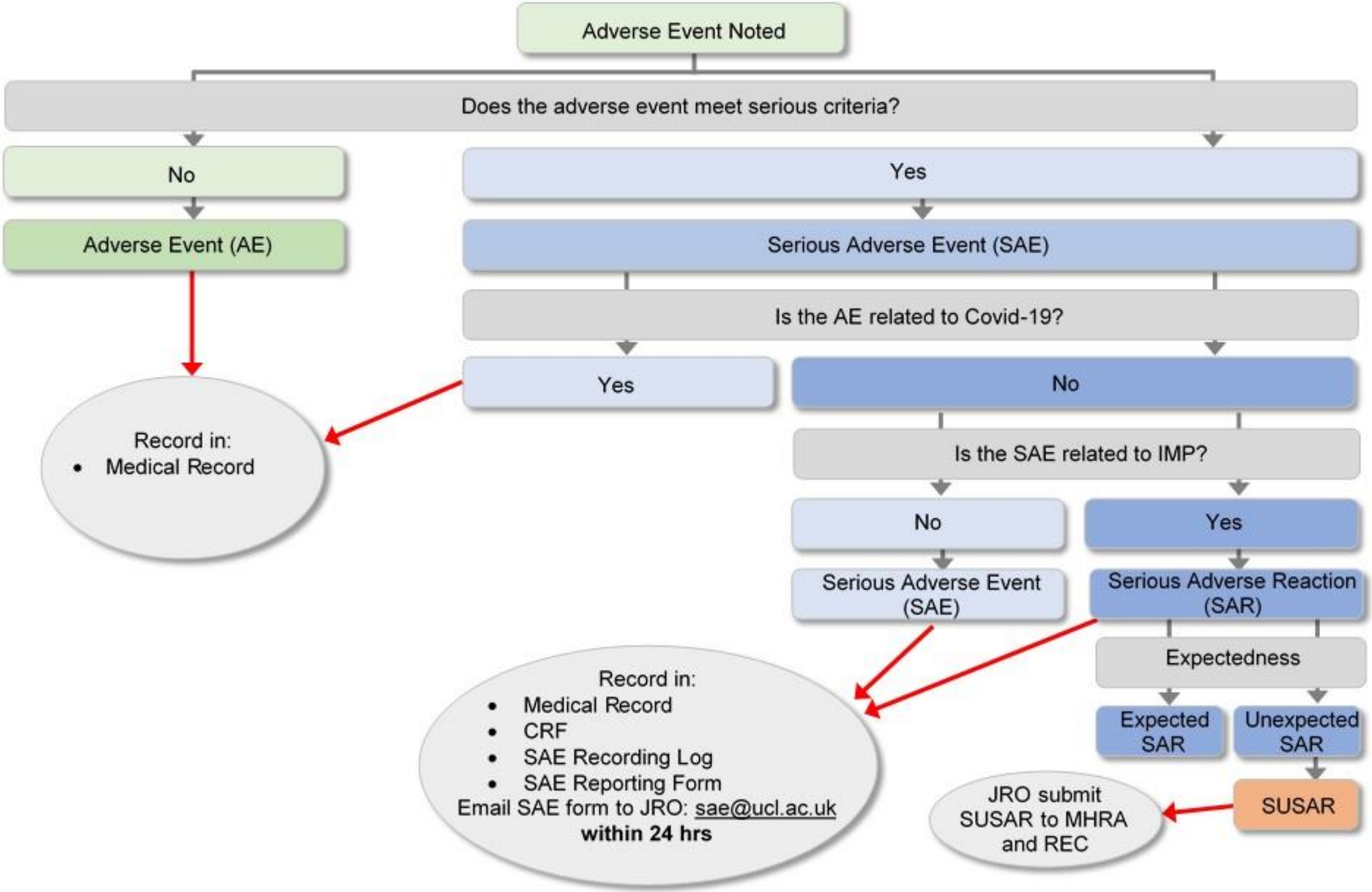

#### 9.5 Serious Adverse Events which do not require immediate reporting

The following events listed below describe anticipated COVID-19 related AEs:

Worsening respiratory failure, fever, venous thrombo-embolic disease, worsening respiratory failure, hospital-acquired secondary infection, organ-failure, lymphopenia, rising CRP, death.

These SAEs will be RECORDED in the participants' medical notes. They will not be recorded in the CRF or SAE Recording Log and SAE Reporting Forms will not be completed and sent to the sponsor.

If the frequency or severity of these events is not consistent with the COVID-19 , the event must be reported to the sponsor as an SAE in the normal way.

#### 9.6 Reporting SUSARs

The sponsor will notify the main REC and MHRA of all SUSARs. SUSARs that are fatal or life-threatening must be notified to the MHRA and REC within 7 days after the sponsor has learned of them. Other SUSARs must be reported to the REC and MHRA within 15 days after the sponsor has learned of them.

#### 9.7 Development Safety Update Reports

The sponsor will provide the main REC and the MHRA with Development Safety Update Reports (DSUR) which will be written in conjunction with the trial team and the Sponsor's office. The report will be submitted within 60 days of the Developmental International Birth Date (DIBD) of the trial each year until the trial is declared ended.

#### 9.8 Pregnancy

Pregnancy is considered to be highly unlikely during the study as participants are hospitalised for the duration of the trial.

There are no adequate and well-controlled studies with dornase alfa in pregnant women. However, animal reproduction studies have been conducted with dornase alfa. In these studies, no evidence of foetal harm was observed in rats and rabbits at doses of dornase alfa up to approximately 600 times the maximum recommended human dose (MRHD).

It is not known whether dornase alfa is present in human milk. In a pharmacokinetic study in Cynomolgus monkeys, levels of dornase alfa detected in milk were less than 0.1% of the maternal serum concentration at 24 hours after dosing [intravenous bolus dose (0.1 mg/kg) of dornase alfa followed by an intravenous infusion (0.080 mg/kg/hr) over a 6-hour period] on post-partum day 14.

Dornase alfa is not contra-indicated in pregnancy or lactation.

In the unlikely event that a female participant or the female partner of a male participant becomes pregnant at any point during the trial, a completed trial specific Pregnancy Reporting Form will be preferably emailed to the Sponsor **,** within 24 hours of his / her becoming aware of the event in line with the Sponsors SOP (JRO/INV/S05). The Chief or Principal Investigator or designee will respond to any queries raised by the sponsor as soon as possible.

| Completed Pregnancy Reporting Forms must be sent within **24 hours** of becoming aware of the event to the Sponsor<br>**Email forms to** |
|---|

The Sponsor must be kept informed of any new developments involving the pregnancy through the completion of a follow-up Pregnancy Reporting Form. Any pregnancy that occurs in a female trial subject during a clinical trial should be followed to termination or to term.

Consent to report information regarding the pregnancy must be obtained from the pregnant participant. A trial-specific pregnancy monitoring information sheet and informed consent form for trial participants and the partners of trial participants must be used for this purpose.

With consent additional information regarding the pregnancy will be collected and reported to the Sponsor, the Sponsor will advise on the length of follow up of the pregnancy/child on a case by case basis.

#### 9.9 Overdose

Overdose is unlikely as dornase alfa will be administered by hospital staff.

Cystic fibrosis participants have received up to 20mg BID for up to 6 days and 10mg BID intermittently (2 weeks on/2 weeks off drug) for 168 days. These doses were well tolerated.

Prescribing information

https://www.gene.com/download/pdf/pulmozyme_prescribing.pdf

#### 9.10 Reporting urgent safety measures and other safety events

If any urgent safety measures are taken the CI/PI or designee shall immediately and in any event no later than 3 days from the date the measures are taken, give written notice to the MHRA, the relevant REC and Sponsor of the measures taken and the circumstances giving rise to those measures.

#### 9.11 Notification of serious breaches to GCP and/or the protocol (SPON/S15)

A “serious breach” is a breach which is likely to affect to a significant degree:

(a) the safety or physical or mental integrity of the participants of the trial, or

(b) the scientific value of the trial.

The sponsor of a clinical trial shall notify the licensing authority in writing of any serious breach of:

(a) the conditions and principles of GCP in connection with that trial, or

(b) the protocol relating to that trial, as amended from time to time, within 7 days of becoming aware of that breach.

The sponsor will be notified immediately of any case where the above definition applies during the trial conduct phase. The sponsor's SOP on the 'Notification of violations, urgent safety measures and serious breaches' will be followed.

### 10 Data management and quality assurance

#### 10.1 Confidentiality

All data will be handled in accordance with the UK Data Protection Act 2018.

The Case Report Forms (CRFs) will bear the participant's initials and UIN. All reports and other results are strictly confidential and access is restricted to relevant healthcare professionals. All of the participant's data will be pseudo-anonymised (according to standard operating procedures) prior to sending data externally for analysis. This will be clearly explained to the participant in the Patient information sheet.

Patient consent for this will be sought.

#### 10.2 Data collection tools and source document identification

Data will be collected on Trial specific case report forms (CRFs) or data collection tools such as CRFs.

Source data are contained in source documents and must be accurately transcribed on to the CRF. Examples of source documents are medical records which include laboratory and other clinical reports etc.

A source document list will be implemented prior to the start of the trial to identify:

which data is to be recorded directly onto the CRF;

which data is recorded firstly into source documents, such as medical notes, and then transcribed into the CRF; and

which data is not to be recorded in the CRF, but only recorded in source documents, e.g., participant questionnaires and diary cards.

#### 10.3 Completing Case Report Forms

All CRFs must be completed and signed by staff that are listed on the site staff delegation log and authorised by the CI/PI or designee to perform this duty. The CI/PI or designee is responsible for the accuracy of all data reported in the CRF.

#### 10.4 Data handling and analysis

A trial specific data management SOP will be in place for the trial. This will contain details of the software to be used for the database, the process of database design, data entry, data quality checks, data queries, data security, database lock and data transfer.

Where data are transferred electronically this will be in accordance with the UK Data Protection Act 1998 as well as UCL Information Security Policy and Trust Information Governance Policy. There will be a documented record of data transfer and measures in place for the recovery of original information after transfer.

### 11 Statistical Considerations

#### 11.1 Outcomes

##### 11.1.1 Primary outcomes

The primary endpoint is the change from baseline in acute phase reactant (C-Reactive Protein (CRP)). This will be measured according to the assessment schedule in section 8.4.1. This endpoint is on a continuous scale.

##### 11.1.2 Secondary outcomes

There are multiple secondary endpoints which will be measured according to the assessment schedule in 8.4.1.

Some of these endpoints are based on change from baseline and are repeated, continuous measures:

- Whole blood count and differential count
- ProCalcitonin (PCT)
- D-dimer
- Oxygen requirement (oxygen flow or oxygenation index)

There is one survival endpoint which is censored/truncated time to event data.

Other secondary endpoints are continuous, absolute measures that are derived only at the end of the assessment period:

- Time on MV
- Length of ICU stay [hours]
- Length of stay in the hospital [days).

The remaining endpoints are on a binary or ordinal scale:

- Incidence of MV
- Incidence of multi-organ failure according to SOFA (Sepsis-related Organ Failure Assessment)
- Incidence of Ventilator-Associated Pneumonia (VAP) or hospital acquired pneumonia
- Ordinal score (WHO scoring tool).

#### 11.2 Sample size and recruitment

##### 11.2.1 Sample size calculation

Sample size calculations were produced using the proc power function in SAS Version 9.4. These were conducted to achieve 80% power to detect difference in the active arm versus the control group at the 5% level of significance. Based on a mean of 99mg/L in the control group and a common standard deviation of 62mg/L derived from the literature (Han et al., 2020; Zhou, 2020), a total sample size of 90 participants would provide sufficient power to detect a greater than a 40% relative difference for CRP in the dornase alfa group compared to the control group. Given the reported average values in severe and non-severe participants and on clinical observations from COVID-19 patients, this difference would be achievable and clinically relevant.

This study will use existing data collected at UCLH from participants admitted with COVID-19 as a comparator group. Participants in the database will be selected to act as controls as follows:

- Apply the inclusion and exclusion criteria of the COVASE study
- Additional selection to identify closest matches

This will give the correct ratio of active versus comparator (ratio of 1:2). To achieve the required power, this would result in 30 participants in the active treatment group and at least 60 in the control. An additional 10 participants will be recruited as a control for the exploratory objectives and to compare the characteristics of enrolled participants with the historical controls. This gives a total of 40 participants enrolled in the study and 60 historical controls.

Participants who drop out of the study will be replaced so the sample size relates to the number of evaluable participants required.

A re-estimation of the sample size will be carried following an interim analysis when 12 participants have been randomised.

##### 11.2.2 Planned recruitment rate

The anticipated recruitment rate is 3-5 participants per week at UCLH. To obtain 40 participants, the recruitment period is likely to last between 8 and 14 weeks. Given the paucity of treatment options and the observed number of cases, the sample size should be easily attainable.

Randomisation methods

Subjects will be allocated to dornase alfa or BAC in a 3:1 ratio in accordance with the randomisation schedule. The randomisation schedule will be based on permuted block randomisation produced using SAS PROC PLAN. Block sizes will vary in multiples of 4.

#### 11.3 Statistical analysis plan

##### 11.3.1 Summary of baseline data and flow of participants

All subjects enrolled in the study will be accounted for in a CONSORT diagram. There will be multiple analysis populations:

1 Primary analysis population - all evaluable patients randomised to dornase alfa + matched historical comparators.

2 Per protocol population - as above but excluding protocol violations.

3 Safety population - all enrolled patients receiving at least one dose of dornase alfa and the comparator groups.

4 Comparator population - the matched historical controls, patients randomised to BAC and historical records linked to biobanked samples.

5 Exploratory analysis population - all evaluable patients randomised to dornase alfa or to BAC plus historical patient data from biobanked samples.

The primary analysis will be conducted using the primary analysis population and is based on the ITT principle. Details on subjects enrolled but not included in the analysis populations will be presented as part of the CONSORT diagram.

The key baseline data that will be used to compare the groups and the analysis populations are age, gender, BMI, baseline CRP, oxygen requirements and the number of comorbidities. In general, continuous data will be summarised using the mean, standard deviation, median, minimum and maximum and categorical data will be represented as frequency counts and percentages.

##### 11.3.2 Primary outcome analysis

The primary outcome analysis will involve the primary endpoint and the primary analysis population. The endpoint will be summarised overall, by treatment group and by day. Data will be summarised using the mean (standard deviation), median (1st and 3rd quartiles), minimum and maximum, and 95% confidence intervals.

The primary endpoint will be compared between groups using a repeated measures mixed model, adjusted for age, gender, BMI, baseline value, oxygen requirements, time from baseline, and the number of comorbidities and with treatment as the main effect and treatment by time as an interaction effect. Prior to analysis, the primary endpoint will be assessed for conformance to normality assumptions and the appropriate transformation will be conducted if necessary. This model-based approach is likely to be more robust to missing or spurious data. Treatment effect will be declared significant at the 5% level of significance.

The primary analysis population comprises the active treatment group and historical matched controls. The matching procedure will include an initial application of the study inclusion and exclusion criteria to identify the subjects that meet the criteria within the database. Further matching will involve the use of propensity scores to select the 60 controls that most closely match with participants in the active treatment group. The propensity score model will include age, gender, BMI, baseline CRP, oxygen requirements and the number of comorbidities and two controls will be matched for each participant in the active group.

##### 11.3.3 Secondary outcome analysis

This study was not powered to detect any effects relating to secondary endpoints. Therefore, the secondary analysis will involve descriptive statistics. In general, continuous data will be summarised using the mean (standard deviation), median (1st and 3rd quartiles), minimum and maximum, and categorical data will be represented as frequency counts (percentages).

A further comparison of the secondary endpoints will involve the appropriate general linear models for binary, continuous or ordinal data with age, gender, BMI, baseline value, oxygen requirements and the number of comorbidities as covariates and treatment as a main effect. This will provide the adjusted means and 95% confidence intervals for the endpoint in the active and control groups. The same model will be used to estimate the difference between the groups and the associated confidence interval.

Continuous endpoints will be assessed for conformance to normality assumptions and the appropriate transformation will be conducted if necessary.

A survival analysis will be conducted on the time to event data. The Kaplan-Meier method will be used to estimate the median survival times and the associated 95% confidence intervals. The time to event data will be censored at 28 days post last dose (Day35) for the randomised participants and at the date of the last electronic record for the historical control group. The survival model will include treatment as a stratification variable. A cox proportional hazards model will be used to generate a hazard ratio and associated confidence intervals, adjusting for age, gender, BMI, baseline value, oxygen requirements and the number of comorbidities as covariates and treatment as a main effect.

The secondary outcome analysis will use the primary analysis population.

##### 11.3.4 Sensitivity and other planned analyses

All primary and secondary analyses will be repeated using the primary analysis population and the per protocol population. The use of the per protocol population is only as a sensitivity analysis to understand the impact of non-compliance.

This study utilises a combination of enrolled and randomised participants, historical controls and banked biosamples. Descriptive statistics will be produced for key baseline variables for all data sources within the comparator population. These will be used to evaluate the comparability of the source data.

There are planned subgroup analyses that relate to secondary objectives. A comparison of the secondary endpoint, time on MV, between treatment groups will be conducted only in participants that received MV. Other subgroup analyses may be performed but these are considered to be exploratory.

Further exploratory analyses are planned for the exploratory endpoints in the study. These analyses will follow the same principles and considerations with regards to the application of statistical approaches for specific data types and data structures. The exploratory analyses will use the exploratory analysis population.

Full details of the planned statistical analysis will be presented in a separate Statistical Analysis Plan (SAP). This will be finalised prior to database lock. The results from all analyses will be presented in the form of tables, figures and listings.

##### 11.4 Interim analysis

An interim analysis will be conducted when 12 participants have been randomised. The results of the interim analysis will be used to re-estimate the sample size if necessary. The interim analysis will be conducted by an independent statistician in a secure, password protected environment. The analysis will involve the production of descriptive statistics for the primary endpoint, AEs and baseline characteristics by study population and by treatment. No formal statistical comparison between the treatment groups will be performed. The sample size re-estimation may result in the recruitment of more subjects than originally planned but not less.

##### 11.5 Other statistical considerations

Any deviation from the original statistical plan will be described and justified in the final report, as appropriate.

#### 12 Record keeping and archiving

At the end of the trial, all essential documentation will be archived securely by the CI and trial sites for a minimum of 25 years from the declaration of end of trial.

Essential documents are those which enable both the conduct of the trial and the quality of the data produced to be evaluated and show whether the site complied with the principles of Good Clinical Practice and all applicable regulatory requirements.

The sponsor will notify sites when trial documentation can be archived. All archived documents must continue to be available for inspection by appropriate authorities upon request.

#### 13 Oversight committees

##### 13.1 Data Monitoring Committee (DMC)

The role of the DMC is to provide advice on data and safety aspects of the trial but not all members are independent. The members consist of the CI, a sponsor physician and a statistician or their respective designees. Meetings of the Committee will be held weekly to review emerging data as well as the results of the interim analysis (Interim Analysis section 11.4), or as necessary to address any issues. The DMC may recommend stopping the study at the interim analysis (or at any stage of the study).

#### 14 Direct Access to Source Data/Documents

The investigator(s)/institution(s) will permit trial-related monitoring, audits, REC review, and regulatory inspection(s), providing direct access to source data/documents. Trial participants are informed of this during the informed consent discussion. Participants will consent to provide access to their medical notes.

### 15 Ethics and regulatory requirements

The sponsor will ensure that the trial protocol, participant information sheet, consent form, GP letter and submitted supporting documents have been approved by the appropriate regulatory body (MHRA in UK) and an appropriate research ethics committee, prior to any participant recruitment. The protocol, all other supporting documents including and agreed amendments, will be documented and submitted for ethical and regulatory approval as required. Amendments will not be implemented prior to receipt of the required approval(s).

Before the site may be opened to recruit participants, the Chief Investigator/Principal Investigator or designee must receive NHS permission in writing from the Trust Research & Development (R&D). It is the responsibility of the CI/ PI or designee at each site to ensure that all subsequent amendments gain the necessary approvals, including NHS Permission (where required) at the site. This does not affect the individual clinician's responsibility to take immediate action if thought necessary to protect the health and interest of individual participants (see section 9.10 for reporting urgent safety measures).

An annual progress report (APR) will be submitted to the REC within 30 days of the anniversary date on which the favourable opinion was given, and annually until the trial is declared ended. The chief investigator will prepare the APR.

Within 90 days after the end of the trial, the CI/Sponsor will ensure that the main REC and the MHRA are notified that the trial has finished. If the trial is terminated prematurely, those reports will be made within 15 days after the end of the trial.

The CI will supply the Sponsor with a report of the clinical trial which complies with the format as defined by the EMA. This will then be uploaded to EudraCT for availability to the MHRA and a copy of the report will be submitted to the main REC, within 1 year after the end of the trial (the exploratory endpoints will be reported separately).

### 16 Monitoring requirement for the trial

The sponsor will determine the appropriate level and nature of monitoring required for the trial. Risk will be assessed on an ongoing basis and adjustments made accordingly.

The degree of monitoring will be proportionate to the objective, purpose, phase, design, size, complexity, blinding, endpoints and risks associated with the trial.

A trial specific oversight and monitoring plan will be established. The trial will be monitored in accordance with the agreed plan.

### 17 Finance

Funding has been sought from LifeArc and outcome is pending.

There are no financial interests by CI, PIs or trial management members in dornase alfa or the eRapid nebuliser.

### 18 Insurance

University College London holds insurance against claims from participants for injury caused by their participation in the clinical trial. Participants may be able to claim compensation if they can prove that UCL has been negligent. However, as this clinical trial is being carried out in a hospital, the hospital continues to have a duty of care to the participant of the clinical trial. University College London does not accept liability for any breach in the hospital's duty of care, or any negligence on the part of hospital employees. This applies whether the hospital is an NHS Trust or otherwise.

Participants may also be able to claim compensation for injury caused by participation in this clinical trial without the need to prove negligence on the part of University College London or another party. Participants who sustain injury and wish to make a claim for compensation should do so in writing in the first instance to the Chief Investigator, who will pass the claim to the Sponsor's Insurers, via the Sponsor's office.

Hospitals selected to participate in this clinical trial shall provide clinical negligence insurance cover for harm caused by their employees and a copy of the relevant insurance policy or summary shall be provided to University College London, upon request.

Indemnity arrangements for the eRapid nebuliser will be in place, with the manufacturer, to cover the malfunction and breakdown of the device.

### 19 Publication policy

There will be weekly team meetings for the staff working on the project. Data generated will be disseminated through medRxiv and bioRxiv and through the Breathing Matters newsletters and website (www.breathingmatters.co.uk and twitter feed @breathingmatter).

#### 21 Appendix 1 - Schedule of Assessments

| Visit No. | 1 | 2 | 3 | 4 | 5 | 6 to 8 | Final visit [8,10] |
|---|---|---|---|---|---|---|---|
| | Day-1 | Day1 | Day3 | Day5 | Day7 | Day9, Day11, Day14 [10] | Day35 or discharge [11] |
| Window of flexibility for timing of visits | ±2 days | | ±1 day | ±1 day | ±1 day | ±1 day | ±2 day |
| Informed Consent | X | | | | | | |
| Medical History | X | | | | | | |
| Physical Examination | X | X | X | X | X | X | X |
| Vital Signs | X | X | X | X | X | X | X |
| Pregnancy test [1] | X | | | | | | X |
| Clinical Laboratory assessments [2] | X | X | X | X | X | X | X |
| Eligibility confirmation | | X | | | | | |
| CRP [3] | X | X | X | X | X | X | X |
| Whole blood count and differential count [4] | X | X | X | X | X | X | X |
| ProCalcitonin (PCT) [3] | X | X | X | X | X | X | X |
| D-Dimer [3] | X | X | X | X | X | X | X |
| Oxygen requirement (oxygen flow or oxygenation index) | X | X | X | X | X | X | X |
| Length of ICU stay (hours) | | X | X | X | X | X | X |
| Length of stay in the hospital (days) | | X | X | X | X | X | X |
| Length of time on mechanical ventilation (days) | | X | X | X | X | X | X |
| Survival (days) | | X | X | X | X | X | X |

| | | | | | | | |
|---|---|---|---|---|---|---|---|
| Ventilator-Associated Pneumonia (VAP) or hospital acquired pneumonia | | X | X | X | X | X | X |
| Acute physiology score + age points + chronic health points (APACHE score) | | X | X | X | X | X | X |
| Ordinal score (WHO scoring tool) | | X | X | X | X | X | X |
| Blood draw for pharmacodynamics (PD) [5] | X | X | X | X | X | X | X |
| Blood draw for biomarkers [6] | X | X | X | X | X | X | X |
| Bronchial secretion for PD and biomarkers [5,6, 7] | | X | X | X | X | X | X |
| nebulised dornase alfa administration [8] | | X | X | X | X | X | |
| Adverse Events review [12] | | X | X | X | X | X | X |
| Concomitant Medication review | X | X | X | X | X | X | X |
| Questions about breathing [9] | | | | | | | X |

1. A urine pregnancy test
2. Clinical Laboratory assessments include: glutamate pyruvate transaminase (GPT / ALAT), glutamic-oxaloacetic transaminase (GOT / ASAT), gamma-glutamyl transferase (gamma-GT), alkaline phosphatase, total bilirubin, creatinine, chloride, potassium, sodium, total protein, albumin, Lactate, Renal function: (Creatinine, Urea and Na, K, Cl), Clotting screen including Prothrombin time, Bone profile (ca2+), High-sensitivity troponin, Ferritin, LDH, Creatinine
3. CRP, PCT and D-Dimer are measured in the clinical laboratory sample
4. Full Blood Count: leukocytes, erythrocytes, haemoglobin, haematocrit, mean corpuscular volume (MCV), mean corpuscular haemoglobin (MCH), platelets, neutrophils, eosinophils, basophils, lymphocytes, monocytes
5. Pharmacodynamics: may include but is not limited to Cell-free DNA (cfDNA), Circulating histone, Citrullinated H3, NET Elisa assay
6. Biomarkers: may include but is not limited to Circulating pro-inflammatory cytokines (e.g. IL-6, TNFα, IL-1β, IL-8), Markers of activation of the coagulation (e.g. fibrin, tissue factor, Von Willebrand factor,

thrombin, thromboxane A2) and complement cascade (e.g. C1q), haemolysis (e.g. red blood cell lysis), NET formation

7. Bronchial secretions will be collected as appropriate for clinical reasons only (e.g. spontaneous expectorant or bronchoscopy during MV)
8. Dornase alfa administered daily at 2.5mg BID by nebuliser
9. In participants who are discharged prior to Day35, a follow-up telephone call will take place to ask about their breathing.
10. In participants who receive dornase alfa (2.5mg BID ) beyond Day7, additional blood draws and procedures will take place on Day9, Day11, Day14
11. Final study visit is at discharge or Day35 whichever comes first
12. Adverse events will be recorded every day and not just on study visit days
